# Genetic and Multi-omic Risk Assessment of Alzheimer’s Disease Implicates Core Associated Biological Domains

**DOI:** 10.1101/2022.12.15.22283478

**Authors:** GA Cary, JC Wiley, J Gockley, S Keegan, L Heath, RR Butler, LM Mangravite, BA Logsdon, FM Longo, A Levey, AK Greenwood, GW Carter, The Emory-Sage-SGC TREAT-AD Center

**Author notes:** These authors participated equally towards the completion of this work (GA Cary, JC Wiley).

## Abstract

Alzheimer’s disease (AD) is the predominant dementia globally, with heterogeneous presentation and penetrance of clinical symptoms, variable presence of mixed pathologies, potential disease subtypes, and numerous associated endophenotypes. Beyond the difficulty of designing treatments that address the core pathological characteristics of the disease, therapeutic development is challenged by the uncertainty of which endophenotypic areas and specific targets implicated by those endophenotypes to prioritize for further translational research. However, publicly-funded consortia driving large-scale open science efforts have produced multiple omic analyses that address both disease risk relevance and biological process involvement of genes across the genome. Here we report the development of an informatic pipeline that draws from genetic association studies, predicted variant impact, and linkage with dementia associated phenotypes to create a genetic risk score. This is paired with a multi omic risk score utilizing extensive sets of both transcriptomic and proteomic studies to identify systems level changes in expression associated with AD. These two elements combined constitute our target risk score that ranks AD risk genomewide. The ranked genes are organized into endophenotypic space through the development of 19 biological domains associated with AD in the described genetics and genomics studies and accompanying literature. The biological domains are constructed from exhaustive gene ontology (GO) term compilations, allowing automated assignment of genes into objectively defined disease-associated biology. This rank and organize approach, performed genome-wide, allows the characterization of aggregations of AD risk across biological domains. The top AD-risk associated biological domains are Synapse, Immune Response, Lipid Metabolism, Mitochondrial Metabolism, Structural Stabilization, and Proteostasis, with slightly lower levels of risk enrichment present within the other 13 biological domains. This provides an objective methodology to localize risk within specific biological endophenotypes, and drill down into the most significantly associated sets of GO-terms and annotated genes for potential therapeutic targets.

## Introduction

Alzheimer’s disease is a complex heterogeneous neurodegenerative disease defined by the extracellular aggregation of amyloid plaques and the intracellular accumulation neurofibrillary tangles comprised of paired helical filaments of hyperphosphorylated tau protein^1-4^. However, while amyloid and tau are hallmarks of the disease, large-scale multi-omic analyses are pointing to the complexity of interwoven biological processes associated with AD pathogenesis. Over a decade ago the National Institute on Aging (NIA) and the Alzheimer’s Association (AA) put together a joint initiative to capture the complexity of AD in the form of a disease ontology, the Common Alzheimer’s and Related Dementias Research Ontology (CADRO)^5^. The goal behind CADRO’s development was to objectively articulate the biological processes and cell types involved in AD pathology and progression. CADRO is used extensively to characterize therapeutic development in AD ^6-12^, and has since been used to track the shift in number and focus of emerging clinical trials. For example, in 2016 most disease modifying therapies in phase III clinical trials were amyloid targeting molecules^5^. However, by 2022 less than one-third of disease modifying therapies in phase III clinical trials are targeting amyloid, with the remainder working across a much broader biological space that includes synaptic targets, neuroprotective agents, metabolic factors, and immune modulators^6^. The diversification is even greater for therapeutic targets in phase I and II clinical trials^6^. Establishing a diverse target portfolio enhances the potential translational impact; the availability of therapeutic targets implicated in an array of disease-linked biology maximizes the potential to intervene through distinct mechanisms, which may be necessary to address the heterogenous AD population and could have the potential to work in coordination^13^. Clinical trials employ CADRO classification to identify the mechanism of action of a therapeutic under investigation. However, the alignment between the gene target of a therapeutic and its ontological classifier is performed manually based upon the judgment of domain experts, and cannot be scaled genome-wide without computationally amenable definitions.

A driving force behind the diversification of the AD target portfolio is an expanding view of AD biology due, in part, to recent efforts that have amassed a wealth of disease-relevant molecular data from a variety of patient cohorts. The Accelerating Medicines Partnership for Alzheimer’s Disease (AMP-AD) consortium, for example, has generated multiple omics datasets from postmortem brain samples (including genomic, transcriptomic, proteomic, metabolomic, etc.) and made these data openly available on the AD Knowledge Portal^14^. These systems-level investigations into AD are a rapidly increasing information domain and each new study contributes large datasets that provide an unbiased view of disease processes across different biological layers. However, each of these datasets can suggest hundreds of genes as potential new therapeutic targets without clear priority. Genome-wide association studies (GWAS) alone have identified over 75 risk loci^15-19^ and analyses of transcriptomic^20-29^ and proteomic^30-35^ data have identified dozens of co-expression modules that consist of hundreds to thousands of genes or proteins each. There are currently over 600 targets that have been nominated by AMP-AD researchers for further therapeutic development (agora.adknowledgeportal.org). Furthermore, these studies each implicate a diverse set of biological processes, pathways, and endophenotypes that are altered in the genesis of, and response to, the late-onset progressive neurodegeneration in AD. The difficulty in performing a unified analysis of these divergent datasets is two-fold: (1) there is no objective and unbiased manner by which to categorize genes into specific AD endophenotypes and (2) there is no integrated, genome-wide methodology to assess and assign AD associated risk.

In this paper we describe data integration across modalities to score, rank, and organize potential AD therapeutic targets at a genome-wide scale, providing the largest resource ever developed to rank and organize AD targets. First, we identified 19 biological domains that capture the preponderance of AD-associated endophenotypes and defined them using an exhaustive set of Gene Ontology (GO) terms, with the intent to keep each domain siloed in a biologically coherent fashion. The extensive sets of GO terms used per biological domain, and the multiplicity of genes annotated to any GO term, provides a classification methodology that spans the majority of the genome. This provides an objective and unbiased organizational strategy to identify gene targets and to assess which AD endophenotypes are especially risk-enriched. Second, we developed a Target Risk Score (TRS), which quantifies dimensions of risk based on genetic association as well as signatures of differential expression in transcriptomic and proteomic data. We show that these tools can be applied to assess which specific genes within large datasets are elevated in disease risk, and to group the most risk-enriched genes within common biological domains, providing a framework for analysis that can be employed across research studies. While we observe that AD risk distributes across all 19 biological domains, we find that the biological domains demonstrating the greatest AD risk association are Synapse, Immune Response, Lipid Metabolisms, Mitochondrial Metabolism, Structural Stabilization, and Proteostasis. Each domain can be examined in more detail by elaborating specific elements of a biological process that are particularly enriched in AD risk—for example, we identify electron transport chain complex I related factors within Mitochondrial Metabolism as one such focal point. The system described here represents the most comprehensive to date, providing genomic coverage of risk mapped onto known AD endophenotypes spanning 27 genetic association studies, transcriptomic signatures from 1,699 brains, proteomic signatures from 1,188 brains, as well as 7,127 Gene Ontology terms structured within the 19 biological domain classifications. These tools are openly available to the research community as a part of the Target Enablement to Accelerate Therapy Development in AD (TREAT-AD) efforts to facilitate the continued diversification of the AD drug development pipeline.

## Results

### Alzheimer’s Disease Biological Domains

The primary goal of defining a structured set of biological domains is to standardize areas of disease-associated biology that can serve as a common reference point for the analysis of large data sets. The CADRO ontology, discussed in the methods section, provided a useful starting place for the development of the biological domains, having been designed for a related purpose, and regularly employed to categorize clinical investigations^6,36^. Leveraging CADRO as a starting place and augmenting this ontology based on literature curation, we established a set of nineteen biological domains that cover the AD endophenotypic space to objectify the processes involved in AD pathogenesis (Supp Table 1). In total we use 7,127 unique GO terms (16.4% of all terms in the GO) to annotate the biological domains, and the number of GO terms annotated to each biological domain varies enormously (Figure 1A, Supp Table 2). The ‘Synapse’ domain requires the largest number of GO terms to define, with 1,379 constitutive GO terms, while ‘Tau Homeostasis’ is the smallest with only 10 GO terms. ‘APP Metabolism’ is the second smallest biological domain; the two smallest biological domains focus on gene-centric processes, requiring fewer terms to annotate than larger domains with broader biological focus, such as ‘Lipid Metabolism’ or ‘Proteostasis’. Each biological domain was designed to be discrete from the others. This is reflected in the sparse overlap of shared GO terms between domains (Figure 1A). The few GO terms that are present in more than one domain are truly inextricable (e.g. the term “mitophagy” defined as “The selective autophagy process in which a mitochondrion is degraded by macroautophagy” legitimately resides in both the Mitochondrial Metabolism and Autophagy domains), in which case the repetition was allowed, as it represents a meaningful intersection of biological areas.

**Fig. 1.**
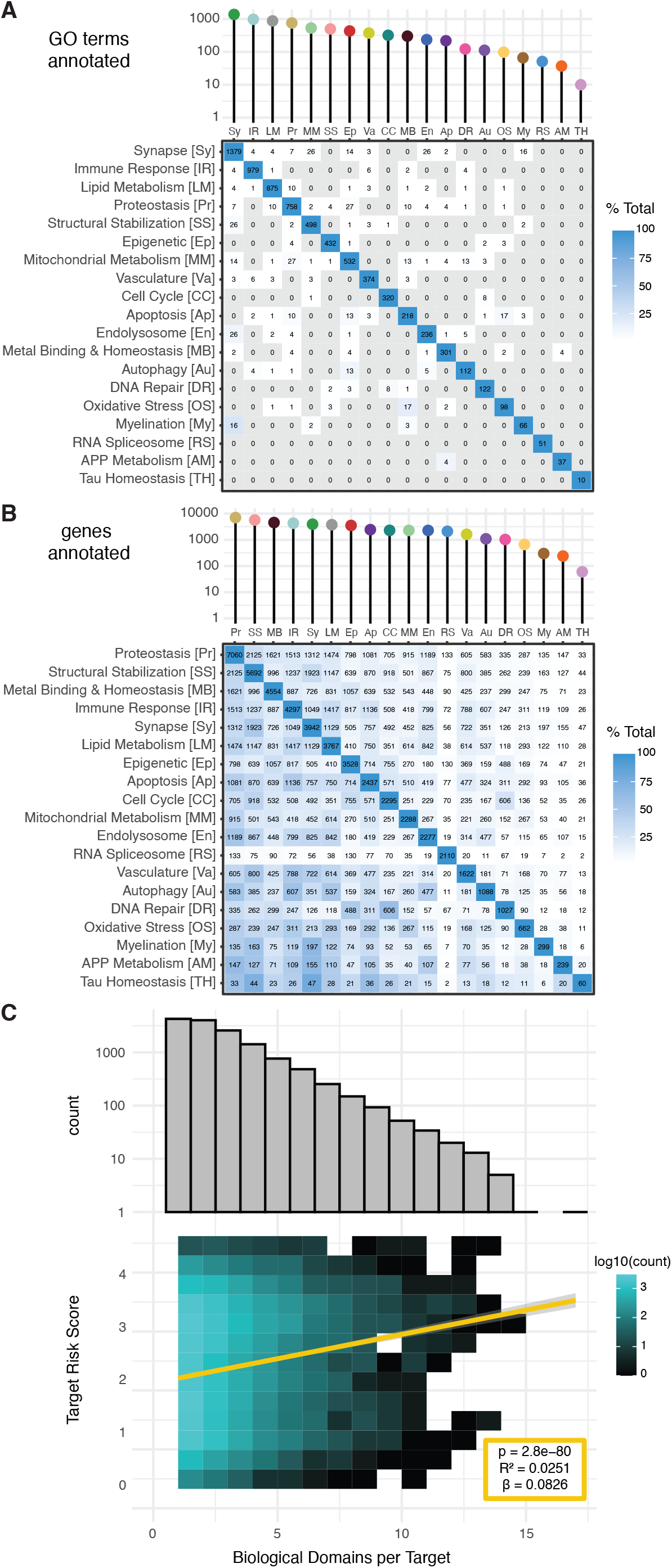
Biological domain demographics. (A) The number of GO terms employed to define each biological domain, shown as an interaction plot, with the total number of terms per domain along the diagonal, and the number of terms shared between domains arranged in the external rows and columns. (B) The number of genes in each biological domain, organized as in (A), with the total genes within each domain along the diagonal, and the pairwise genes shared between the domains heat mapped within the rows and columns. (C) The top histogram shows the frequency of a gene annotation across the 19 biological domains for genes annotated to at least 1 biological domain. The histogram shows a decline in the number of genes annotated to multiple biological domains, with genes mapping to a single biological domain being the most numerous (4,258). The lower plot shows the positive correlation between gene annotation in multiple biological domains and its Target Risk Score, with higher scoring genes participating in multiple biological domains.

The genes annotated to the biological domains reveal a different pattern, with high levels of promiscuity between the domains (Figure 1B). The number of genes annotated to each domain is roughly proportional to the number of GO terms per domain, with almost two orders of magnitude separating the largest domain ‘Proteostasis’ and the smallest ‘Tau Homeostasis’. Many genes are annotated to multiple GO terms subordinate to multiple biological domains. This may represent a convergence of biologically related processes, or the pleiotropy associated with a given gene’s function. While a plurality of annotated genes (30%) are only annotated to a single biological domain, many participate in multiple domains each (Figure 1C), and genes annotated to more biological domains tend to have higher overall Target Risk Scores (see below) (Pearson r = 0.158, p = 2.8×10^−80^).

### Target Risk Score Overview

The TREAT-AD Target Risk Score (TRS) is a metric designed to rank potential disease involvement of specific genes based on multiple independent lines of evidence to objectify the prioritization of potential targets and disease-linked biology. The TRS has two component elements: genetic risk and multi-omic risk, discussed in more detail in the sections belo. Each component score is derived from a meta-analysis harmonizing multiple datasets. The genetic component of the TRS is weighted more heavily, receiving up to 3 points, while multi-omic risk has a maximum of 2 points. The rationale for providing more weight to the genetics score component reflects the greater success in clinical trials for targets with genetic support^37-39^.

### Genetic Risk Score Component

The genetic risk score component is a summary of genetic evidence supporting the target gene’s association with late-onset Alzheimer’s disease from multiple genetic studies. The score is based on genetic evidence retrieved from both genome wide association (GWAS) and GWAS by proxy (GWAX) along with quantitative trait locus (QTL) studies. In total, 27 different Alzheimer’s association data sources were queried (Supp Table 3). These datasets are not fully independent, as the patient cohorts used partially overlap between studies, and as such many genes are scored with associations across multiple studies (Supp Fig 1A). Given the differences between the populations sampled and methodologies applied, we chose a windowed approach to assign variants to genes rather than accounting for the various linkage disequilibrium blocks in the source data. Furthermore, as the goal is to score as many targets as possible for genetic association with AD, we selected a permissive significance threshold for variant inclusion in order to stratify targets that are nominally significant in association with AD but do not meet a more standard criteria for genome-wide significance (e.g. p < 5×10^−8^). Had we restricted our analyses to genes that meet this threshold, only 1,706 of the 60,664 assessed genes (2.81%) would be included (Supp Fig 1B). This would have omitted from consideration the vast majority of genes, including many genes that have notable association with AD but have not yet reached genome-wide significance for various reasons, such as a lack of statistical power and/or the limited diversity in cohort ancestry. Performing gene set enrichment analyses (GSEA) using the combined GWAS score to rank genes (Supp Fig 1C) enriches GO terms from the Immune Response, Synapse, and Lipid Metabolism biological domains (Supp Fig 1D).

We analyzed genes with identified eQTL or pQTL from studies where genetic variation is associated with expression differences in postmortem AD brains. In contrast with the GWA studies, only 492 genes are identified with a significant QTL in all three study populations used (Supp Fig 1E-G), which is consistent with previous reports of limited overlap between brain eQTL and pQTL^40^. Accordingly, GSEA using the integrated QTL signal across the studies to rank genes enriches GO terms from only 5 biological domains - namely Cell Cycle, Proteostasis, Oxidative Stress, Mitochondrial Metabolism, and Immune Response (Supp Fig 1H).

In addition to accounting for the association of a gene with AD traits, we also sought to assess the predicted severity of identified variants. We collected 7,198,607 unique variants with at least a nominal association to AD or related phenotypes, including 50,833 variants predicted to affect the coding sequence of a target gene (Supp Fig 2A) and 1,365,759 variants associated with altered gene or protein expression (eQTL or pQTL) in postmortem AD brain (Supp Fig 2D). Variant severity is scored differently for coding and noncoding variants; while the vast majority of identified variants are noncoding (99.29%), the prediction of functional effects of variation is more straightforward for coding variants. Coding variant severity was assessed for 17,892 distinct genes, 13,088 of which (73.2%) are identified with at least one deleterious coding variant. In general, genes with larger CDS lengths tend to have a higher number of deleterious coding variants (Supp Fig 2B). In addition, there are 939 genes that are among the least tolerant of loss-of-function variation (bottom 10% gnomAD LOEUF) where we identify at least one deleterious coding variant associated with AD. The coding variant summary is enriched for genes in Structural Stabilization, Proteostasis, Immune Response, Endolysosome, Synapse, Epigenetic, and Mitochondrial Metabolism biological domains (Supp Fig 2C). Noncoding variant severity was assessed for variants identified through QTL studies (i.e. those associated with altered expression of the target gene or protein) for a total of 15,812 distinct genes. The noncoding variant summary score integrates information from the RegulomeDB probability score and DeepSEA mean -log e-value (Supp Fig 2E). The noncoding variant summary is enriched for terms from the Immune Response and Proteostasis biological domains (Supp Fig 2F). For the 7,394 genes assessed for both coding and noncoding variant severity, there is a very weak positive correlation between the coding and noncoding variant summary scores (Pearson r = 0.05, p = 1.19×10^−5^, Supp Fig 2G).

**Fig. 2.**
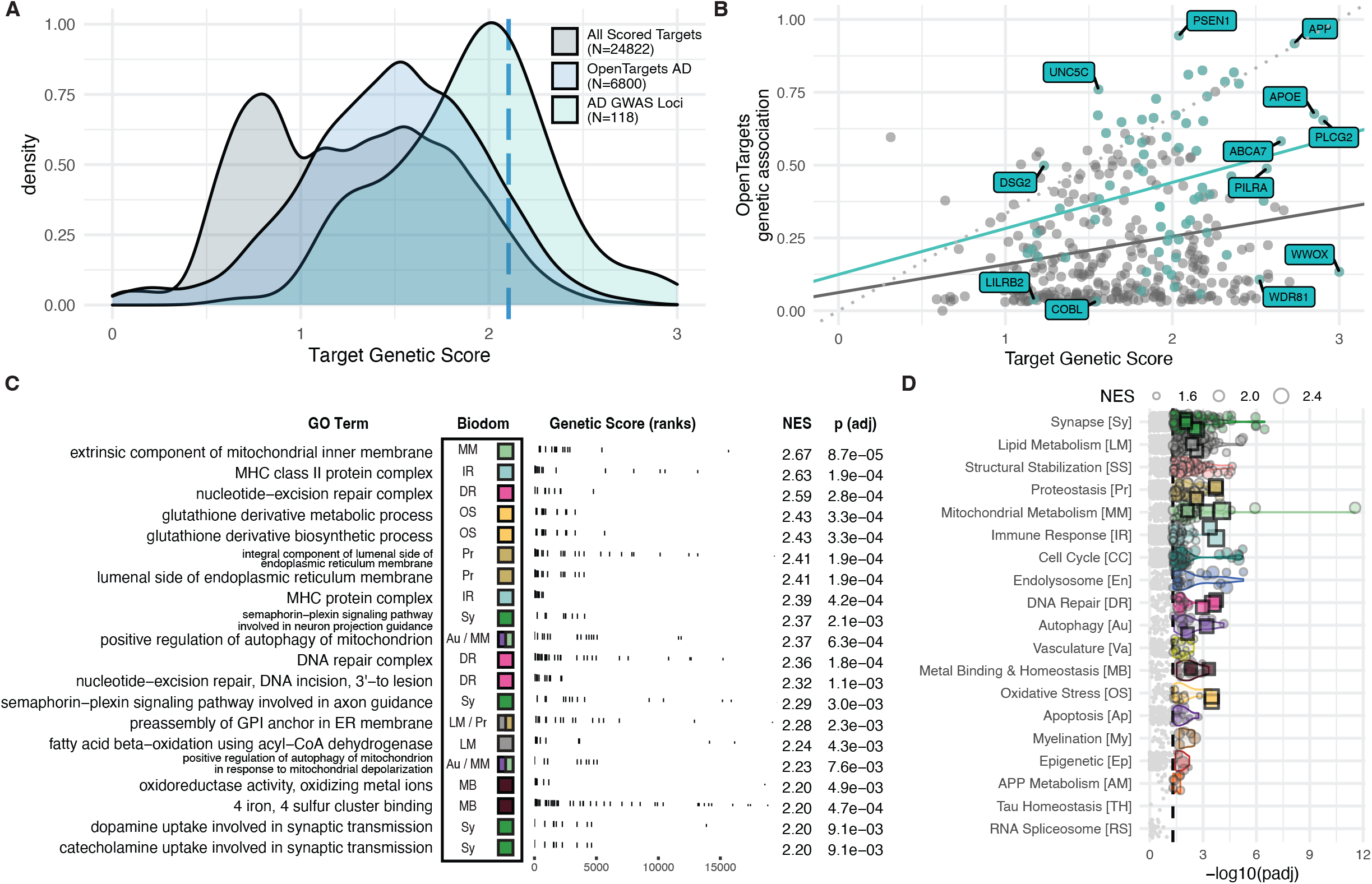
The genetic risk score component. (A) The distribution of all genetic scores (gray), the subset of targets evaluated by the Open Targets platform (blue), and the set of defined AD GWAS loci derived from various sources (green). Dashed blue line indicates the 95th percentile score. (B) Comparison of genetic risk based scores from the TREAT-AD TRS and the Open Targets genetic association score. Plotted in green are the set of all genes within known AD GWAS loci scored by both metrics, several of which are labeled. The dashed line represents an equivalent score on each metric. (C) Top GO terms significantly enriched using the TREAT-AD target genetics score, arranged by normalized enrichment score (NES). For each GO term, the associated biological domain(s) are indicated by the filled square and abbreviations (biological domain colors and abbreviations can be referenced from panel D). (D) Enrichment statistics for all biological domain terms. Each point is a GO term within the indicated biological domain and the size of the point is scaled by the GSEA normalized enrichment score (NES). The GO terms identified in panel (C) are indicated as square points with bold borders. The biological domains are ordered on the y-axis by the number of significantly enriched GO terms identified.

The score also incorporates phenotypic evidence supporting a given target’s potential to impact AD relevant phenotypes from both human and model organism sources. The human phenotype score leverages the Human Phenotype Ontology to query the phenotypic abnormalities annotated to each gene. 4,131 genes have at least one phenotype in common with AD (MONDO:0004975) or dementia (MONDO:0001627) (Supp Fig 3A), and these are enriched for terms annotated to 15 biological domains, but predominantly to the Synapse, Mitochondrial Metabolism, Lipid Metabolism, and Proteostasis biological domains (Supp Fig 3B). For relevant ortholog phenotypes we queried the uPheno ontology^41^ and scored 8,901 genes with orthologs that have at least one phenotype in common with AD or dementia (Supp Fig 3C). Genes with a strong ortholog phenotype score are enriched in GO terms from 17 biological domains, with Synapse, Immune Response, and Lipid Metabolism being the three with the largest number of enriched GO terms (Supp Fig 3D). For the 3,063 genes with relevant phenotypes detected in both human and model organism resources there is a relatively weak positive correlation in phenotype scores (pearson r = 0.20, p = 6.7×10^−30^, Supp Fig 3E). We provide an additional weight to genes that have been positively identified within MODEL-AD studies to be associated with LOAD^42^. MODEL-AD is an NIA funded effort to develop new animal models that better represent features of human AD pathophysiology based on signatures from AD GWAS. We include in our score whether genes have a model in development by MODEL-AD as well as the maximum correlation between human transcriptomic module gene expression from AMP-AD and mouse model gene expression, where available.

**Fig. 3.**
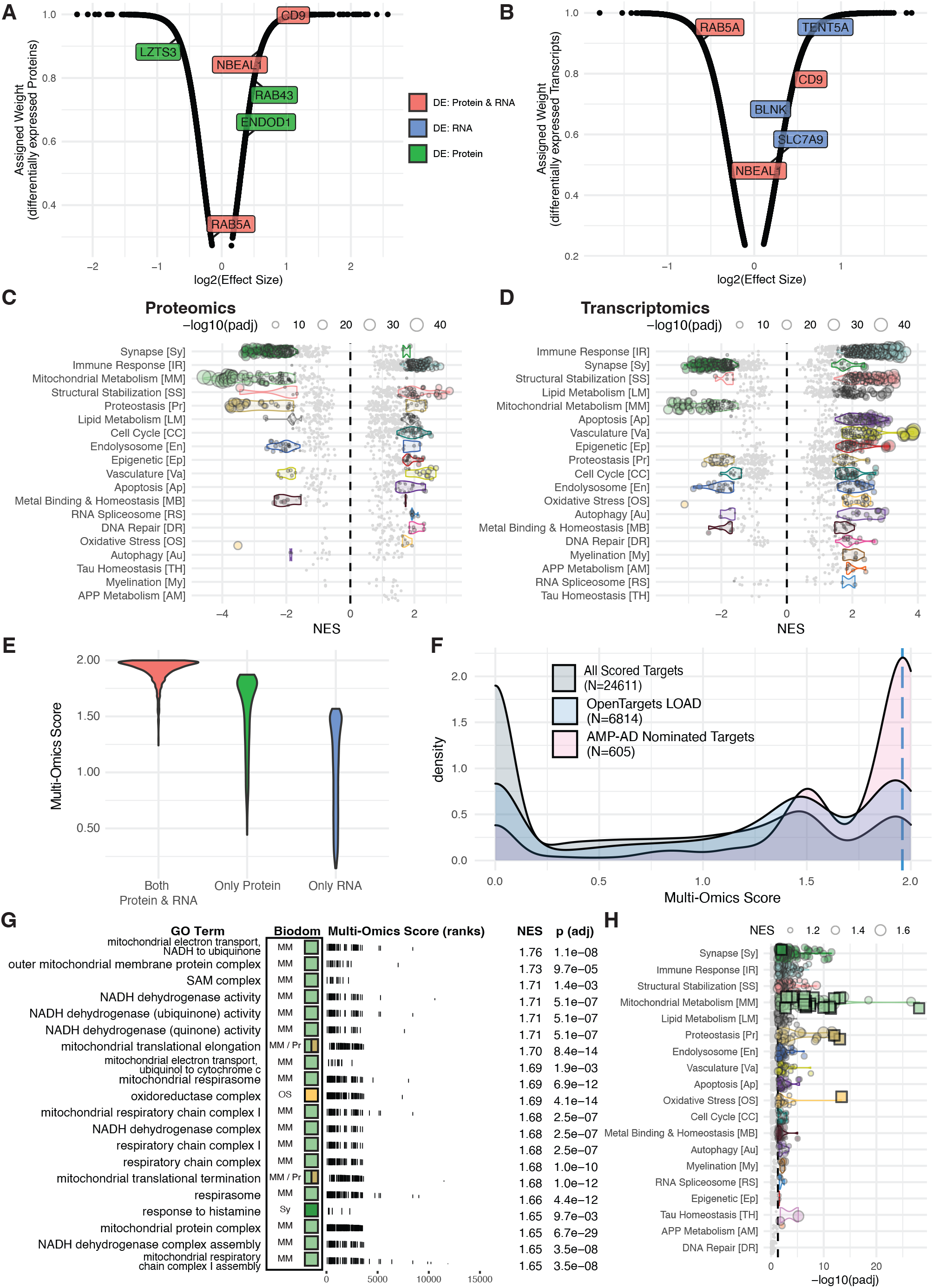
The multi-omic risk score component. Assigned weights as a function of log(effect size) of significantly differentially expressed (A) transcripts and (B) proteins. Labeled points in A-B represent example genes where the color indicates significantly differential expression in only RNA-seq (blue), only proteomics (green), or both RNA-Seq and proteomics (red). (C) Enrichment statistics for all biological domain GO terms based on proteomics meta-analysis treatment effect. Each point is a GO term within the indicated biological domain, the size of the point is scaled by the GSEA adjusted p-value, and the NES indicates if the GO term is up- or down-regulated. The biological domains are ordered on the y-axis by the number of significantly enriched GO terms identified from each domain. (D) Enrichment statistics for all biological domain GO terms based on transcriptomics meta-analysis treatment effect. Each point is a GO term within the indicated biological domain, the size of the point is scaled by the GSEA adjusted p-value, and the NES indicates if the GO term is up- or down-regulated. The biological domains are ordered on the y-axis by the number of significantly enriched GO terms identified from each domain. (E) Assigned weights as a function of log(effect size) of significantly differentially expressed combined weight values as a function of log10 (harness adjusted scaled values), scaled to 2. (F) The distribution of multi-omic risk scores for all scored genes (grey), those scored by the Open Targets platform (blue), and scored genes that are also nominated by a team from the AMP-AD consortium (pink). Dashed blue line indicates the 95th percentile score. (G) Top GO terms significantly enriched using the TREAT-AD target multi-omic risk score, arranged by normalized enrichment score (NES). For each GO term, the associated biological domain(s) are indicated by the filled square and abbreviations (biological domain colors and abbreviations can be referenced from panel H). (H) Enrichment statistics for all biological domain terms. Each point is a GO term within the indicated biological domain and the size of the point is scaled by the GSEA normalized enrichment score (NES). The GO terms identified in panel (G) are indicated as square points with bold borders. The biological domains are ordered on the y-axis by the number of significantly enriched GO terms identified.

The genetic risk score was calculated for a total of 60,664 genes. After tallying all genetic evidence sources we found support for 52,375 genes, though the vast majority of these have only weak or sparse evidence connecting the locus to Alzheimer’s risk. In an effort to limit spurious associations, the list was reduced to only the top 25 percent of ranked genes (15,166 genes) or those scored in the multi-omic analysis (see below). This resulted in a final list of 24,278 scored genes (Supp Table 4). The genes re-introduced to genetic scoring based on their inclusion in the multi-omic analyses are represented as the lower mode of the distribution (Fig 2A, between 0-1). Genes contained within known AD GWAS loci are enriched among the top scores (Fig 2A). Genes identified as high in AD risk by Open Targets (https://www.opentargets.org/)^43-46^, a large-scale effort to rank genes based on genetic support for translational relevance, were also among the top scoring genes (Fig 2A). Comparing the TREAT-AD genetics score with the Open Targets genetic association score for AD (Fig 2B) reveals a very weak positive correlation (Pearson r = 0.185, p = 4.4×10^−4^) which is stronger when only considering known AD GWAS genes (Pearson r = 0.321, p = 6.7×10^−3^). In general, most genes receive a relatively higher score from the TREAT-AD genetics score. GSEA using the genetics score enriches GO terms from 17 of 19 biological domains; the biological domains with the largest number of enriched GO terms are Synapse, Lipid Metabolism, and Structural Stabilization (Fig 2C-D, Supp Table 5). The Open Targets genetic association score enriches GO terms from 10 biological domains, with terms in the APP Metabolism domain by far the most significantly enriched and Synapse, Immune Response, and Lipid Metabolism being the domains with most terms enriched (Supp Fig 4C). The relative emphasis of APP Metabolism in the Open Targets score likely reflects the inclusion of evidence from early onset autosomal dominant forms of AD in which causal variants are clustered in the Presenilin genes (PSEN1, PSEN2) involved in proteolytic processing of APP, whereas the TREAT-AD genetics score draws primarily from genetic associations of late onset sporadic forms of the disease. Notably Synapse, Lipid Metabolism, Structural Stabilization, and Immune Response are among the biological domains with the largest number of enriched GO terms for both scores.

**Fig. 4.**
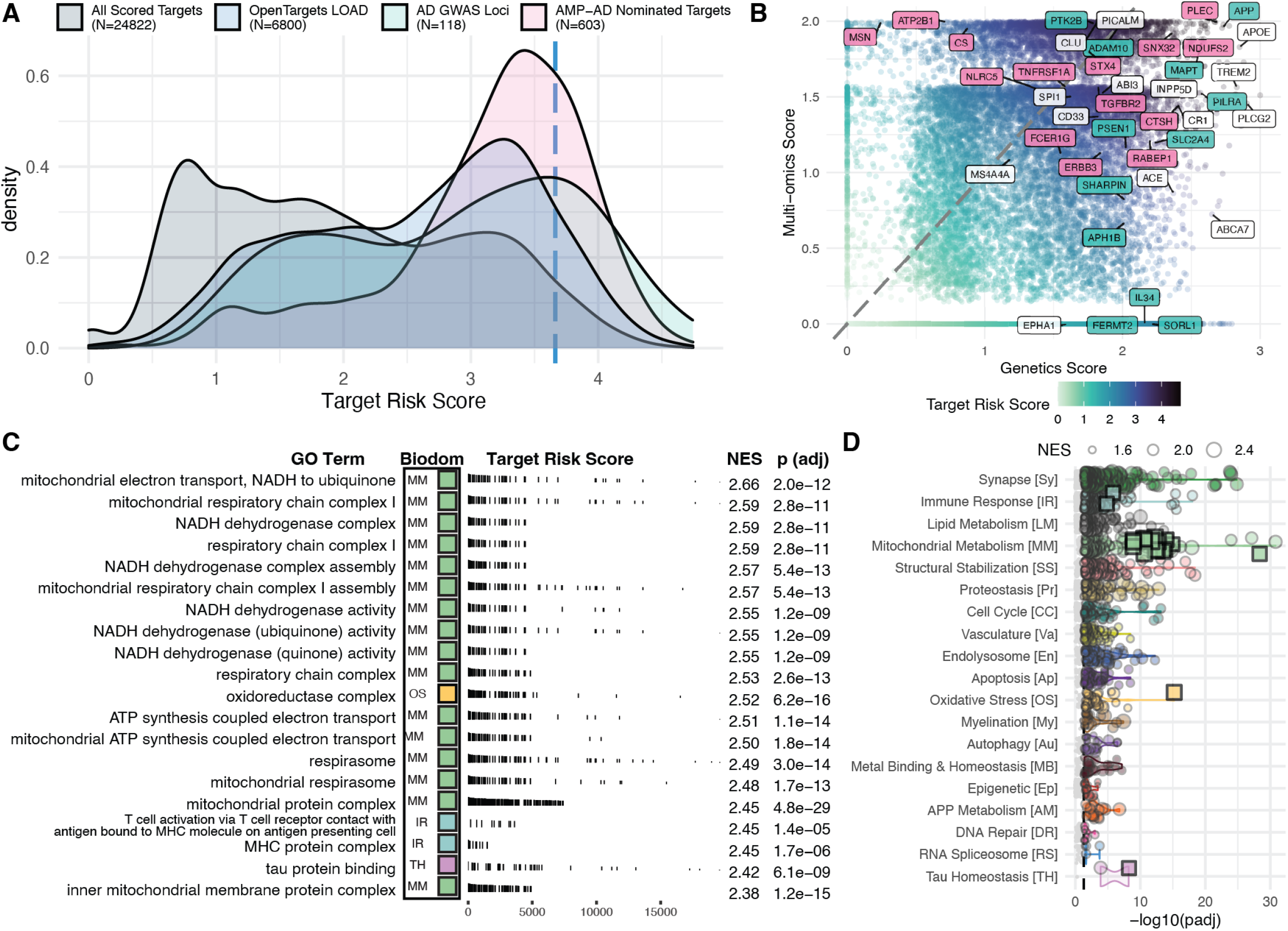
The composite TREAT-AD Target Risk Score (TRS). (A) The distribution of TRS for all scored targets (gray), those scored by the Open Targets platform (blue), those nominated by a team from the AMP-AD consortium (pink), and the set of defined AD GWAS loci derived from various sources (green). Dashed blue line indicates the 95th percentile score. (B) Comparison of the genetics score and the multi-omic score for all targets and point color indicates the composite TRS for the target. The genes that are labeled represent a subset of either GWAS genes (green), AMP-AD nominated genes (pink), or both (white). (C) Top GO terms significantly enriched using the TRS, arranged by normalized enrichment (NES). For each term, the resident biological domains are indicated by the filled square (biological domain colors can be referenced from panel D). (D) Enrichment statistics for all biological domain GO terms. Each point is a GO term within the indicated biological domain and the size of the point is scaled by the GSEA normalized enrichment score (NES). The terms identified in panel (C) are indicated as square points with bold borders. The biological domain terms are ordered on the y-axis by the number of significantly enriched terms identified from each domain.

### Multi-omic Risk Score Component

The multi-omic risk score is a summary metric encapsulating available evidence supporting that target gene expression is altered in the brains of AD patients. The score makes use of proteomic and transcriptomic datasets generated as part of the consortium efforts of AMP-AD (Supp Tables 6 and 7). For each data modality (i.e. transcriptomic or proteomic), a meta-analysis of samples is used to generate weights for significantly differentially expressed genes based on the observed fold changes (Fig 3A & 3B). Using ratio of means meta-analysis with a random effects model for each data modality, we analyzed the directionality of expression change due to AD using GSEA (Fig 3C & 3D). This analysis demonstrates that Synapse and Mitochondrial Metabolism are the biological domains with the largest number of down-regulated GO terms and Immune Response and Structural Stabilization are the biological domains with the largest number of up-regulated GO terms – across both proteomics (Fig 3C) and transcriptomics (Fig 3D).

The calculated weights for each modality are combined using a scoring harness (Supp Table 8) that yields a higher score for targets with (A) evidence of differential expression at both the protein level and the RNA level, followed by (B) those targets that are only significantly differentially expressed at the protein level, followed lastly by (C) those that are only significantly differentially expressed at the RNA level (Fig 3E). The rationale for this harness is two-fold: first, concordant evidence from multiple data modalities leads to higher confidence that a target gene’s expression is altered in AD brains, and second, that protein levels more accurately relate to the state of the biology within an in vivo system^35,47,48^. Importantly the effect of the harness is tuned to prioritize this imposed hierarchy without disregarding results specific to a single profiling modality. This reflects a balance between higher relevance of proteomic evidence to disease state versus the increased sensitivity of detection for transcriptomics. We compare the distribution of all scored targets (Supp Table 9) to those scored by the Open Targets Platform^43-46^ and those nominated for follow-up by the AMP-AD consortium^49,50^. The 605 targets nominated by AMP-AD investigators tend to have higher multi-omic scores than the population of all targets (Fig 3F), and there is a very weak correlation between the multi-omic score and the number of nominations received by a given target (r = 0.153; Supp Fig 5E). Numerous targets receive only one nomination, yet are ranked among the highest multi-omic scores. Conversely, there are many targets that received several nominations with a low multi-omic score. This likely reflects the fact that AMP-AD investigators use diverse methods to identify targets, beyond differential expression analysis, and that some modalities (e.g. metabolomics) are not yet included within the multi-omic score. Future efforts will include work to integrate additional data modalities into the score. Using the multi-omic score to rank genes to perform GSEA results in the enrichment of GO terms from 18 biological domains, with DNA Repair the only domain with no terms enriched. The biological domains with the largest number of enriched terms include Synapse, Immune Response, Structural Stabilization, and Mitochondrial Metabolism (Fig 3G-H, Supp Table 10). Terms from the Mitochondrial Metabolism domain are both among the most significantly enriched (Fig 3H) and have the highest normalized enrichment score (NES) (Fig 3G). Interestingly, the top terms within Mitochondrial Metabolism focus upon mitochondrial translation and complex I of the electron transport chain (Fig 3G), showing these terms are the most significantly down-regulated biological processes associated with LOAD.

**Fig. 5.**
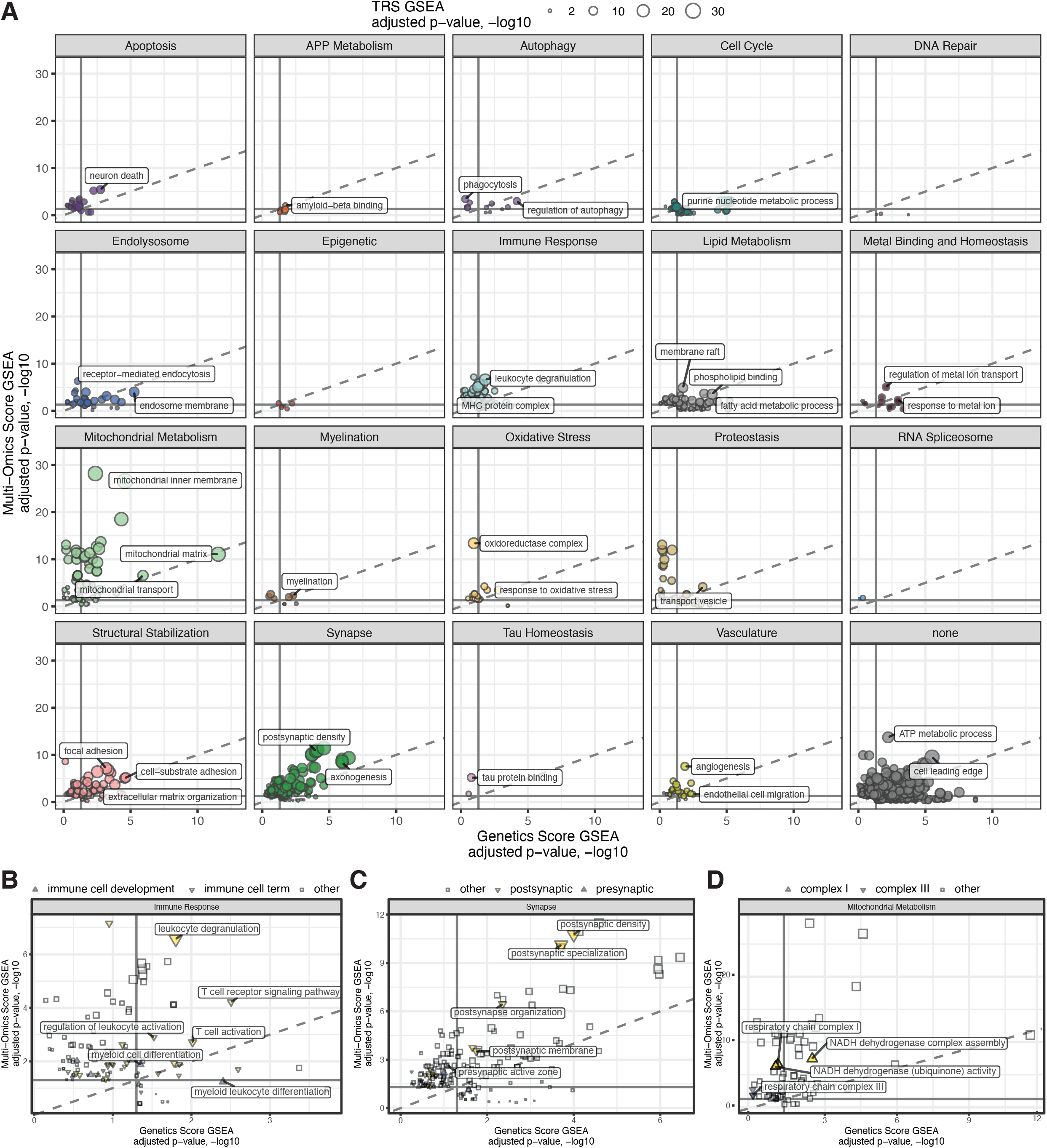
Comparing biological domain enrichments across risk modalities. (A) Relative significance of biological domain term enrichment using the genetics score component (x-axis), the multi-omic score component (y-axis). and the overall TRS (point size). (B) Term enrichments from the Immune Response biological domain, subset by immune cell differentiation terms (blue), other immune cell terms (yellow), and other terms from the Immune Response biological domain (grey). (C) Term enrichments from the Synapse biological domain, subset by postsynaptic terms (yellow), presynaptic terms (blue), and other terms from the Synapse biological domain (grey). (D) Term enrichments from the Mitochondrial Metabolism biological domain, subset by mitochondrial electron transport chain complex I (yellow), complex III (blue), and other terms from the Mitochondrial Metabolism biological domain (grey). Complexes II, IV, and V were not significantly enriched in this analysis.

### Composite Target Risk Score

The TRS is a metric derived by summing the component risk scores (i.e. genetic and multi-omic scores). The maximum observed score for any target is 4.74 out of a total of 5 (Supp Table 11). As with the component scores, the top TRS scores are enriched for GWAS loci, AMP-AD nominated targets, and targets considered by the Open Targets platform (Fig 4A). Considering both the genetic and multi-omic scores for each target (Fig 4B), the targets with the top TRS tend to have relatively higher genetic scores. When we compare the TREAT-AD overall TRS with the Open Targets target score (Supp Fig 4A) we see that many targets receive a relatively higher score from the TRS, likely due to the unique inclusion of disease-relevant transcriptomic and proteomic datasets from the AMP-AD consortium in the TRS. AD GWAS loci are generally scored highly by both the TREAT-AD TRS and the OpenTargets target score metrics. GSEA using the overall TRS enriches 3,142 GO terms, including 1,358 (43.5%) annotated to at least one of each of the 19 biological domains (Supp Table 12). The biological domains with the largest number of enriched GO terms are Synapse, Immune Response, and Lipid Metabolism (Fig 4D). In comparison, the Open Targets target score enriches terms from 16 of 19 biological domains, and the domains with the largest number of enriched terms are also Synapse, Lipid Metabolism, and Immune Response (Supp Fig 4B).

Considering all biological domain GO terms that are significantly enriched with the TRS, some have more significant enrichment from GSEA using the genetics score versus the multi-omic score to rank genes (Fig 5A, Supp Tables 5, 10, and 12). For example, from the Lipid Metabolism biological domain the “fatty acid metabolic process” term is enriched using both scores, but for the genetics score based tests the adjusted p-value for the term is 5.9×10^−6^ while for the multi-omic score based test the adjusted p-value is 7.9×10^−3^. The inverse is true for the “membrane raft” term which is more significant using the multi-omic scores (adjusted p = 1.3×10^−5^) compared to the genetics scores (adjusted p = 1.8×10^−2^). A similar dichotomy is observed for terms associated with developmental processes from the Immune Response domain, especially “myeloid leukocyte differentiation”, having relatively more significant enrichment from the genetics score whereas corresponding terms for mature structures and associated processes (e.g. “leukocyte degranulation”) have a relatively more significant enrichment from the multi-omic score (Fig 5B). Other biological domain resident dichotomies are more consistent across risk modalities. For example, considering the all presynaptic and postsynaptic terms from the Synapse biological domain, there are more postsynaptic terms enriched (40 terms) relative to presynaptic terms (9 terms) and the postsynaptic terms are enriched with more significance across the TRS as well as the genetic and multi-omic score components (Fig 5C). The optimal targets reflect a parity of ranking between the two contributing modalities, with accumulation of both genetic and multi-omic risk. For example, enrichment of genetic risk without multi-omic risk can indicate risk in developmental processes that are not present in later stages or disease or postmortem tissue, whereas enrichment of multi-omic risk without genetic risk can indicate processes that are changing as a response to disease pathology but are not causal to underlying disease etiology. Moreover, processes that are risk enriched across two distinct measures increases our confidence of disease association. For example, while the multiomic score enriches terms related to mitochondrial electron transport chain complexes I and III, the genetics score only enriches terms related to complex I (Fig 5D), yet both measures point to the centrality of electron transport chain related events in our scored AD risk. GO terms close to the diagonal dashed line (Fig 5A), as seen in Structural Stabilization with the term “cell-substrate adhesion”, suggest that the risk associated with genes in these terms are embedded equivalently in each data modality and therefore are worth consideration for further resource development. The GO terms that are significantly enriched but do not associate with any biological domain (i.e. Fig 5A, ‘none’) seem to reflect either biological process categorization that is too general to be mapped into endophenotypic space, such as “ATP Metabolic Process”, or terms that map to high order positions, or do not imply any specific associated biological process, in the molecular function or cell component ontologies within GO, such as “Cell Leading Edge” (Figure 5A) also rendering them uncategorizable in endophenotypic space.

### Example Use Case: AMP-AD Co-expression Modules

To demonstrate how this information can be useful more broadly, we apply these approaches to published co-expression modules for transcripts^20,22^ and proteins^35^ produced by AMP-AD consortia members. For each module we calculated the median TRS and genetics and multi-omic component scores (Supp Table 13), performed GO term enrichment analyses using the identities of the genes in each module, and categorized enriched GO terms into biological domains. The STGblue module has both the highest median TRS and the highest median multi-omic scores of the 30 transcript co-expression modules from Wan *et al*, while the DLPFCblue module has the highest median genetics score (Fig 6A). Among the 45 protein co-expression modules from Johnson *et al*, the M11 Cell-ECM Interaction module has the highest median TRS and the highest median multi-omic score while M27 Extracellular Matrix has the highest median genetics score (Fig 6B). The GO term enrichments for the three proteomics modules with the top median TRS are primarily from the Structural Stabilization (Fig 6C & 6D) and Mitochondrial Metabolism (Fig 6E) biological domains. Comparing the GO term enrichments for the modules from each study with the highest median genetics scores and highest median multi-omic scores, we note several common enrichments across modules from all three studies. The GO terms enriched for proteins in modules with the highest median multi-omic (M11, Supp Fig 6B) and highest median genetics scores (M27, Supp Fig 6C) are predominantly from the Structural Stabilization biological domain. This is also similar for the genes in the transcriptomic sub-modules from Milind *et al*, where the enriched GO terms from the sub-module with the highest median multi-omic score (IFGturquoise_2, Supp Fig 6F) are primarily from the Structural Stabilization biological domain. The transcriptomic sub-module with the highest median genetics score (PHGturquoise_2, Supp Fig 6E) is enriched for many terms from the Immune Response biological domain. The biological domains enriched for each of the top-scoring transcriptomic modules from Wan *et al* are very similar - primarily Immune Response, Structural Stabilization, Vasculature, and Lipid Metabolism, in varying orders (Supp Fig 6H and I). In Wan *et al* these modules were associated with microglial, pericyte and endothelial cell types relevant to these biological domains. The large number of biological domains enriched is likely due to the fact that these modules are large (between 504 and 4,673 genes), compared with the proteomic modules (between 28 and 2,231 proteins) and the transcriptomic sub-modules (between 77 and 2,090 genes), so more terms are enriched for each module and the overall pattern of enrichment is less specific. The top risk-enriched co-expression modules from each study implicate terms from the Structural Stabilization and Immune Response biological domains. Moreover there are GO terms that are shared among these co-expression modules, including “focal adhesion” and “extracellular matrix” for the Structural Stabilization biological domain and “neutrophil degranulation” and “cytokine-mediated signaling pathway” for the Immune Response biological domain.

**Fig. 6.**
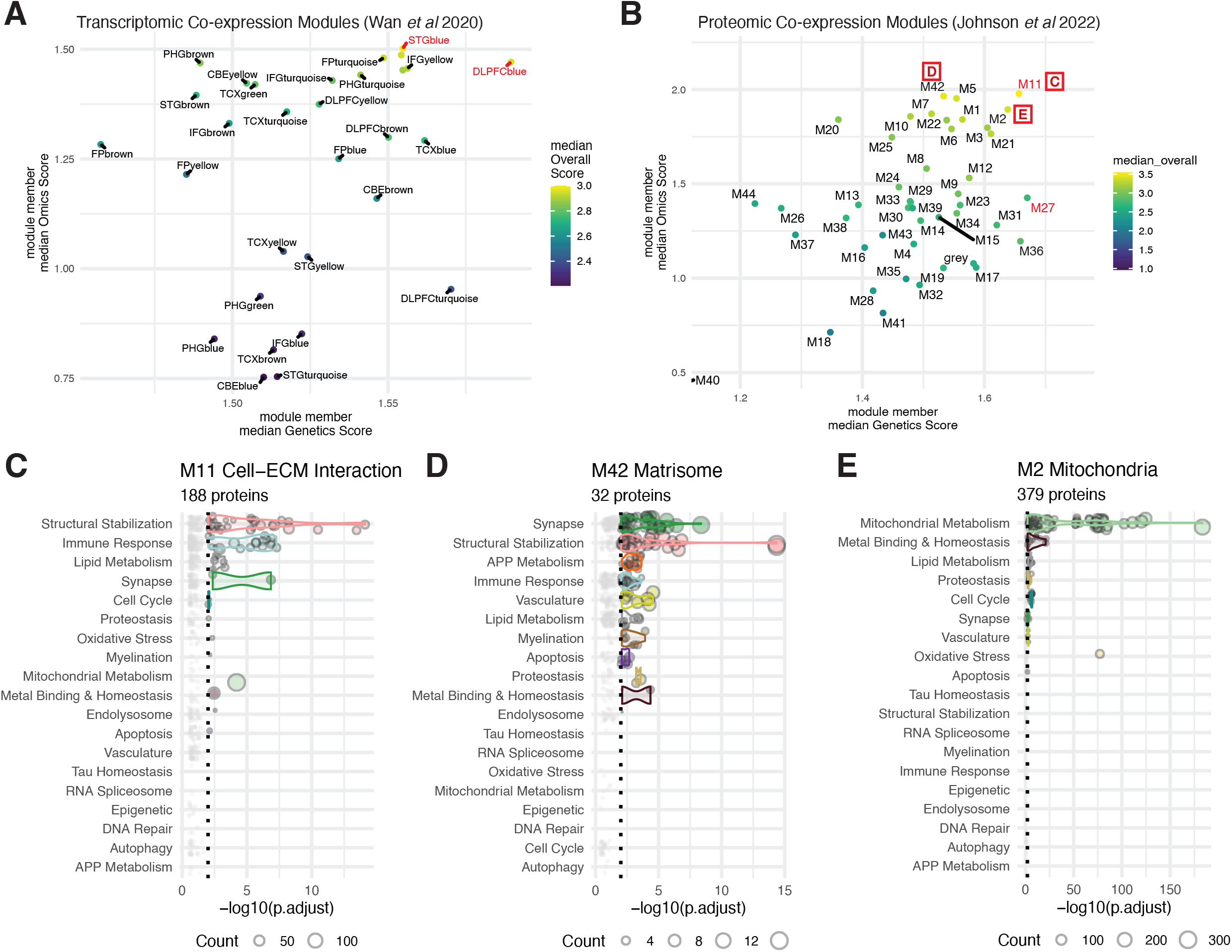
Top scoring AMP-AD co-expression modules and biological domain enrichments. (A) Median genetics scores (x-axis), median multi-omic scores (y-axis), and overall scores (color) are plotted for each transcriptomic co-expression module from Wan *et al*. Top scoring modules on each dimension are highlighted in red; STGblue has the highest median overall TRS and multi-omic score and DLPFCblue has the highest median genetics score. (B) Median genetics scores (x-axis), median multi-omic scores (y-axis), and overall scores (color) are plotted for each proteomic co-expression module from Johnson *et al*. Top scoring modules on each dimension are highlighted in red; M11 has the highest median overall TRS and highest median multi-omic score, while M27 has the highest median genetics score. The modules with top three median TRS, for which GO term enrichments are plotted, are indicated with red letters corresponding to the figure panel. (C-E) Biological domain GO term enrichment statistics for module member proteins. Each point is a term within the indicated biological domain and the size of the point is scaled by the number of proteins in that term that were included in the module. The name and size of each module is indicated above each plot.

## Discussion

The work presented here represents the most comprehensive effort to date to employ an unbiased computational system of AD risk assessment in coordination with an objective, systematic and comprehensive alignment with AD endophenotypes. Broadly, we have executed this task in two parts. First, we have developed an AD risk scoring paradigm that draws from multiple genetic association studies as well as large multi-omic datasets consisting of 1,699 brains with transcriptomic data (Supp Table 6) and 1,188 brains with proteomic data (Supp Table 7), each drawn from multiple large brain bank studies, to objectively rank genes genome wide. Second, we have developed 19 biological domains that align with known AD endophenotypes, defining each with a specific and exhaustive set of GO terms and annotated gene sets (Supp Table 2). This enables the organization of risk associated genes into objective and unbiased biological processes, to facilitate the characterization of risk enriched areas for therapeutic development. The goal is to provide the scientific community with an evaluative resource that can be employed across studies and modalities to help integrate knowledge about potential domains and targets for future investigation as well as provide a unified framework for defining exactly what is meant by an AD endophenotype. The biological domains described in this work have already been adopted by several groups - including the AD Knowledge Portal^14^ (https://adknowledgeportal.synapse.org), the AD Atlas^51^ (https://adatlas.helmholtz-muenchen.de), Agora (agora.adknowledgeportal.org), the AMP-AD Consortium, and the TREAT-AD Consortium and others - as a common organizational reference tool. The expanding utilization of these biological domain endophenotypic definitions will provide the opportunity for increased interoperability and harmonization across large platform research efforts. Both methodologies - risk scoring and biological domain annotation - are easily updated as new information becomes available, as they draw from open-source communal resources central to AD biological investigation.

The quantitative scoring of genetic and multi-omic components of the TRS enables us to perform enrichment tests on both the composite score as well as the individual score components. In this way we can interrogate genes across the biological domains for cumulative risk, and also assess whether the risk comes primarily from one data modality. These analyses (Fig 2-4) highlight the observation that the different modalities emphasize independent biological areas of disease risk, but employed in unison are a powerful tool for mapping disease risk into specific biological areas within objectively defined disease-associated endophenotypes (e.g. Fig 5A, Supp Tables 5, 10, and 12).

Using the biological domain framework, we investigated which biological domains were the most up- and down-regulated based on post-mortem differential transcriptomic and proteomic abundance analyses. The top two down-regulated areas of AD-linked biology are Mitochondrial Metabolism and Synapse (Fig 3C and 3D), which are also the biological domains with the most significantly enriched terms in both the genetics and multi-omic score as well as the overall TRS. The down-regulation of synaptic genes span both pre- and postsynaptic gene sets, however there are 3-4 times as many postsynaptic GO terms enriched and postsynaptic terms are more significantly enriched across all component risk scores (Fig 5C). This aligns with previous research into the role postsynaptic mechanisms play in the maintenance of dendrites and synaptic plasticity, lost during the cognitive decline of AD ^52-54^. Numerous studies have demonstrated elements of mitochondrial dysfunction and hypometabolism in AD, corroborating the data-driven narrative emerging from these studies^1,55-57^. Furthermore, these two domains of AD-linked biology are associated with cognitive stability in aging^58^, and may be coordinately progressive in AD^56,57,59^. One of the long-term objectives of our approach is to be able to detect frameworks of convergent biology involved in disease progression. While the analysis presented here only points to the independent identification of mitochondrial hypometabolism and synaptic dysfunction or loss, future work employing a broader set of analytical approaches may be able to use risk enrichment in portions of the biological domains to highlight aspects of interacting biology at the intersection of these domains.

There are multiple up-regulated biological domains demonstrated within our analysis, chief among them are Immune Response, Structural Stabilization, Lipid Metabolism and Proteostasis (Figure 3C and 3D). The investigation of microglial activation in AD pathological progression has received increased attention recently, with respect to both janus faces of microglia: the homeostatic and degenerative roles. A meaningful discussion of these areas is beyond the scope of this work, but the topic has been extensively reviewed in recent years^60-64^. Similarly, structural stabilization is observed in many recent brain proteomic studies, suggesting a compelling role for heparin binding proteins, extracellular factors, and cytoskeleton associated proteins in either resilience or progression of AD^35,65^. The field of metabolomics focuses extensively upon dysfunction in lipid metabolism in AD, and is mapped out within the AD Atlas (adatlas.helmholtzmuenchen.de) of the AD Metabolomics Consortium^66-68^. Proteostasis is highly implicated in AD as well, suggesting complex linkages between autophagy and proteome stability, changes in chaperone mediated protein processing and ER stress responses may all play a role in AD^69-72^. The identification of sets of biological domains that are up-regulated and align with AD risk provides defined areas of biology to explore further for both disease driving mechanisms and compensatory processes that may facilitate future therapeutic development.

In order to assess how our ranking system compares to resources that perform a similar function, we benchmarked our scoring process with corresponding rankings from the Open Targets platform^43-45,73^. The two methodologies share several features in common; both approaches integrate results from genetic association studies to implicate genes with variation that contributes to disease risk, both capture expression differences relevant to the disease, and both identify animal models with phenotypic relevance to disease. These similarities are reflected in the correlation observed between the two scores (e.g. Supp Fig 4A). The TRS is distinct in a number of important ways. The TRS includes both transcriptomic and proteomic datasets derived from multiple brain tissues from numerous brain banks via our partners in the AMP-AD Consortium. Similarly, information pertaining to relevant AD mouse models from the MODEL-AD centers is included. We benchmarked the scoring output of Open Targets and the TRS and observed very similar patterns of enrichment (e.g. Supp Fig 4B), with the Synapse, Lipid Metabolism, and Immune Response biological domains among the strongest enrichments for each score. However, the enrichments using the TRS implicate Mitochondrial Metabolism (120 enriched terms) and Tau Homeostasis (2 enriched terms) more strongly than the enrichments based on the Open Targets score (1 enriched term and 0 enriched terms, respectively).

Finally, we used the TRS and biological domain framework to assess sets of co-expression modules produced from large scale genomic AD investigations. We found highly similar biological domain enrichments for the top scoring co-expression modules. The proteins and transcripts from co-expression modules and submodules that implicate extracellular matrix and cell junction biology (M11, STGblue, and IFGturquoise_2) are enriched for terms from the Structural Stabilization biological domain, share enrichments in the “focal adhesion” and “extracellular matrix” GO terms, and tend to have high median multi-omic scores relative to other modules. The proteins and transcripts from co-expression modules and submodules predominantly implicating immune function (M21, DLPFCblue, and PHGturqoise_2) are enriched for terms from the Immune Response biological domain, share enrichments in the “neutrophil degranulation”, and “interferon-gamma-mediated signaling pathway”, among others, and tend to have high median genetics scores relative to other modules. Thus, using a common framework to rank and organize the genes and proteins that emerge from these systems-level studies highlights common processes associated with increased risk emerging from multimodal platforms of investigation.

## Conclusion

This work is the largest integrated effort to combine genetic and multi-omic AD risk scoring with an automatable system of endophenotypic genetic characterization. The dual processes of TRS ranking genes across the genome and assembling the risk areas into biological domains points to a consistency between our objective analyses and observations made across the field, which supports our approach to identifying focal areas of AD risk. The advantage of our system is that we can utilize the comprehensive representation of AD risk genes organized in an unbiased fashion into specific areas of biology to expand the study of disease domain transitions, by identifying potential points of convergence between interacting domains, and examining the genetic entities at those cross-roads. This approach is utilized by the TREAT-AD center led by Emory-Sage-SGC to help identify specific dark targets for future exploration as potential therapeutics – the informatics and material resources developed will be made openly available to the AD scientific community. These approaches will continue to be refined and expanded based upon newly emerging data and input from the scientific community. We hope these resources and analytical techniques may help the field foster current efforts leading to growth of novel translational approaches, or the repurposing of therapeutics developed in divergent fields, for use in the treatment of AD. The objective identification of the ranked areas of disease risk scored genome wide and organized into defined biological domains highlight the significance of multiple domains of biology for translational development, that current resources can help hone into specific subdomains–such as mitochondrial complex I related factors and postsynaptic targets, as well as up-regulated targets in the Immune Response and Structural Stabilization biological domains.

## Methods

### Alzheimer’s Disease Biological Domains & Enrichment Analysis

#### Methodological Overview

The development of the biological domains broadly encompasses two distinct processes: the selection and the definition of each biological domain. The selection of the biological domains is guided by the attempt to exhaustively identify the endophenotypes and biological areas linked to AD pathogenesis. As Alzheimer’s disease (AD) is a heterogeneous neurodegenerative pathology with multiple interacting biological events either stemming from, or contributing to, the central disease sequelae, there have been repeated efforts to exhaustively categorize the subpathologies and endophenotypes in AD. One of the most developed resources is the Common Alzheimer’s and Related Dementias Research Ontology (CADRO) developed by the National Institute on Aging in association with the Alzheimer’s Association^5^. As CADRO is already in standard use for drug development classification, we leveraged this resource to help guide the initial stages of identification of relevant biological domains of AD. However, as the focus of the biological domains is upon the identification of subprocesses and pathologies in AD that may cut across cell-types, the inclusion of CADRO terms involved a rearrangement of the structure to facilitate cell-type autonomy of disease processes. For example, autophagy is made an independent biological domain as it does not occur exclusively within immune cells. The identification of biological domains was expanded beyond CADRO to be maximally inclusive of data derived from large scale consortia studies in different areas of disease relevance. The expansion goal is two-fold: first, to be as comprehensive as possible across AD research; and secondly, to align with our own scoring criteria. Consequently, we focused upon the categorization of processes implicated by GWAS studies and large scale multi-omic investigations. We also categorized the AD hypothesis literature to ensure we were not missing any key concepts or fields of study. While this scope lends to an unbounded examination of disease linked biological traits, we attempted to be broad enough in the biological domain definitions to capture large areas of related disease process and to constrain the studies leveraged to those primary publications within each area of the field. The process is detailed below.

#### Genetic Considerations in Selecting Biological Domains

In genetics, we focused on key genome wide association studies (GWAS), the newer genome wide association study by proxy (GWAX)^15-19,74-80^ using the parental disease status, and whole exome sequencing studies^81-95^ that have transpired over the last decade. The identification of potential genetic risk associated with individual genes is represented in the genetics score (detailed below), and the goal here is not to recapitulate the scoring methodology, but to assess the potential biological contexts of the imputed genes’ biological function. The characterization of gene function was completed by examining its functional classification within UniProt and Entrez gene, the linked biological process gene ontology terms, and the description within the primary literature. We examined both those genes that were validated through eQTL or pQTL^40,50^, as well as the lead statistically associated gene, and the gene set analysis results obtained via MAGMA analysis^96^ performed in most of the abovementioned studies. While there are a multitude of caveats to the interpretation of the genetically identified loci, recently reviewed by Goate et al^97^, our goal was to capture potential biological relevance of the genetic observations, providing a biological space for future gene validation results. We acknowledge this approach increases sensitivity of the biologically associated areas at the potential cost of decreases in specificity, but as the goal is to create a broad hypothesis space, we deemed this trade off acceptable at the present time. The genetic investigation recapitulated many of the biological domains identified within CADRO (Supp Table 1) and reclassified within our structure as Immune Response, Endolysosomal Trafficking, APP Metabolism, Tau Homeostasis, Lipid Metabolism, Synapse, and Epigenetics (recently reviewed^97-99^).

#### Genomic Consideration in Selecting the Biological Domains

The genomic branch of our investigation involved the interrogation of the primary large postmortem transcriptomic or proteomic studies performed in the last five years. There are a multitude of brain proteomic studies associated with TREAT-AD or AMP-AD investigations that have identified functional modules of co-expressed genes that associate with various parameters of neurodegenerative pathology^21,30,32,33,35,58,65,100-106^. These modules generally center upon specific biological functions, and hence are amenable to biological domain subordination. One novel biological domain that we created to describe a set of genes involved in binding extracellular matrix, forming cell-cell junctions, or translating interactions from the extracellular milieu to the intracellular cytoskeleton. We named this biological domain Structural Stabilization as these proteins play a role in intracellular and intercellular structure critical to cellular processes, where shape, or the modification of cellular shape, may be essential for biological function. The leading modules in many studies were in alignment with core features called out within the CADRO ontology, which we took as dual validation of the biological significance of these domains: Synapse, Mitochondrial Metabolism, Oxidative Stress, Proteostasis, Immune Response, Endolysosome, APP Metabolism, Vasculature, Lipid Metabolism and Tau Homeostasis. The nomenclature we use varies from that provided by CADRO, but the conceptual overlap is high, with 14 of the 19 biological domains having some instantiation within CADRO (Supp Table 1). Several genomics studies implicated RNA splicing factors as potentially involved in disease state, promoting the development of the RNA Spliceosome biological domain^33,100,107,108^.

#### Literature Consideration in Selecting the Biological Domains

We interrogated the literature for other biological domains through a focused search of Alzheimer’s related hypothesis papers, specifically querying for the co-occurrence of “Alzheimer” and “hypothesis” within the title. The search produced 463 articles, 51 of which were excluded for topics not pertaining to molecular causes of disease, such as lifestyle factors that may protect against disease risk, imaging observations, or isolated speculation about systems level therapeutics. Of the remaining 412 papers, the largest groups were the amyloid hypothesis (n=151), which maps onto the APP Metabolism biological domain, and synaptic (n=90) hypotheses, mapping onto the Synapse biological domain, followed by Immune Response (n=32) related disease mechanisms (Supp Fig 7). Nested within the synaptic hypotheses articles were topics covering the cholinergic hypothesis, as well as other hypotheses focused upon other specific neurotransmitter systems, such as glutamate, serotonin and dopamine. All of the identified biological domains were supported by at least 1 hypothesis paper, however, several hypotheses were identified for which we did not previously have coverage. These included Cell cycle (n=12), DNA Repair (n=6), and Metal Binding and Homeostasis (n=28)–the latter consisted of a set of hypotheses focused predominantly upon iron and copper. Following the identification of significant GO term enrichment within each (see Figures 2-4), we elected to retain these biological domains. The literature identified numerous hypotheses we elected not to include at this time. Those include: the calcium cascade hypotheses (n=15), diabetes hypotheses (n=11) and microbial infection hypotheses (n=26). The calcium cascade hypothesis was not included as it significantly overlaps with the Apoptosis, Mitochondrial Metabolism and Proteostasis biological domains. The diabetes hypothesis was not included as glucose metabolic processes downstream of transport are already modeled within Mitochondrial Metabolism, and we are attempting to keep the biological domains discrete and as siloed as possible. The microbial infection hypothesis was not developed as a biological domain, as the main endogenous genetic signal for this domain is already resident in the host response related terms within the Immune Response biological domain, and we are not examining exogenous genomic information within our analysis framework.

#### Conclusions on the Selection of the Biological Domains

The 19 biological domains identified in this work through the examination of CADRO and the areas discussed above elucidate a set of endophenotypes that have been already noted in the field–as each domain has at least one supporting hypothesis paper associated with it (Supp Fig 7). Each of these biological domains as implemented, the details of which are discussed below, demonstrate strong AD risk signal via our scoring methodology in either genetics, genomics or both. We decided to conclude the development of biological domains with these core domains as analysis of the GSEA enriched GO terms (methodology below) that did not participate in the characterized biological domains appeared to be uncategorizable into disease endophenotypic space for one of several reasons: (1) the term was a high-level but non-specific term (such as ATP Metabolic Process), (2) it involved cellular localization information that also did not relate to specific disease related processes (such as Cell Leading Edge), or (3) related to molecular process information that was either generic or not categorizable within a specific biological domain (such as tyrosine kinase). Consequently, we deemed the current build sufficient for our initial release. We will continue to look for additional disease linked processes that intersect with the current biological endophenotypes, as we observed that the majority of genes belong to multiple domains.

#### Implementation of the Biological Domains

The implementation of the biological domains for drug target identification studies and future biological investigations of AD pathogenic mechanisms required a strategy that was (i) objective, (ii) automatable, (iii) easily intelligible, and (iv) communally modifiable. Based on these criteria, we elected to use an exhaustive elaboration of gene ontology (GO) terms associated with each biological domain as the core definition. A five part development cycle was followed for the instantiation of each domain (Supp Fig 8). The first step was to identify the starting query terms to employ in searching the ontology. For example, for Immune Response, “innate immune” was one of the high-level query terms invoked. The terms linked to key points within the ontology that allowed a local search of parent and child terms—those falling within the conceptual biological space of the biological domain were collected and aggregated as step 2. The third step involved the use of these terms within the EBI GO infrastructure to expand the set of linked terms to identify GO annotation terms that may have been missed from the query-based ontology evaluation. These terms were incorporated into the GO term definition space. The fourth step involved the manual examination of each term within the biological domain definition architecture to ensure that query expansion had not identified spurious terms, or child terms that were the junction of two parents from inside and outside the biological domain space that no longer were consistent with the biological domain. This took extensive and iterative rounds of review. Once the domains were adequately defined by the GO term collection, the fifth and final step was performed, and gene level annotation was populated from BioMart for each GO term within each biological domain. In this manner we created a fluid system for traversing between high level biological concepts and individual constituent genes. For each of the GO terms enumerated within the biological domains, the genes annotated to that term were retrieved from Ensembl BioMart using the biomaRt R package^109,110^.

#### Target Risk Score (TRS) Development and Process

The goal behind the generation of the TREAT-AD Target Risk Score (TRS) is to develop a scoring infrastructure to objectively, and with minimum bias, assess AD risk association genome wide, leveraging and integrating all available data types. The contributing data types may evolve over time. Here we have initiated the process drawing from genetics, transcriptomics, and proteomics. The composite scoring method delineated below enables us to rank all genes for linkage with AD.

#### Genetic Risk Score Component

The genetic component of AD risk, or genetics score, queries genetic evidence attributable to all loci identified by Ensembl (GRCh38, version 104)(8) as “gene”, “pseudogene” or “ncRNA_gene” resulting in a total 60,664 loci. The score is based on evidence retrieved from both genome wide association studies (GWAS) as well as GWAS by proxy (GWAX) studies^15,16,74,111-123^ and quantitative trait locus (QTL) studies^40,50^ (see Supp Table 3). For consistency across the different GWAS studies used, nominally significant variants (unadjusted p value < 0.05) from anywhere within a 200 kb window surrounding a given target gene’s coordinates are assigned to the gene. For QTL studies, significant variants (FDR < 0.05) must affect the expression of the identified target to be assigned. For each study type (i.e. GWAS or QTL), the score incorporates both the number of studies with significant genetic variants assigned to the target, the minimum significance values of identified variants assigned to the target across studies of that type, and the mean rank of the minimum significance values of identified variants across all studies of that type.

The score also includes further functional characterization of identified variants, both coding and noncoding. The severity of coding and splice-site variants was assessed with ANNOVAR^124^ using the dbnsfp35a^125^ and dbscsnv11^126^ databases. For each variant, the average rank score predictions of deleterious coding and splice site variants were calculated, and the maximum rank score across all variants assigned to a target is reported. In addition, the number and fraction of deleterious coding variants per gene are included, where a variant is called deleterious when at least 3 predictors classified the variant as deleterious. Finally, the coding variant summary score also includes the propensity for each gene to accommodate deleterious coding variants (based on the gnomAD LOEUF score)^127^. For noncoding variants, only those identified in one of the queried QTL studies are considered. The severity of noncoding variants was assessed using both the RegulomeDB (regulomedb.org) ^128^ probability score as well as the DeepSEA (hb.flatironinstitute.org/deepsea) ^129^ mean -log e-value (MLE) variant score.

The score also incorporates phenotypic evidence supporting a given target from both Human and animal model sources. Phenotypes for human genes and orthologs were accessed via the Monarch Initiative API ^41,130^. Human phenotypes for each target were extracted from the Human Phenotype Ontology (hpo.jax.org)^127^ and the number of phenotypes in common with AD (MONDO:0004975) or dementia (MONDO:0001627) are normalized against all phenotypic abnormalities annotated to a gene. Phenotypes of orthologs to human genes extracted from the Unified Phenotype Ontology (uPheno) (https://www.ebi.ac.uk/ols/ontologies/upheno) and the number of phenotypes in common with AD and dementia are normalized against all phenotypic abnormalities annotated to all orthologs of a gene. Finally, the score also includes whether a target has a model in development through the MODEL-AD consortium (model-ad.org/strain-table) as well as the maximum correlation between mouse model gene expression and AMP-AD transcriptional module gene expression^131^.

The Genetic Risk Score for a target (Supp Table 4) is then calculated as the sum of the inverse rank for each of the following evidence categories, scaled to a total of 3 points: number of GWAS studies, minimum GWAS p-value across studies, mean rank of the minimum GWAS p-value across studies, number of QTL studies, minimum QTL FDR across studies, mean rank of the minimum QTL FDR across studies, coding variant summary, noncoding variant summary, human phenotype score, model organism phenotype score, and MODEL-AD strain and correlation.

### Multi-omic Risk Score Component

#### Transcriptomic Weight

A ratio of means meta-analysis with a random effects model^132,133^ was applied to transcriptomics data from RNA-Seq profiling from 8 neocortical tissues to identify differentially expressed features between cases and controls (see Supp Table 6). Feature rank was determined by ordering the absolute fold change between cases and controls and rank was converted to a decimal statistic between zero and one. A logistic regression model was implemented to predict a feature rank from absolute log fold change. This predicted 0 to 1 weight for each feature served as the gene feature’s input transcriptomic weight value to determine a gene feature’s genomic harness weight value and corresponding TREAT-AD genomic score value.

#### Proteomic Weight

A ratio of means meta-analysis with a random effects model^132,133^ was applied to proteomics data from both label-free quantitation (LFQ) and Tandem Mass Tagging (TMT) shot-gun profiling methods generated from 8 neocortical tissues to identify differentially expressed features between cases and controls (see Supp Table 7). Feature rank was determined by ordering the absolute fold change between cases and controls and rank was converted to a decimal statistic between zero and one. A logistic regression model was implemented to predict a feature rank from absolute log fold change. This predicted 0 to 1 weight for each feature served as the gene feature’s input proteomic weight value to determine a gene feature’s genomic harness weight value and corresponding TREAT-AD genomic score value.

#### Multi-omic Harness

To harness both sets of weights into a single value between zero and two to constitute the genomics portion of a gene feature’s contribution to the overall target risk score, a weighted adjustment by weight-modality was applied. Beyond collapsing the weight values into a single statistic, the omics harness is designed to weight proteomics more heavily than transcriptomics to account for the practicality of therapeutic intervention strategies at the protein level rather than the transcript level. Proteomic and Transcriptomic weights were combined by ENSG gene identifiers. In the instance of multiple proteomic identifiers mapping to a single ENSG, the greatest weight value was selected from isoforms which were significant (FDR < 0.05). In the case of multiple isoforms mapping to an ENSG identifier and none were significantly associated with disease, a random isoform’s weight was selected. A scoring harness was applied to combine the Transcriptomic and Proteomic weights of each ENSG gene identifier (Supp Table 8). Genes were ranked by their harness value and ranks were converted to 0-1 decimal. As in the transcriptomic and proteomic weights, a binomial model was fitted to predict 0-1 adjusted rank from the second-degree polynomial of the log harness values. This model was used to compute a predicted genomics weight from zero to one where one corresponds to a greater harness value. For genes with no statistically significant RNA or protein values this omics weight was set to zero. The omics weight was then multiplied by 2 to attain the points value the multi-omic risk score component contributed to a gene’s overall score.

### GSEA Analysis using the Biological Domains

To assess the relative enrichment of different biological domains and their constituent GO terms, gene set enrichment analysis (GSEA) was performed using the gseGO function from the clusterProfiler R package^134^ and the results were then categorized into biological domains based on the GO ID of enriched terms. The input for each enrichment analysis were non-zero target scores, in descending order. We performed this analysis separately for each component score: genetics, multi-omic, and combined target risk. The displayed results include the normalized enrichment score (NES) as well as the Benjamini-Hochberg corrected p-value (p adj) for GO terms annotated to each biological domain. For co-expression module enrichment analyses, the identities of genes and proteins in each module were used for GO term enrichment analysis using the enrichGO function from the clusterProfiler package and the results were then categorized into biological domains based on the GO ID of enriched terms.

### Other data

A number of other data sets are used in this work. The list of AD GWAS hits is derived from integrating genes identified in three sources: the supplementary table that accompanies Neuner *et al* 2020^135^, Supplementary Table 5 from Bellenguez *et al* 2022 that identifies all genome-wide significant loci^15^ and the list of AD loci with genetic evidence compiled by the ADSP Gene Verification Committee (adsp.niagads.org/index.php/gvctop-hits-list/). The Open Targets^43-45,73^ disease association scores for AD (platform.opentargets.org/disease/MONDO_0004975/associations), including data type scores, were accessed using the Open Targets API. The identities of currently nominated targets from the AMP-AD consortium, listed on the Agora site (agora.adknowledgeportal.org/genes/, (https://www.synapse.org/#!Synapse:syn12540368)) were accessed using the synapseR R client^136^.

## Supporting information

Literature Hypothesis Papers

## Data Availability

All data produced in the present work are contained in the manuscript.

## ACKNOWLEDGEMENTS

The authors would like to acknowledge the support of Drs. Lea T. Grinberg, Joshua M. Shulman, David Li-Kroeger, Jessica E. Young, Suman Jayadev, Ranjita Betarbet, and Benoit Lehallier for insightful discussions and suggestions during the development of these resources. The Target Enablement to Accelerate Therapy Development for Alzheimer’s Disease (TREAT-AD) Consortium was established by the National Institute on Aging (NIA). The research reported in this manuscript was led by the Emory-Sage-SGC TREAT center and supported by grant U54AG065187 from the NIA. Certain data used in this study were prepared, archived, and distributed by the National Institute on Aging Alzheimer’s Disease Data Storage Site (NIAGADS) at the University of Pennsylvania (U24AG041689), funded by the NIA; detailed citations and accessions can be found in supplemental table 3. Data used in this study from the Accelerating Medicines Partnership Program for Alzheimer’s Disease (AMP-AD) Consortium members below:

**Mayo RNAseq Study:** Study data were provided by the following sources: The Mayo Clinic Alzheimer’s Disease Genetic Studies, led by Dr. Nilufer Ertekin-Taner and Dr. Steven G. Younkin, Mayo Clinic, Jacksonville, FL using samples from the Mayo Clinic Study of Aging, the Mayo Clinic Alzheimer’s Disease Research Center, and the Mayo Clinic Brain Bank. Data collection was supported through funding by NIA grants P50 AG016574, R01 AG032990, U01 AG046139, R01 AG018023, U01 AG006576, U01 AG006786, R01 AG025711, R01 AG017216, R01 AG003949, NINDS grant R01 NS080820, CurePSP Foundation, and support from Mayo Foundation. Study data includes samples collected through the Sun Health Research Institute Brain and Body Donation Program of Sun City, Arizona. The Brain and Body Donation Program is supported by the National Institute of Neurological Disorders and Stroke (U24 NS072026 National Brain and Tissue Resource for Parkinson’s Disease and Related Disorders), the National Institute on Aging (P30 AG19610 Arizona Alzheimer’s Disease Core Center), the Arizona Department of Health Services (contract 211002, Arizona Alzheimer’s Research Center), the Arizona Biomedical Research Commission (contracts 4001, 0011, 05-901 and 1001 to the Arizona Parkinson’s Disease Consortium) and the Michael J. Fox Foundation for Parkinson’s Research

**ROSMAP:** We are grateful to the participants in the Religious Order Study, the Memory and Aging Project. This work is supported by the US National Institutes of Health [U01 AG046152, R01 AG043617, R01 AG042210, R01 AG036042, R01 AG036836, R01 AG032990, R01 AG18023, RC2 AG036547, P50 AG016574, U01 ES017155, KL2 RR024151, K25 AG041906-01, R01 AG30146, P30 AG10161, R01 AG17917, R01 AG15819, K08 AG034290, P30 AG10161 and R01 AG11101.

**Mount Sinai Brain Bank (MSBB):** This work was supported by the grants R01AG046170, RF1AG054014, RF1AG057440 and R01AG057907 from the NIH/National Institute on Aging (NIA). R01AG046170 is a component of the AMP-AD Target Discovery and Preclinical Validation Project. Brain tissue collection and characterization was supported by NIH HHSN271201300031C.

## Notes

### Competing Interest Statement

F.L. is a founder, has equity interest and serves as a paid consultant for PharmatrophiX, a company executing development of small molecule treatments for neurodegenerative and other disorders. F.L. serves as a paid consultant for Red Tree Venture Capital, a firm investing in biotechnology companies developing treatments for neurological and other disorders. A.L. is a founder, has equity interest and serves as a paid consultant for EmTheraPro, a company developing novel biomarkers and treatments for Alzheimer's disease and related disorders. A.L. serves as a paid consultant for Karuna Pharmaceuticals, Cognito Therapeutics, MEPSGEN, and receives royalties from Linus Health.  

### Author Declarations

Western Institutional Review Board - Copernicus Group (WCG) IRB of Sage Bionetworks gave ethical approval for this work.

