## Supplemental Figures for "Genetic and Multi-omic Risk Assessment of Alzheimer’s Disease Implicates Core Associated Biological Domains"

[a]The Jackson Laboratory, Bar Harbor ME
[b]Sage Bionetworks, Seattle WA
[c]Stanford University, Stanford CA
[d]Emory University, Atlanta GA
[e]adknowledgeportal.org/TREATAD-ESS-Center
[1]These authors participated equally towards the completion of this work

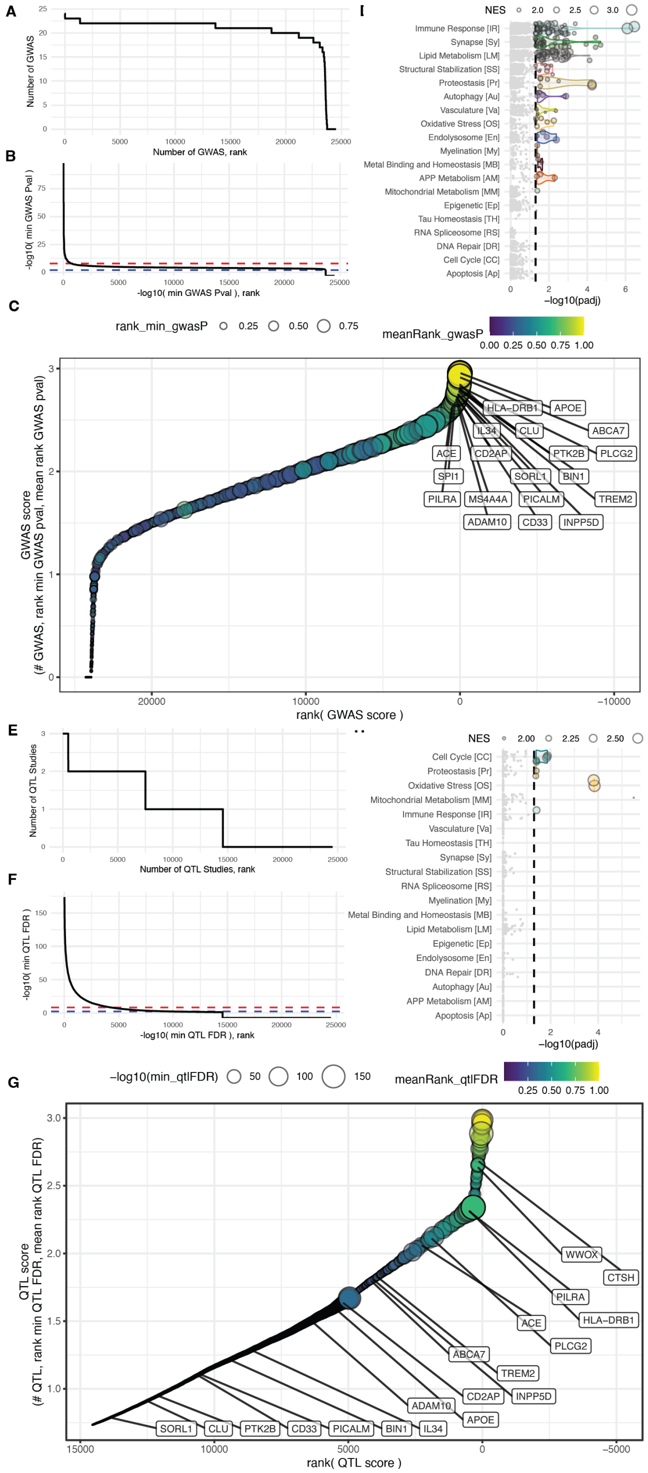


**Fig. 1.** Summary of evidence from genetic association studies. Rank plots of the number of GWAS or GWAX studies (A) as well as eQTL or pQTL (E) where a target is found to be associated with Alzheimer's related traits with at least nominal significance are shown. Rank plots of the minimum p-value of a variant associations with a gene (-log10 transformed) from GWAS/GWAX studies (B) and the minimum significance of a QTL association (F) from any study are shown. The red dashed line shows a typical cutoff for considering an association to be significant genome-wide (i.e. 1x10-8) and the blue dashed line shows a relaxed threshold for inclusion (i.e. 1x10-2). (C) The rank-plot of the combined GWAS score, which includes the number of studies where a gene is significantly associated with AD traits, the rank among all targets of the minimum p-value across all studies, and the average rank of a gene's minimum p-values within each study. The rank minimum GWAS p-value (-log10 transformed) across all studies is displayed as point size whereas the mean rank of p-values within each study is displayed as point color. Several identified GWAS loci are identified and cluster at the top of the distribution. (G) The rank-plot of the combined QTL score, which includes the number of studies where significant variants are found associated with the expression of a gene in AD brain, the rank among all targets of the minimum QTL FDR across all studies, and the average rank of a gene's minimum QTL FDR within each study. The rank minimum QTL FDR across all studies , (-log10 transformed) is displayed as point size whereas the mean rank of FDR within each study is displayed as point color. Several identified GWAS loci are identified, but do not cluster at the top of the distribution nor do we expect they would. Enrichment statistics for all biological domain terms using the combined GWAS score (D) or the combined QTL score (H) to rank targets. Each point is a GO term within the indicated biological domain and the size of the point is scaled by the GSEA normalized enrichment score (NES). The biological domains are ordered on the y-axis by the number of significantly enriched GO terms identified from each domain.

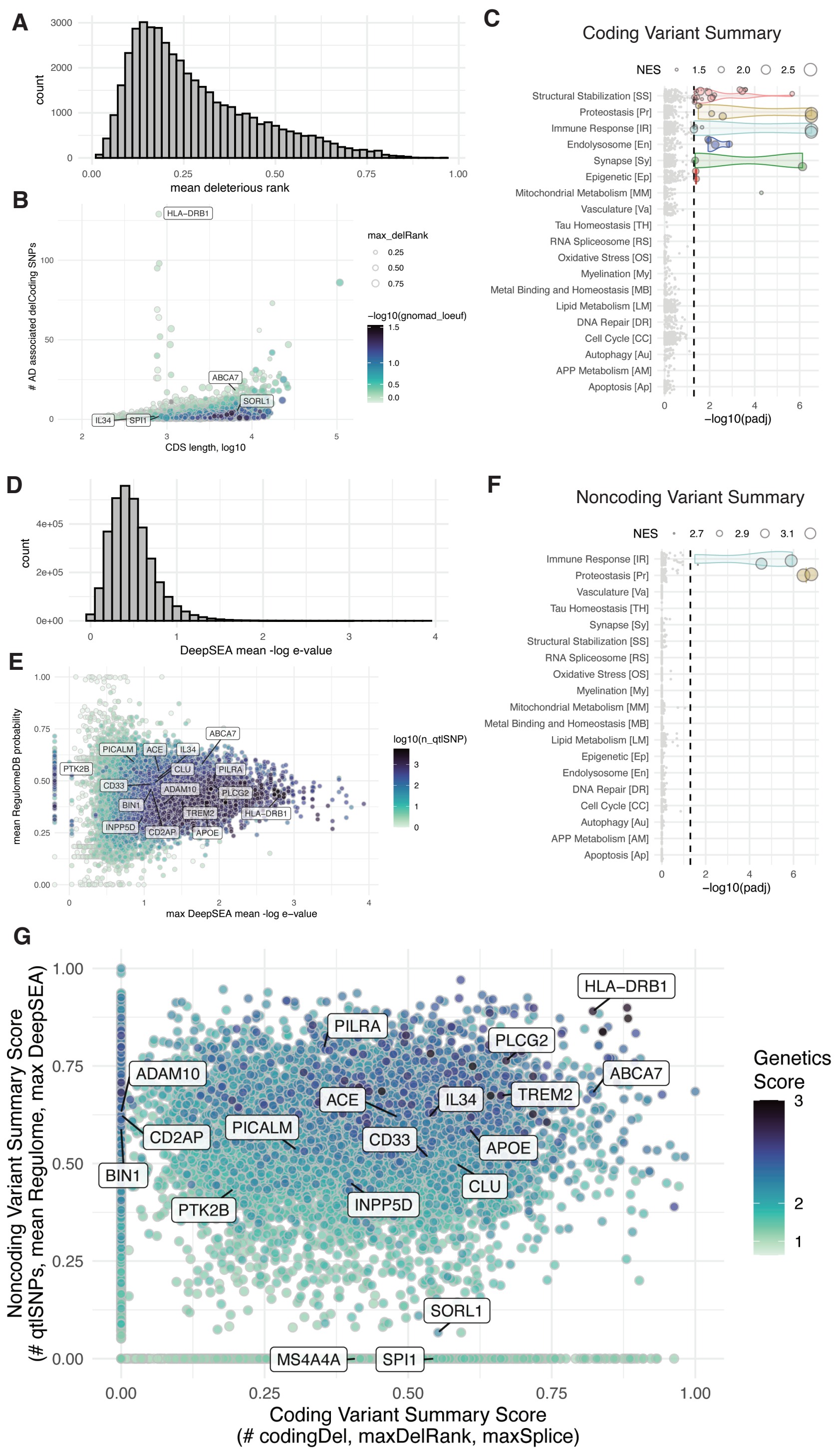


**Fig. 2.** Summary of the assessment of variant severity linked with AD traits. (A) Distribution of the average rank of how deleterious a variant is predicted to be for the 50,833 coding variants identified. (B) The number of AD associated deleterious coding variants associated with a gene is plotted against the length of the coding sequence for each of the 17,892 genes with at least one AD-associated coding variant. The size of each point is the maximum rank of predicted deleterious variants across all associated variants, and the color is the LOEUF score from the gnomAD coalition (-log10 transformed), where low values represent low tolerance for loss-of-function variants for a given gene. Enrichment statistics for all biological domain terms using the coding variant summary score (C) or the noncoding variant summary score (F) to rank targets. Each point is a GO term within the indicated biological domain and the size of the point is scaled by the GSEA normalized enrichment score (NES). The biological domains are ordered on the y-axis by the number of significantly enriched GO terms identified from each domain. (D) Distribution of the average DeepSEA e-value for the 1,365,759 noncoding variants associated with altered expression of a target gene in AD brains. (E) Comparison of the maximum DeepSEA e-value for variants associated with a target versus the average RegulomeDB probability score for associated variants. The color represents the number of distinct variants associated with altered expression of the given target gene, log10 transformed. (G) Comparison of the coding variant summary score - comprised of the number of deleterious coding variants identified, the maximum rank of how deleterious a variant is predicted to be, as well as the maximum score for variants predicted to affect splicing - to the noncoding variant summary score - comprised of the number of SNPs associated with altered expression of the gene, the mean Regulome DB probability score for variants associated with a gene, and the maximum DeepSEA -log10(e-value) for variants associated with a gene. The points are colored by the value of the final Genetics Score. For all scatterplots, the location of genes from several loci implicated by GWAS are indicated.

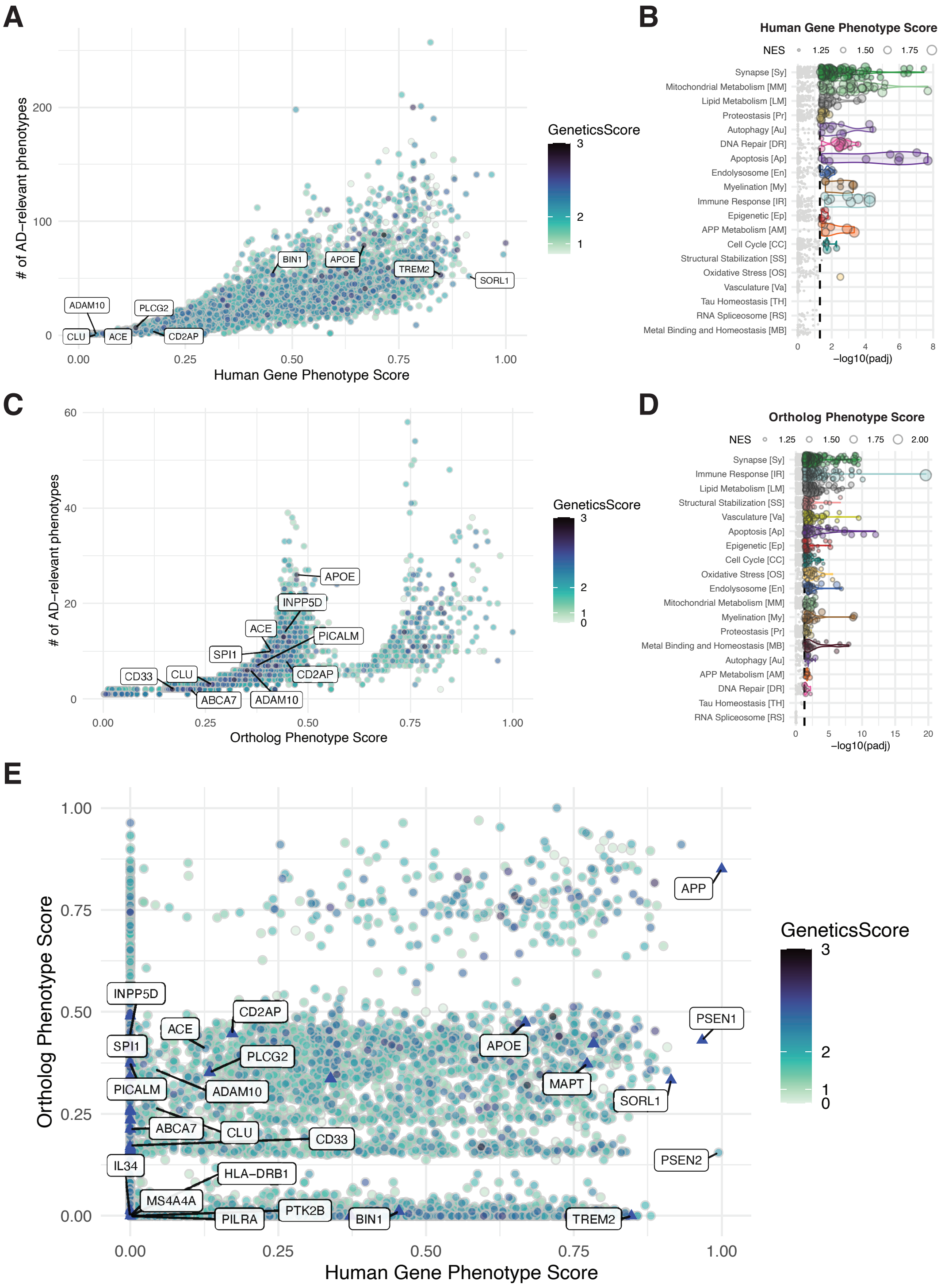


**Fig. 3.** Summary of the assessment of related phenotypes. (A) The combined human phenotype score for each gene is plotted against the total number of AD-relevant phenotypes attributable to that gene. The point color represents the final Genetics Score for each gene. Enrichment statistics for all biological domain terms using the human phenotype summary score (B) or the ortholog phenotype summary score (D) to rank targets. Each point is a GO term within the indicated biological domain and the size of the point is scaled by the GSEA normalized enrichment score (NES). The biological domains are ordered on the y-axis by the number of significantly enriched GO terms identified from each domain. (C) The combined phenotype score for model organism phenotypes is plotted against the number of AD-relevant phenotypes attributable to that gene. The point color represents the final Genetics Score for each gene. The bi-modality of the ortholog phenotype score generally reflects genes that have orthologs associated with multiple AD-relevant phenotypes (0.5-1) versus those associated with only dementia-relevant phenotypes (0-0.5). (E) Comparison of the human phenotype score and the ortholog phenotype score. The point color represents the final Genetics Score for each gene. Genes for which there is a relevant model generated as part of MODEL-AD efforts are shown as a triangle. For all scatterplots, the location of genes from several loci implicated by GWAS are indicated.

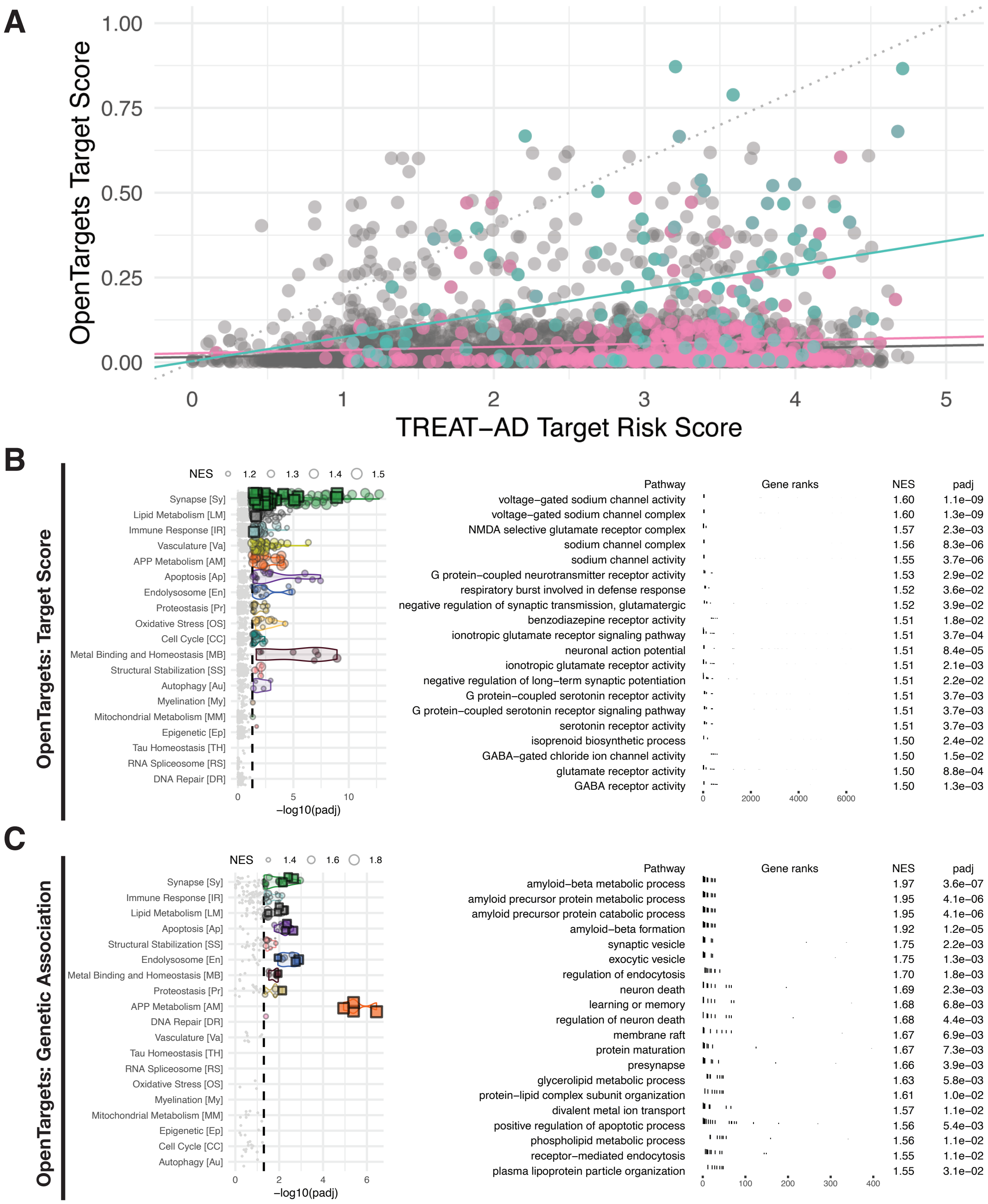


**Fig. 4.** Comparison of computed AD risk scores with the OpenTargets risk scores. (A) Comparison of the TREAT-AD target risk score (TRS) with the OpenTargets target score for all genes. Genes nominated by members of the AMP-AD consortium are shaded in pink, whereas genes implicated within known GWAS loci are shaded in green. Biological domain term enrichments for the OpenTargets target score (B) and the OpenTargets genetics score (C). In each panel, the plot on the left shows the enrichment statistics for all biological domain terms. Each point is a GO term within the indicated biological domain and the size of the point is scaled by the GSEA normalized enrichment score (NES). The biological domains are ordered on the y-axis by the number of significantly enriched GO terms identified from each domain. The plot on the right shows the top GO terms significantly enriched using the TREAT-AD target genetics score, arranged by normalized enrichment score (NES).

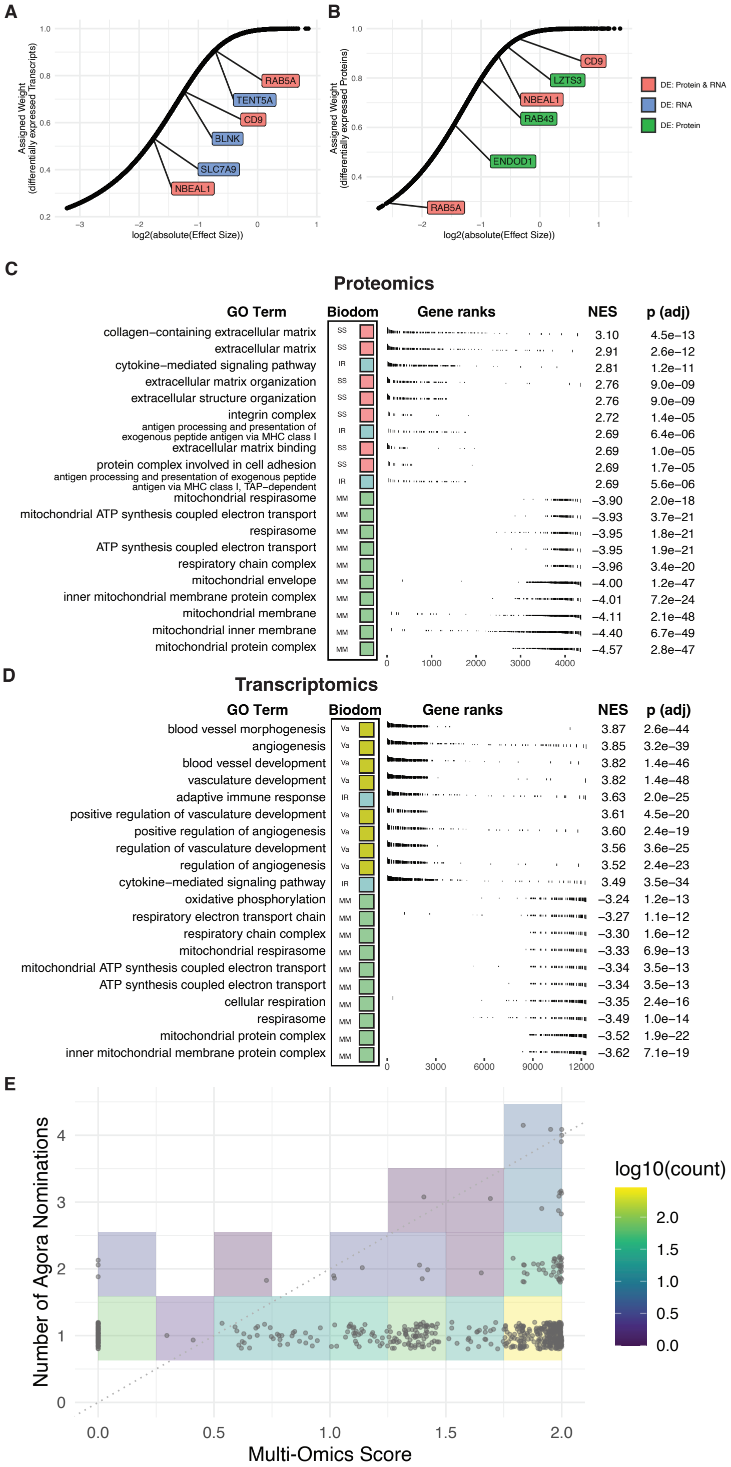


**Fig. 5.** Features of the Multi-Omics score. Assigned weights as a function of log( effect size ) of significantly differentially expressed transcripts (A) and proteins (B). Identified genes are those significantly differentially expressed in only RNA-seq (blue), only proteomics (green), and both RNA-Seq and proteomics (red). Representative genes were chosen at the first quartile, median, and third quartile range of effect size from genes with only RNA differential expression (blue), proteins (green), and combined weight value distribution of genes with both differentially expressed RNA and proteomics signal (red). Top GO terms significantly enriched using the proteomics meta-analysis treatment effect (C) and RNA-seq meta-analysis treatment effect (D), arranged by normalized enrichment score (NES). For each GO term, the associated biological domain(s) are indicated by the filled square and abbreviations. (E) Multi-Omics score for each target plotted against the number of nominations targets have received from AMP-AD investigators as reported on Agora (agora.adknowledgeportal.org/genes/).

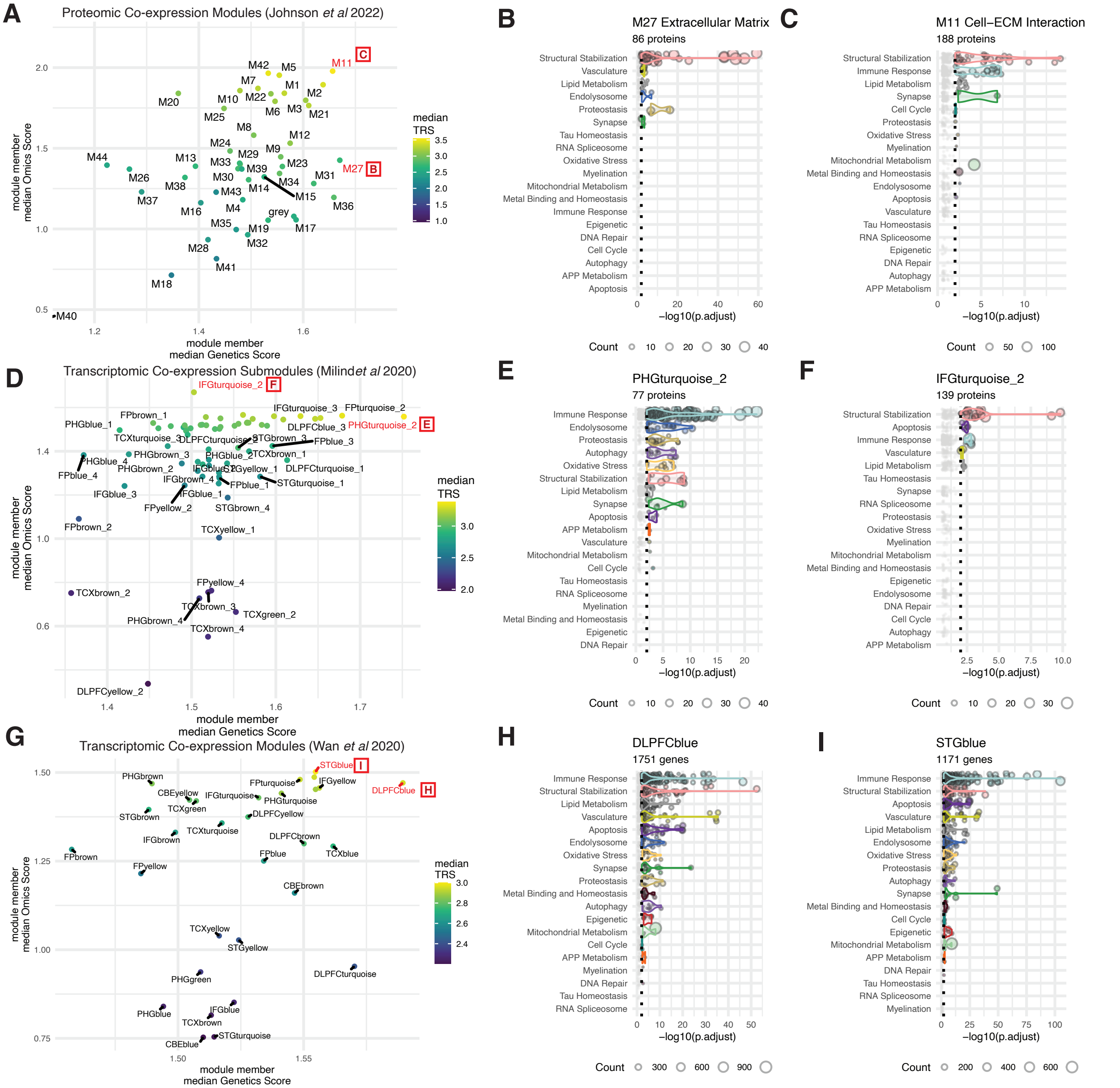


**Fig. 6.** Transcriptomic and proteomic module scores and biological domains. For each proteomics co-expression module from Johnson et al (A), transcriptomic co-expression sub-module from Milind et al (D), and transcriptomic co-expression module from Wan et al (G) the median genetics score is plotted against the median omics score for all genes or proteins within the module. Each point is colored by the median Target Risk Score (TRS) for all genes or proteins within the module. For the indicated modules (in red) from the overall sets in A, D, and G, the biological domain term enrichments using a hypergeometric overrepresentation test are plotted (B-C, E-F, H-I). Each point is a GO term within the indicated biological domain and the size of the point is scaled by the number of proteins or genes from the module that are annotated to that term. The biological domains are ordered on the y-axis by the number of significantly enriched GO terms identified from each domain.

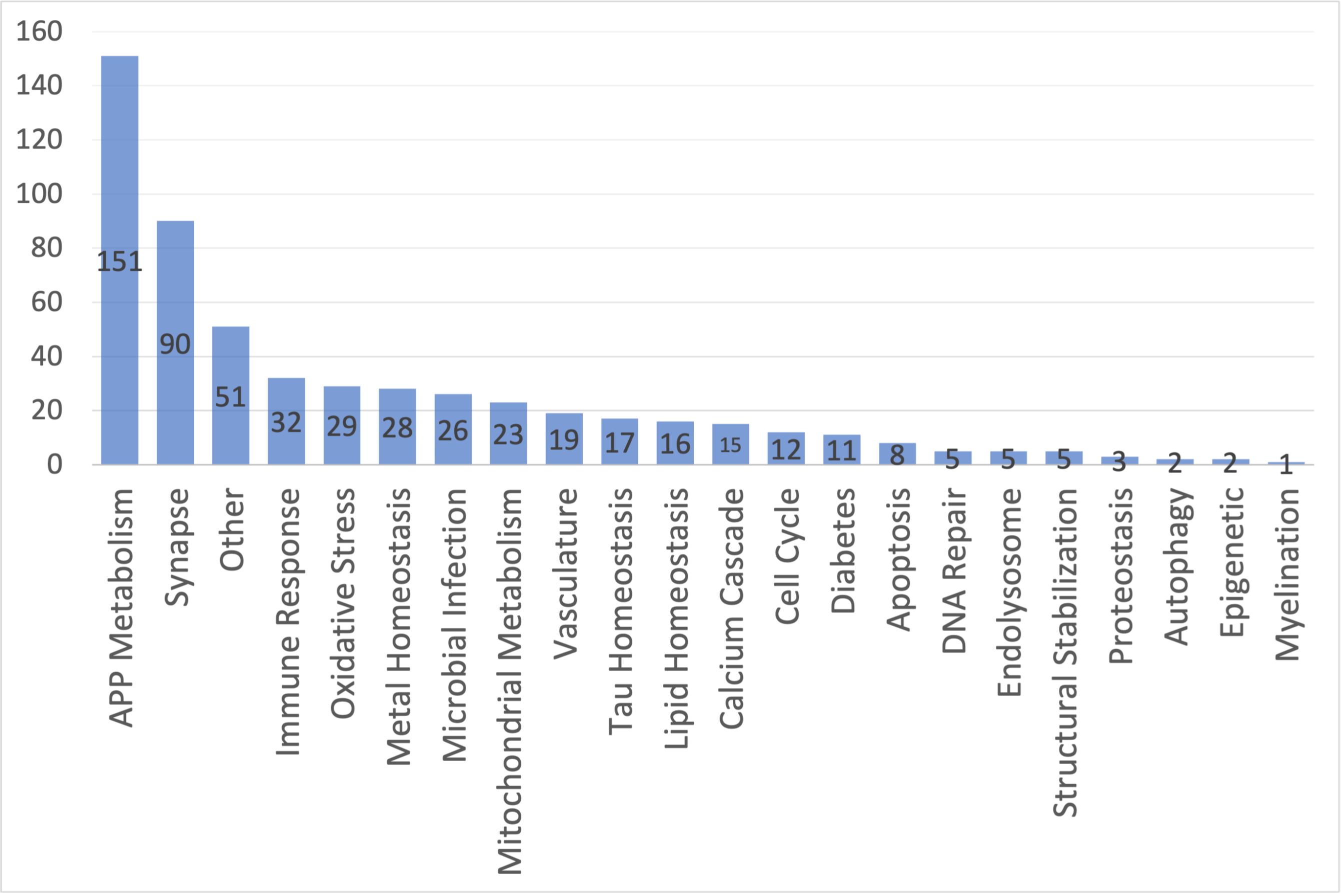


**Fig. 7.** Quantitative assessment of published AD hypotheses. The publically available literature was queried using a PubMed search for “Alzheimer” AND “hypothesis” in the article title (last updated: Oct 19, 2022), which resulted in 463 identified papers. The published hypotheses were organized by the primary subject matter posited by the article, and aligned with the existing biological domains, when appropriate. The graph above shows the number of articles across 22 different hypothesis topics. The largest group of papers fell within the amyloid hypothesis, with 151 published papers, followed by Synapse (90 papers) and Immune response (32 papers). There were 52 papers that discussed hypotheses that did not bin into molecular etiological descriptions, focusing on life-style or device mediated approaches to treating or monitoring AD. Twenty-two categories were identified in total, and 19 are instantiated as biological domains.

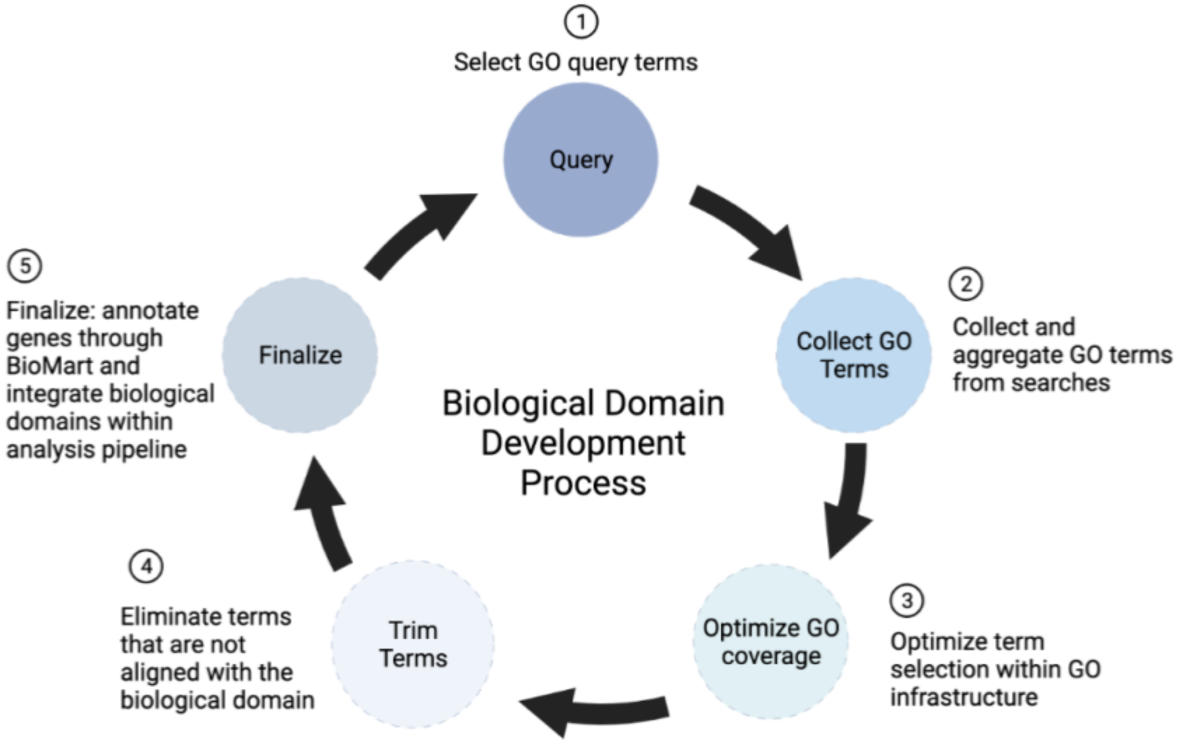


**Fig. 8.** Demonstration of the developmental process for each biological domain. Step 1 Launch exhaustive GO searches using terms definitional to that biological domain. Step 2 Collect an unabridged set of GO terms returned from the searches. Step 3 Select the GO terms with annotated gene sets and significant centrality with the GO. Expand to related terms not previously identified by direct examination of the ontology tree and neighbor terms. Step 4 prune out the GO terms that are not unique and consistent with the specific meaning of that biological domain. Step 5 finalize the biological domain by populating it at the gene level using BioMart to complete the construction of the biological domain at the (a) conceptual (b) GO term and (c) gene specific levels.
