## Supplementary material for "Genetic and Multi-omic Risk Assessment of Alzheimer’s Disease Implicates Core Associated Biological Domains": Literature Hypothesis Papers

Literature Hypotheses (ranked by prevalence):

1. Amyloid hypothesis [1-151]
2. Synapse Hypothesis [21,35,60,83,97,110,134,142-144,147,152-230]
3. Other [34,100,140,141,223,231-276]
4. Immune Response Hypothesis [3,15,25,38,68,70,125,229,277-300]
5. Oxidative Stress Hypothesis [2,11,86,131,134,278,289,301-322]
6. Metal Binding and Homeostasis Hypothesis [10,69,86,114,125,150,155,282,298,303,304,323-339]
7. Microbial Infection Hypothesis [3,38,155,238,281,283,285,287,288,293,300,340-353]
8. Mitochondrial Metabolism Hypothesis [62,256,289,312,318,354-371]
9. Vasculature Hypothesis [20,69,95,163,185,248,249,290,372-382]
10. Tau Homeostasis Hypothesis [34,37,39,56,71,98,148,156,184,284,336,383-388]
11. Lipid Homeostasis Hypothesis [229,237,255,278,360,387,389-398]
12. Calcium Cascade Hypothesis [146,168,388,399-410]
13. Cell Cycle Hypothesis [256,309,313,316,407,411-417]
14. Diabetes Hypothesis [101,241,291,311,418-424]
15. Apoptosis Hypothesis [16,52,136,146,147,188,402,425]
16. DNA Repair Hypothesis [2,11,290,304,426,427]
17. Endolysosome Hypothesis [237,402,404,428,429]
18. Structural Stabilization Hypothesis [277,430-433]
19. Proteostasis Hypothesis [434-436]
20. Autophagy Hypothesis [402,404]
21. Epigenetic [78,437]
22. Myelination [163]


1 Volloch, V. & Rits-Volloch, S. The Amyloid Cascade Hypothesis 2.0: On the Possibility of Once-in-a-Lifetime-Only Treatment for Prevention of Alzheimer's Disease and for Its Potential Cure at Symptomatic Stages. *J Alzheimers Dis Rep* **6**, 369-399, doi:10.3233/adr-220031 (2022).

2 Sanders, O. D., Rajagopal, L. & Rajagopal, J. A. The oxidatively damaged DNA and amyloid-β oligomer hypothesis of Alzheimer's disease. *Free Radic Biol Med* **179**, 403-412, doi:10.1016/j.freeradbiomed.2021.08.019 (2022).

3 Phuna, Z. X. & Madhavan, P. A reappraisal on amyloid cascade hypothesis: the role of chronic infection in Alzheimer's disease. *Int J Neurosci*, 1-19, doi:10.1080/00207454.2022.2045290 (2022).

4 Levin, J. *et al.* Testing the amyloid cascade hypothesis: Prevention trials in autosomal dominant Alzheimer disease. *Alzheimers Dement*, doi:10.1002/alz.12624 (2022).

5 Krafft, G. A., Jerecic, J., Siemers, E. & Cline, E. N. ACU193: An Immunotherapeutic Poised to Test the Amyloid β Oligomer Hypothesis of Alzheimer's Disease. *Front Neurosci* **16**, 848215, doi:10.3389/fnins.2022.848215 (2022).

6 Karran, E. & De Strooper, B. The amyloid hypothesis in Alzheimer disease: new insights from new therapeutics. *Nat Rev Drug Discov* **21**, 306-318, doi:10.1038/s41573-022-00391-w (2022).

7 Frisoni, G. B. *et al.* The probabilistic model of Alzheimer disease: the amyloid hypothesis revised. *Nat Rev Neurosci* **23**, 53-66, doi:10.1038/s41583-021-00533-w (2022).
8 Alawode, D. O. T., Fox, N. C., Zetterberg, H. & Heslegrave, A. J. Alzheimer's Disease Biomarkers Revisited From the Amyloid Cascade Hypothesis Standpoint. *Front Neurosci* **16**, 837390, doi:10.3389/fnins.2022.837390 (2022).
9 Zaretsky, D. V. & Zaretskaia, M. V. Mini-review: Amyloid degradation toxicity hypothesis of Alzheimer's disease. *Neurosci Lett* **756**, 135959, doi:10.1016/j.neulet.2021.135959 (2021).
10 Squitti, R. *et al.* Copper Imbalance in Alzheimer's Disease and Its Link with the Amyloid Hypothesis: Towards a Combined Clinical, Chemical, and Genetic Etiology. *J Alzheimers Dis* **83**, 23-41, doi:10.3233/jad-201556 (2021).
11 Sanders, O. D., Rajagopal, L. & Rajagopal, J. A. Does oxidatively damaged DNA drive amyloid-β generation in Alzheimer's disease? A hypothesis. *J Neurogenet* **35**, 351-357, doi:10.1080/01677063.2021.1954641 (2021).
12 Richard, E., den Brok, M. & van Gool, W. A. Bayes analysis supports null hypothesis of anti-amyloid beta therapy in Alzheimer's disease. *Alzheimers Dement* **17**, 1051-1055, doi:10.1002/alz.12379 (2021).
13 Okazawa, H. Intracellular amyloid hypothesis for ultra-early phase pathology of Alzheimer's disease. *Neuropathology* **41**, 93-98, doi:10.1111/neup.12738 (2021).
14 Musiek, E. S., Gomez-Isla, T. & Holtzman, D. M. Aducanumab for Alzheimer disease: the amyloid hypothesis moves from bench to bedside. *J Clin Invest* **131**, doi:10.1172/jci154889 (2021).
15 Lim, B., Prassas, I. & Diamandis, E. P. From the amyloid hypothesis to the autoimmune hypothesis of Alzheimer's disease. *Diagnosis (Berl)* **9**, 280-281, doi:10.1515/dx-2021-0108 (2021).
16 Inagaki, C. [Amyloid β hypothesis in Alzheimer's disease and Cl(-)-ATPase-Neuronal cell death via PI4KIIα inhibition and recovery agents]. *Nihon Yakurigaku Zasshi* **156**, 166-170, doi:10.1254/fpj.20095 (2021).
17 Vijayan, D. & Chandra, R. Amyloid Beta Hypothesis in Alzheimer's Disease: Major Culprits and Recent Therapeutic Strategies. *Curr Drug Targets* **21**, 148-166, doi:10.2174/1389450120666190806153206 (2020).
18 Uddin, M. S. *et al.* Revisiting the Amyloid Cascade Hypothesis: From Anti-Aβ Therapeutics to Auspicious New Ways for Alzheimer's Disease. *Int J Mol Sci* **21**, doi:10.3390/ijms21165858 (2020).
19 Tolar, M., Abushakra, S. & Sabbagh, M. The path forward in Alzheimer's disease therapeutics: Reevaluating the amyloid cascade hypothesis. *Alzheimers Dement* **16**, 1553-1560, doi:10.1016/j.jalz.2019.09.075 (2020).
20 Stakos, D. A. *et al.* The Alzheimer's Disease Amyloid-Beta Hypothesis in Cardiovascular Aging and Disease: JACC Focus Seminar. *J Am Coll Cardiol* **75**, 952-967, doi:10.1016/j.jacc.2019.12.033 (2020).
21 Li, S. & Selkoe, D. J. A mechanistic hypothesis for the impairment of synaptic plasticity by soluble Aβ oligomers from Alzheimer's brain. *J Neurochem* **154**, 583-597, doi:10.1111/jnc.15007 (2020).

22 Imbimbo, B. P., Ippati, S. & Watling, M. Should drug discovery scientists still embrace the amyloid hypothesis for Alzheimer's disease or should they be looking elsewhere? *Expert Opin Drug Discov* **15**, 1241-1251, doi:10.1080/17460441.2020.1793755 (2020).
23 Daly, T., Houot, M., Barberousse, A., Agid, Y. & Epelbaum, S. Amyloid-β in Alzheimer's Disease: A Study of Citation Practices of the Amyloid Cascade Hypothesis Between 1992 and 2019. *J Alzheimers Dis* **74**, 1309-1317, doi:10.3233/jad-191321 (2020).
24 Caselli, R. J., Knopman, D. S. & Bu, G. An agnostic reevaluation of the amyloid cascade hypothesis of Alzheimer's disease pathogenesis: The role of APP homeostasis. *Alzheimers Dement* **16**, 1582-1590, doi:10.1002/alz.12124 (2020).
25 Arshavsky, Y. I. Alzheimer's Disease: From Amyloid to Autoimmune Hypothesis. *Neuroscientist* **26**, 455-470, doi:10.1177/1073858420908189 (2020).
26 Zhang, H. & Zheng, Y. [β Amyloid Hypothesis in Alzheimer's Disease:Pathogenesis,Prevention,and Management]. *Zhongguo Yi Xue Ke Xue Yuan Xue Bao* **41**, 702-708, doi:10.3881/j.issn.1000-503X.10875 (2019).
27 Paroni, G., Bisceglia, P. & Seripa, D. Understanding the Amyloid Hypothesis in Alzheimer's Disease. *J Alzheimers Dis* **68**, 493-510, doi:10.3233/jad-180802 (2019).
28 Panza, F., Lozupone, M., Watling, M. & Imbimbo, B. P. Do BACE inhibitor failures in Alzheimer patients challenge the amyloid hypothesis of the disease? *Expert Rev Neurother* **19**, 599-602, doi:10.1080/14737175.2019.1621751 (2019).
29 Panza, F. *et al.* Are antibodies directed against amyloid-β (Aβ) oligomers the last call for the Aβ hypothesis of Alzheimer's disease? *Immunotherapy* **11**, 3-6, doi:10.2217/imt-2018-0119 (2019).
30 Modrego, P. & Lobo, A. A good marker does not mean a good target for clinical trials in Alzheimer's disease: the amyloid hypothesis questioned. *Neurodegener Dis Manag* **9**, 119-121, doi:10.2217/nmt-2019-0006 (2019).
31 McDade, E. Reply to: Major Clinical Trials Failed the Amyloid Hypothesis of Alzheimer's Disease. *J Am Geriatr Soc* **67**, 848-849, doi:10.1111/jgs.15826 (2019).
32 Huang, Y. M., Shen, J. & Zhao, H. L. Major Clinical Trials Failed the Amyloid Hypothesis of Alzheimer's Disease. *J Am Geriatr Soc* **67**, 841-844, doi:10.1111/jgs.15830 (2019).
33 Hillen, H. The Beta Amyloid Dysfunction (BAD) Hypothesis for Alzheimer's Disease. *Front Neurosci* **13**, 1154, doi:10.3389/fnins.2019.01154 (2019).
34 Duyckaerts, C., Clavaguera, F. & Potier, M. C. The prion-like propagation hypothesis in Alzheimer's and Parkinson's disease. *Curr Opin Neurol* **32**, 266-271, doi:10.1097/wco.0000000000000672 (2019).
35 Dourlen, P., Kilinc, D., Malmanche, N., Chapuis, J. & Lambert, J. C. The new genetic landscape of Alzheimer's disease: from amyloid cascade to genetically driven synaptic failure hypothesis? *Acta Neuropathol* **138**, 221-236, doi:10.1007/s00401-019-02004-0 (2019).
36 Mullane, K. & Williams, M. Alzheimer's disease (AD) therapeutics - 1: Repeated clinical failures continue to question the amyloid hypothesis of AD and the current understanding of AD causality. *Biochem Pharmacol* **158**, 359-375, doi:10.1016/j.bcp.2018.09.026 (2018).

37 Kametani, F. & Hasegawa, M. Reconsideration of Amyloid Hypothesis and Tau Hypothesis in Alzheimer's Disease. *Front Neurosci* **12**, 25, doi:10.3389/fnins.2018.00025 (2018).
38 Fulop, T. *et al.* Can an Infection Hypothesis Explain the Beta Amyloid Hypothesis of Alzheimer's Disease? *Front Aging Neurosci* **10**, 224, doi:10.3389/fnagi.2018.00224 (2018).
39 Beyond the Amyloid Hypothesis of Alzheimer's Disease: Tau Pathology Takes Center Stage. *ACS Chem Neurosci* **9**, 2519, doi:10.1021/acschemneuro.8b00610 (2018).
40 Ricciarelli, R. & Fedele, E. The Amyloid Cascade Hypothesis in Alzheimer's Disease: It's Time to Change Our Mind. *Curr Neuropharmacol* **15**, 926-935, doi:10.2174/1570159x15666170116143743 (2017).
41 Murray, B., Sharma, B. & Belfort, G. N-Terminal Hypothesis for Alzheimer's Disease. *ACS Chem Neurosci* **8**, 432-434, doi:10.1021/acschemneuro.7b00037 (2017).
42 Luo, J., Wärmländer, S., Gräslund, A. & Abrahams, J. P. Cross-interactions between the Alzheimer disease amyloid-β peptide and other amyloid proteins. A FURTHER ASPECT OF THE AMYLOID CASCADE HYPOTHESIS. *J Biol Chem* **292**, 2046, doi:10.1074/jbc.A116.714576 (2017).
43 Kepp, K. P. Ten Challenges of the Amyloid Hypothesis of Alzheimer's Disease. *J Alzheimers Dis* **55**, 447-457, doi:10.3233/jad-160550 (2017).
44 Hardy, J. The discovery of Alzheimer-causing mutations in the APP gene and the formulation of the "amyloid cascade hypothesis". *Febs j* **284**, 1040-1044, doi:10.1111/febs.14004 (2017).
45 Selkoe, D. J. & Hardy, J. The amyloid hypothesis of Alzheimer's disease at 25 years. *EMBO Mol Med* **8**, 595-608, doi:10.15252/emmm.201606210 (2016).
46 Mullard, A. Alzheimer amyloid hypothesis lives on. *Nat Rev Drug Discov* **16**, 3-5, doi:10.1038/nrd.2016.281 (2016).
47 Luo, J., Wärmländer, S. K., Gräslund, A. & Abrahams, J. P. Cross-interactions between the Alzheimer Disease Amyloid-β Peptide and Other Amyloid Proteins: A Further Aspect of the Amyloid Cascade Hypothesis. *J Biol Chem* **291**, 16485-16493, doi:10.1074/jbc.R116.714576 (2016).
48 Levin, O. S. & Vasenina, E. E. [Twenty-five years of the amyloid hypothesis of alzheimer disease: advances, failures and new perspectives]. *Zh Nevrol Psikhiatr Im S S Korsakova* **116**, 3-9, doi:10.17116/jnevro2016116623-9 (2016).
49 Le Couteur, D. G., Hunter, S. & Brayne, C. Solanezumab and the amyloid hypothesis for Alzheimer's disease. *Bmj* **355**, i6771, doi:10.1136/bmj.i6771 (2016).
50 Kretner, B. *et al.* Generation and deposition of Aβ43 by the virtually inactive presenilin-1 L435F mutant contradicts the presenilin loss-of-function hypothesis of Alzheimer's disease. *EMBO Mol Med* **8**, 458-465, doi:10.15252/emmm.201505952 (2016).
51 Harrison, J. R. & Owen, M. J. Alzheimer's disease: the amyloid hypothesis on trial. *Br J Psychiatry* **208**, 1-3, doi:10.1192/bjp.bp.115.167569 (2016).
52 Tanila, H. [Curative treatment of Alzheimer's disease is still missing--is amyloid plaque hypothesis the wrong starting point?]. *Duodecim* **131**, 243-248 (2015).

53 Suzuki, Y. *et al.* Reduced CSF Water Influx in Alzheimer's Disease Supporting the β-Amyloid Clearance Hypothesis. *PLoS One* **10**, e0123708, doi:10.1371/journal.pone.0123708 (2015).
54 Parodi, J., Ormeño, D. & Ochoa-de la Paz, L. D. Amyloid pore-channel hypothesis: effect of ethanol on aggregation state using frog oocytes for an Alzheimer's disease study. *BMB Rep* **48**, 13-18, doi:10.5483/bmbrep.2015.48.1.125 (2015).
55 Barage, S. H. & Sonawane, K. D. Amyloid cascade hypothesis: Pathogenesis and therapeutic strategies in Alzheimer's disease. *Neuropeptides* **52**, 1-18, doi:10.1016/j.npep.2015.06.008 (2015).
56 Morris, G. P., Clark, I. A. & Vissel, B. Inconsistencies and controversies surrounding the amyloid hypothesis of Alzheimer's disease. *Acta Neuropathol Commun* **2**, 135, doi:10.1186/s40478-014-0135-5 (2014).
57 Lin, L. X. *et al.* Feasibility of β-sheet breaker peptide-H102 treatment for Alzheimer's disease based on β-amyloid hypothesis. *PLoS One* **9**, e112052, doi:10.1371/journal.pone.0112052 (2014).
58 Karran, E. & Hardy, J. A critique of the drug discovery and phase 3 clinical programs targeting the amyloid hypothesis for Alzheimer disease. *Ann Neurol* **76**, 185-205, doi:10.1002/ana.24188 (2014).
59 Igarashi, H., Suzuki, Y., Kwee, I. L. & Nakada, T. Water influx into cerebrospinal fluid is significantly reduced in senile plaque bearing transgenic mice, supporting beta-amyloid clearance hypothesis of Alzheimer's disease. *Neurol Res* **36**, 1094-1098, doi:10.1179/1743132814y.0000000434 (2014).
60 Gouras, G. K., Willén, K. & Faideau, M. The inside-out amyloid hypothesis and synapse pathology in Alzheimer's disease. *Neurodegener Dis* **13**, 142-146, doi:10.1159/000354776 (2014).
61 Drachman, D. A. The amyloid hypothesis, time to move on: Amyloid is the downstream result, not cause, of Alzheimer's disease. *Alzheimers Dement* **10**, 372-380, doi:10.1016/j.jalz.2013.11.003 (2014).
62 Demetrius, L. A., Magistretti, P. J. & Pellerin, L. Alzheimer's disease: the amyloid hypothesis and the Inverse Warburg effect. *Front Physiol* **5**, 522, doi:10.3389/fphys.2014.00522 (2014).
63 Castello, M. A. & Soriano, S. On the origin of Alzheimer's disease. Trials and tribulations of the amyloid hypothesis. *Ageing Res Rev* **13**, 10-12, doi:10.1016/j.arr.2013.10.001 (2014).
64 Bayer, T. A. & Wirths, O. Focusing the amyloid cascade hypothesis on N-truncated Abeta peptides as drug targets against Alzheimer's disease. *Acta Neuropathol* **127**, 787-801, doi:10.1007/s00401-014-1287-x (2014).
65 Tayeb, H. O., Murray, E. D., Price, B. H. & Tarazi, F. I. Bapineuzumab and solanezumab for Alzheimer's disease: is the 'amyloid cascade hypothesis' still alive? *Expert Opin Biol Ther* **13**, 1075-1084, doi:10.1517/14712598.2013.789856 (2013).
66 Tamaoka, A. [The pathophysiology of Alzheimer's disease with special reference to "amyloid cascade hypothesis"]. *Rinsho Byori* **61**, 1060-1069 (2013).

67 Mullane, K. & Williams, M. Alzheimer's therapeutics: continued clinical failures question the validity of the amyloid hypothesis-but what lies beyond? *Biochem Pharmacol* **85**, 289-305, doi:10.1016/j.bcp.2012.11.014 (2013).
68 McGeer, P. L. & McGeer, E. G. The amyloid cascade-inflammatory hypothesis of Alzheimer disease: implications for therapy. *Acta Neuropathol* **126**, 479-497, doi:10.1007/s00401-013-1177-7 (2013).
69 Lucas, H. R. & Rifkind, J. M. Considering the vascular hypothesis of Alzheimer's disease: effect of copper associated amyloid on red blood cells. *Adv Exp Med Biol* **765**, 131-138, doi:10.1007/978-1-4614-4989-8_19 (2013).
70 Gandy, S., Haroutunian, V., DeKosky, S. T., Sano, M. & Schadt, E. E. CR1 and the "vanishing amyloid" hypothesis of Alzheimer's disease. *Biol Psychiatry* **73**, 393-395, doi:10.1016/j.biopsych.2013.01.013 (2013).
71 Braak, H. & Del Tredici, K. Amyloid-β may be released from non-junctional varicosities of axons generated from abnormal tau-containing brainstem nuclei in sporadic Alzheimer's disease: a hypothesis. *Acta Neuropathol* **126**, 303-306, doi:10.1007/s00401-013-1153-2 (2013).
72 Teich, A. F. & Arancio, O. Is the amyloid hypothesis of Alzheimer's disease therapeutically relevant? *Biochem J* **446**, 165-177, doi:10.1042/bj20120653 (2012).
73 Reitz, C. Alzheimer's disease and the amyloid cascade hypothesis: a critical review. *Int J Alzheimers Dis* **2012**, 369808, doi:10.1155/2012/369808 (2012).
74 Morelli, L., Perry, G. & Tagliavini, F. The contribution of the amyloid hypothesis to the understanding of Alzheimer's disease: a critical overview. *Int J Alzheimers Dis* **2012**, 709613, doi:10.1155/2012/709613 (2012).
75 Kung, H. F. The β-Amyloid Hypothesis in Alzheimer's Disease: Seeing Is Believing. *ACS Med Chem Lett* **3**, 265-267, doi:10.1021/ml300058m (2012).
76 Dong, S., Duan, Y., Hu, Y. & Zhao, Z. Advances in the pathogenesis of Alzheimer's disease: a re-evaluation of amyloid cascade hypothesis. *Transl Neurodegener* **1**, 18, doi:10.1186/2047-9158-1-18 (2012).
77 Checler, F. & Turner, A. J. Journal of Neurochemistry special issue on Alzheimer's disease: 'amyloid cascade hypothesis--20 years on'. *J Neurochem* **120 Suppl 1**, iii-iv, doi:10.1111/j.1471-4159.2011.07603.x (2012).
78 Bórquez, D. A. & González-Billault, C. The amyloid precursor protein intracellular domain-fe65 multiprotein complexes: a challenge to the amyloid hypothesis for Alzheimer's disease? *Int J Alzheimers Dis* **2012**, 353145, doi:10.1155/2012/353145 (2012).
79 Wostyn, P., van Dam, D., Audenaert, K. & de Deyn, P. P. Genes involved in cerebrospinal fluid production as candidate genes for late-onset Alzheimer's disease: a hypothesis. *J Neurogenet* **25**, 195-200, doi:10.3109/01677063.2011.620191 (2011).
80 Li, N. [Progress of treating Alzheimer's diseases by "A beta cascade hypothesis" based Chinese materia medica]. *Zhongguo Zhong Xi Yi Jie He Za Zhi* **31**, 1714-1720 (2011).
81 Lambert, J. C. & Amouyel, P. Genetics of Alzheimer's disease: new evidences for an old hypothesis? *Curr Opin Genet Dev* **21**, 295-301, doi:10.1016/j.gde.2011.02.002 (2011).

82 Karran, E., Mercken, M. & De Strooper, B. The amyloid cascade hypothesis for Alzheimer's disease: an appraisal for the development of therapeutics. *Nat Rev Drug Discov* **10**, 698-712, doi:10.1038/nrd3505 (2011).

83 Ferreira, S. T. & Klein, W. L. The Aβ oligomer hypothesis for synapse failure and memory loss in Alzheimer's disease. *Neurobiol Learn Mem* **96**, 529-543, doi:10.1016/j.nlm.2011.08.003 (2011).

84 Cummings, J. Alzheimer's disease: clinical trials and the amyloid hypothesis. *Ann Acad Med Singap* **40**, 304-306 (2011).

85 Armstrong, R. A. The pathogenesis of Alzheimer's disease: a reevaluation of the "amyloid cascade hypothesis". *Int J Alzheimers Dis* **2011**, 630865, doi:10.4061/2011/630865 (2011).

86 Tabner, B. J., Mayes, J. & Allsop, D. Hypothesis: soluble aβ oligomers in association with redox-active metal ions are the optimal generators of reactive oxygen species in Alzheimer's disease. *Int J Alzheimers Dis* **2011**, 546380, doi:10.4061/2011/546380 (2010).

87 Sugimoto, H. [Development of anti-Alzheimer's disease drug based on beta-amyloid hypothesis]. *Yakugaku Zasshi* **130**, 521-526, doi:10.1248/yakushi.130.521 (2010).

88 Simón, A. M., Frechilla, D. & del Río, J. [Perspectives on the amyloid cascade hypothesis of Alzheimer's disease]. *Rev Neurol* **50**, 667-675 (2010).

89 Saxena, U. Alzheimer's disease amyloid hypothesis at crossroads: where do we go from here? *Expert Opin Ther Targets* **14**, 1273-1277, doi:10.1517/14728222.2010.528285 (2010).

90 Gandy, S. Testing the amyloid hypothesis of Alzheimer's disease in vivo. *Lancet Neurol* **9**, 333-335, doi:10.1016/s1474-4422(10)70055-7 (2010).

91 Ethell, D. W. An amyloid-notch hypothesis for Alzheimer's disease. *Neuroscientist* **16**, 614-617, doi:10.1177/1073858410366162 (2010).

92 Coomaraswamy, J. *et al.* Modeling familial Danish dementia in mice supports the concept of the amyloid hypothesis of Alzheimer's disease. *Proc Natl Acad Sci U S A* **107**, 7969-7974, doi:10.1073/pnas.1001056107 (2010).

93 Pimplikar, S. W. Reassessing the amyloid cascade hypothesis of Alzheimer's disease. *Int J Biochem Cell Biol* **41**, 1261-1268, doi:10.1016/j.biocel.2008.12.015 (2009).

94 Kreft, A. F., Martone, R. & Porte, A. Recent advances in the identification of gamma-secretase inhibitors to clinically test the Abeta oligomer hypothesis of Alzheimer's disease. *J Med Chem* **52**, 6169-6188, doi:10.1021/jm900188z (2009).

95 Jaeger, L. B. *et al.* Testing the neurovascular hypothesis of Alzheimer's disease: LRP-1 antisense reduces blood-brain barrier clearance, increases brain levels of amyloid-beta protein, and impairs cognition. *J Alzheimers Dis* **17**, 553-570, doi:10.3233/jad-2009-1074 (2009).

96 Hardy, J. The amyloid hypothesis for Alzheimer's disease: a critical reappraisal. *J Neurochem* **110**, 1129-1134, doi:10.1111/j.1471-4159.2009.06181.x (2009).

97 Sugimoto, H. [New approaches for the development of anti-Alzheimer's disease drugs based on the cholinergic hypothesis and amyloid hypothesis]. *Nihon Yakurigaku Zasshi* **131**, 338-340, doi:10.1254/fpj.131.338 (2008).

98 Small, S. A. & Duff, K. Linking Abeta and tau in late-onset Alzheimer's disease: a dual pathway hypothesis. *Neuron* **60**, 534-542, doi:10.1016/j.neuron.2008.11.007 (2008).
99 Shirwany, N. A., Payette, D., Xie, J. & Guo, Q. The amyloid beta ion channel hypothesis of Alzheimer's disease. *Neuropsychiatr Dis Treat* **3**, 597-612 (2007).
100 Schmitt, H. P. epsilon-Glycation, APP and Abeta in ageing and Alzheimer disease: a hypothesis. *Med Hypotheses* **66**, 898-906, doi:10.1016/j.mehy.2005.11.016 (2006).
101 Qiu, W. Q. & Folstein, M. F. Insulin, insulin-degrading enzyme and amyloid-beta peptide in Alzheimer's disease: review and hypothesis. *Neurobiol Aging* **27**, 190-198, doi:10.1016/j.neurobiolaging.2005.01.004 (2006).
102 Oprisiu, R., Serot, J. M., Godefroy, O., Black, S. E. & Fournier, A. Plasma amyloid-beta concentrations in Alzheimer's disease: an alternative hypothesis. *Lancet Neurol* **5**, 1001-1002; author reply 1002-1003, doi:10.1016/s1474-4422(06)70612-3 (2006).
103 Nguyen, J. T., Yamani, A. & Kiso, Y. Views on amyloid hypothesis and secretase inhibitors for treating Alzheimer's disease: progress and problems. *Curr Pharm Des* **12**, 4295-4312, doi:10.2174/138161206778792976 (2006).
104 Hardy, J. Alzheimer's disease: the amyloid cascade hypothesis: an update and reappraisal. *J Alzheimers Dis* **9**, 151-153, doi:10.3233/jad-2006-9s317 (2006).
105 Hardy, J. Has the amyloid cascade hypothesis for Alzheimer's disease been proved? *Curr Alzheimer Res* **3**, 71-73, doi:10.2174/156720506775697098 (2006).
106 Golde, T. E., Dickson, D. & Hutton, M. Filling the gaps in the abeta cascade hypothesis of Alzheimer's disease. *Curr Alzheimer Res* **3**, 421-430, doi:10.2174/156720506779025189 (2006).
107 Fullwood, N. J., Hayashi, Y. & Allsop, D. Plasma amyloid-beta concentrations in Alzheimer's disease: an alternative hypothesis. *Lancet Neurol* **5**, 1000-1001; author reply 1002-1003, doi:10.1016/s1474-4422(06)70611-1 (2006).
108 Fagan, T. *et al.* Alzheimer Research Forum Live Discussion: Now you see them, now you don't: The amyloid channel hypothesis. *J Alzheimers Dis* **9**, 219-224, doi:10.3233/jad-2006-9213 (2006).
109 Tanzi, R. E. & Bertram, L. Twenty years of the Alzheimer's disease amyloid hypothesis: a genetic perspective. *Cell* **120**, 545-555, doi:10.1016/j.cell.2005.02.008 (2005).
110 Tanzi, R. E. The synaptic Abeta hypothesis of Alzheimer disease. *Nat Neurosci* **8**, 977-979, doi:10.1038/nn0805-977 (2005).
111 Roy, S. & Rauk, A. Alzheimer's disease and the 'ABSENT' hypothesis: mechanism for amyloid beta endothelial and neuronal toxicity. *Med Hypotheses* **65**, 123-137, doi:10.1016/j.mehy.2004.08.031 (2005).
112 Marchesi, V. T. An alternative interpretation of the amyloid Abeta hypothesis with regard to the pathogenesis of Alzheimer's disease. *Proc Natl Acad Sci U S A* **102**, 9093-9098, doi:10.1073/pnas.0503181102 (2005).
113 Golde, T. E. The Abeta hypothesis: leading us to rationally-designed therapeutic strategies for the treatment or prevention of Alzheimer disease. *Brain Pathol* **15**, 84-87, doi:10.1111/j.1750-3639.2005.tb00104.x (2005).
114 Exley, C. The aluminium-amyloid cascade hypothesis and Alzheimer's disease. *Subcell Biochem* **38**, 225-234, doi:10.1007/0-387-23226-5_11 (2005).

115 LeVine, H., 3rd. The Amyloid Hypothesis and the clearance and degradation of Alzheimer's beta-peptide. *J Alzheimers Dis* **6**, 303-314, doi:10.3233/jad-2004-6311 (2004).
116 Lee, H. G. *et al.* Challenging the amyloid cascade hypothesis: senile plaques and amyloid-beta as protective adaptations to Alzheimer disease. *Ann N Y Acad Sci* **1019**, 1-4, doi:10.1196/annals.1297.001 (2004).
117 Kowalska, A. [The beta-amyloid cascade hypothesis: a sequence of events leading to neurodegeneration in Alzheimer's disease]. *Neurol Neurochir Pol* **38**, 405-411 (2004).
118 Gutierrez-Zepeda, A. & Luo, Y. Testing the amyloid toxicity hypothesis of Alzheimer's disease in transgenic Caenorhabditis elegans model. *Front Biosci* **9**, 3333-3338, doi:10.2741/1485 (2004).
119 Sommer, B. Alzheimer's disease and the amyloid cascade hypothesis: ten years on. *Curr Opin Pharmacol* **2**, 87-92, doi:10.1016/s1471-4892(01)00126-6 (2002).
120 Rosenblum, W. I. Structure and location of amyloid beta peptide chains and arrays in Alzheimer's disease: new findings require reevaluation of the amyloid hypothesis and of tests of the hypothesis. *Neurobiol Aging* **23**, 225-230, doi:10.1016/s0197-4580(01)00283-4 (2002).
121 Robinson, S. R. & Bishop, G. M. Abeta as a bioflocculant: implications for the amyloid hypothesis of Alzheimer's disease. *Neurobiol Aging* **23**, 1051-1072, doi:10.1016/s0197-4580(01)00342-6 (2002).
122 Morgan, D. & Keller, R. K. What evidence would prove the amyloid hypothesis? Towards rational drug treatments for Alzheimer's disease. *J Alzheimers Dis* **4**, 257-260, doi:10.3233/jad-2002-4317 (2002).
123 Hashimoto, K. & Iyo, M. [Amyloid cascade hypothesis of Alzheimer's disease and alpha 7 nicotinic receptor]. *Nihon Shinkei Seishin Yakurigaku Zasshi* **22**, 49-53 (2002).
124 Hardy, J. & Selkoe, D. J. The amyloid hypothesis of Alzheimer's disease: progress and problems on the road to therapeutics. *Science* **297**, 353-356, doi:10.1126/science.1072994 (2002).
125 Dominguez, D. I. & De Strooper, B. Novel therapeutic strategies provide the real test for the amyloid hypothesis of Alzheimer's disease. *Trends Pharmacol Sci* **23**, 324-330, doi:10.1016/s0165-6147(02)02038-2 (2002).
126 Gerlai, R. Alzheimer's disease: beta-amyloid hypothesis strengthened! *Trends Neurosci* **24**, 199, doi:10.1016/s0166-2236(00)01799-9 (2001).
127 Fletcher, L. Vaccine tests key Alzheimer's disease hypothesis. *Nat Biotechnol* **19**, 104-105, doi:10.1038/84340 (2001).
128 Selkoe, D. J. Toward a comprehensive theory for Alzheimer's disease. Hypothesis: Alzheimer's disease is caused by the cerebral accumulation and cytotoxicity of amyloid beta-protein. *Ann N Y Acad Sci* **924**, 17-25, doi:10.1111/j.1749-6632.2000.tb05554.x (2000).
129 Lovestone, S. Fleshing out the amyloid cascade hypothesis: the molecular biology of Alzheimer's disease. *Dialogues Clin Neurosci* **2**, 101-110, doi:10.31887/DCNS.2000.2.2/slovestone (2000).
130 Butcher, J. Alzheimer's amyloid hypothesis gains support. *Lancet* **356**, 2161, doi:10.1016/s0140-6736(00)03504-2 (2000).

131 Yatin, S. M., Aksenov, M. & Butterfield, D. A. The antioxidant vitamin E modulates amyloid beta-peptide-induced creatine kinase activity inhibition and increased protein oxidation: implications for the free radical hypothesis of Alzheimer's disease. *Neurochem Res* **24**, 427-435, doi:10.1023/a:1020997903147 (1999).
132 Teplow, D. B. Truncating the amyloid cascade hypothesis: the role of C-terminal Abeta peptides in Alzheimer's disease. *Neurobiol Aging* **20**, 71-73; discussion 87, doi:10.1016/s0197-4580(99)00013-5 (1999).
133 Larner, A. J. Hypothesis: amyloid beta-peptides truncated at the N-terminus contribute to the pathogenesis of Alzheimer's disease. *Neurobiol Aging* **20**, 65-69, doi:10.1016/s0197-4580(99)00014-7 (1999).
134 Subramaniam, R. *et al.* The free radical antioxidant vitamin E protects cortical synaptosomal membranes from amyloid beta-peptide(25-35) toxicity but not from hydroxynonenal toxicity: relevance to the free radical hypothesis of Alzheimer's disease. *Neurochem Res* **23**, 1403-1410, doi:10.1023/a:1020754807671 (1998).
135 Small, D. H. The Sixth International Conference on Alzheimer's disease, Amsterdam, The Netherlands, July 1998. The amyloid cascade hypothesis debate: emerging consensus on the role of A beta and amyloid in Alzheimer's disease. *Amyloid* **5**, 301-304, doi:10.3109/13506129809007304 (1998).
136 Neve, R. L. & Robakis, N. K. Alzheimer's disease: a re-examination of the amyloid hypothesis. *Trends Neurosci* **21**, 15-19, doi:10.1016/s0166-2236(97)01168-5 (1998).
137 Swaab, D. F. & Salehi, A. The pathogenesis of Alzheimer disease: an alternative to the amyloid hypothesis. *J Neuropathol Exp Neurol* **56**, 216 (1997).
138 Smith, M. A. & Perry, G. The pathogenesis of Alzheimer disease: an alternative to the amyloid hypothesis. *J Neuropathol Exp Neurol* **56**, 217 (1997).
139 Schehr, R. S. Amyloid hypothesis of Alzheimer's rides high--for now. *Nat Biotechnol* **15**, 19-20, doi:10.1038/nbt0197-19 (1997).
140 Rosenblum, W. I. The pathogenesis of Alzheimer disease: an alternative to the amyloid hypothesis. *J Neuropathol Exp Neurol* **56**, 213 (1997).
141 Larner, A. J. The pathogenesis of Alzheimer disease: an alternative to the amyloid hypothesis. *J Neuropathol Exp Neurol* **56**, 214-215 (1997).
142 Ehrenstein, G., Galdzicki, Z. & Lange, G. D. The choline-leakage hypothesis for the loss of acetylcholine in Alzheimer's disease. *Biophys J* **73**, 1276-1280, doi:10.1016/s0006-3495(97)78160-8 (1997).
143 Terry, R. D. The pathogenesis of Alzheimer disease: an alternative to the amyloid hypothesis. *J Neuropathol Exp Neurol* **55**, 1023-1025 (1996).
144 Pollard, H. B., Arispe, N. & Rojas, E. Ion channel hypothesis for Alzheimer amyloid peptide neurotoxicity. *Cell Mol Neurobiol* **15**, 513-526, doi:10.1007/bf02071314 (1995).
145 Greenberg, B. D. & Murphy, M. F. Toward an integrated discovery and development program in Alzheimer's disease: the amyloid hypothesis. *Neurobiol Aging* **15 Suppl 2**, S105-109, doi:10.1016/0197-4580(94)90184-8 (1994).
146 Arispe, N., Pollard, H. B. & Rojas, E. beta-Amyloid Ca(2+)-channel hypothesis for neuronal death in Alzheimer disease. *Mol Cell Biochem* **140**, 119-125, doi:10.1007/bf00926750 (1994).

147 Roberts, G. W., Nash, M., Ince, P. G., Royston, M. C. & Gentleman, S. M. On the origin of Alzheimer's disease: a hypothesis. *Neuroreport* **4**, 7-9, doi:10.1097/00001756-199301000-00001 (1993).
148 Armstrong, R. A., Myers, D. & Smith, C. U. The spatial patterns of plaques and tangles in Alzheimer's disease do not support the 'cascade hypothesis'. *Dementia* **4**, 16-20, doi:10.1159/000107291 (1993).
149 Hardy, J. A. & Higgins, G. A. Alzheimer's disease: the amyloid cascade hypothesis. *Science* **256**, 184-185, doi:10.1126/science.1566067 (1992).
150 Constantinidis, J. Hypothesis regarding amyloid and zinc in the pathogenesis of Alzheimer disease: potential for preventive intervention. *Alzheimer Dis Assoc Disord* **5**, 31-35, doi:10.1097/00002093-199100510-00004 (1991).
151 Schweber, M. A possible unitary genetic hypothesis for Alzheimer's disease and Down syndrome. *Ann N Y Acad Sci* **450**, 223-238, doi:10.1111/j.1749-6632.1985.tb21495.x (1985).
152 Kawabata, S. Excessive/Aberrant and Maladaptive Synaptic Plasticity: A Hypothesis for the Pathogenesis of Alzheimer's Disease. *Front Aging Neurosci* **14**, 913693, doi:10.3389/fnagi.2022.913693 (2022).
153 Guzmán-Ramos, K., Osorio-Gómez, D. & Bermúdez-Rattoni, F. Cognitive Impairment in Alzheimer's and Metabolic Diseases: A Catecholaminergic Hypothesis. *Neuroscience* **497**, 308-323, doi:10.1016/j.neuroscience.2022.05.031 (2022).
154 Ovsepian, S. V., O'Leary, V. B., Hoschl, C. & Zaborszky, L. Integrated phylogeny of the human brain and pathobiology of Alzheimer's disease: A unifying hypothesis. *Neurosci Lett* **755**, 135895, doi:10.1016/j.neulet.2021.135895 (2021).
155 Nara, P. L., Sindelar, D., Penn, M. S., Potempa, J. & Griffin, W. S. T. Porphyromonas gingivalis Outer Membrane Vesicles as the Major Driver of and Explanation for Neuropathogenesis, the Cholinergic Hypothesis, Iron Dyshomeostasis, and Salivary Lactoferrin in Alzheimer's Disease. *J Alzheimers Dis* **82**, 1417-1450, doi:10.3233/jad-210448 (2021).
156 Mondragón-Rodríguez, S., Salgado-Burgos, H. & Peña-Ortega, F. Circuitry and Synaptic Dysfunction in Alzheimer's Disease: A New Tau Hypothesis. *Neural Plast* **2020**, 2960343, doi:10.1155/2020/2960343 (2020).
157 Krashia, P., Nobili, A. & D'Amelio, M. Unifying Hypothesis of Dopamine Neuron Loss in Neurodegenerative Diseases: Focusing on Alzheimer's Disease. *Front Mol Neurosci* **12**, 123, doi:10.3389/fnmol.2019.00123 (2019).
158 Hampel, H. *et al.* Revisiting the Cholinergic Hypothesis in Alzheimer's Disease: Emerging Evidence from Translational and Clinical Research. *J Prev Alzheimers Dis* **6**, 2-15, doi:10.14283/jpad.2018.43 (2019).
159 Lee, J. *et al.* Sex-Related Reserve Hypothesis in Alzheimer's Disease: Changes in Cortical Thickness with a Five-Year Longitudinal Follow-Up. *J Alzheimers Dis* **65**, 641-649, doi:10.3233/jad-180049 (2018).
160 Vakalopoulos, C. Alzheimer's Disease: The Alternative Serotonergic Hypothesis of Cognitive Decline. *J Alzheimers Dis* **60**, 859-866, doi:10.3233/jad-170364 (2017).

161 Persson, K. *et al.* MRI-assessed atrophy subtypes in Alzheimer's disease and the cognitive reserve hypothesis. *PLoS One* **12**, e0186595, doi:10.1371/journal.pone.0186595 (2017).
162 Hampel, H. *et al.* WITHDRAWN: Revisiting the cholinergic hypothesis in Alzheimer's disease: Emerging evidence from translational and clinical research. *Alzheimers Dement*, doi:10.1016/j.jalz.2017.08.016 (2017).
163 Lacalle-Aurioles, M. *et al.* The Disconnection Hypothesis in Alzheimer's Disease Studied Through Multimodal Magnetic Resonance Imaging: Structural, Perfusion, and Diffusion Tensor Imaging. *J Alzheimers Dis* **50**, 1051-1064, doi:10.3233/jad-150288 (2016).
164 Konishi, K. *et al.* Hypothesis of Endogenous Anticholinergic Activity in Alzheimer's Disease. *Neurodegener Dis* **15**, 149-156, doi:10.1159/000381511 (2015).
165 Hachisu, M. *et al.* Beyond the Hypothesis of Serum Anticholinergic Activity in Alzheimer's Disease: Acetylcholine Neuronal Activity Modulates Brain-Derived Neurotrophic Factor Production and Inflammation in the Brain. *Neurodegener Dis* **15**, 182-187, doi:10.1159/000381531 (2015).
166 Fotiou, D., Kaltsatou, A., Tsiptsios, D. & Nakou, M. Evaluation of the cholinergic hypothesis in Alzheimer's disease with neuropsychological methods. *Aging Clin Exp Res* **27**, 727-733, doi:10.1007/s40520-015-0321-8 (2015).
167 Ashford, J. W. Treatment of Alzheimer's Disease: The Legacy of the Cholinergic Hypothesis, Neuroplasticity, and Future Directions. *J Alzheimers Dis* **47**, 149-156, doi:10.3233/jad-150381 (2015).
168 Popugaeva, E. & Bezprozvanny, I. Can the calcium hypothesis explain synaptic loss in Alzheimer's disease? *Neurodegener Dis* **13**, 139-141, doi:10.1159/000354778 (2014).
169 Cochran, J. N., Hall, A. M. & Roberson, E. D. The dendritic hypothesis for Alzheimer's disease pathophysiology. *Brain Res Bull* **103**, 18-28, doi:10.1016/j.brainresbull.2013.12.004 (2014).
170 Brier, M. R., Thomas, J. B. & Ances, B. M. Network dysfunction in Alzheimer's disease: refining the disconnection hypothesis. *Brain Connect* **4**, 299-311, doi:10.1089/brain.2014.0236 (2014).
171 Shimohama, S. [Development of therapies for Alzheimer's disease based on cholinergic hypothesis-status quo and future directions]. *Rinsho Shinkeigaku* **53**, 1036-1038, doi:10.5692/clinicalneurol.53.1036 (2013).
172 Hori, K. *et al.* [Proposal of endogenous anticholinergic hypothesis in Alzheimer disease]. *Nihon Shinkei Seishin Yakurigaku Zasshi* **33**, 117-126 (2013).
173 Daulatzai, M. A. Dysfunctional nucleus tractus solitarius: its crucial role in promoting neuropathogenetic cascade of Alzheimer's dementia--a novel hypothesis. *Neurochem Res* **37**, 846-868, doi:10.1007/s11064-011-0680-2 (2012).
174 Cattaneo, A. & Calissano, P. Nerve growth factor and Alzheimer's disease: new facts for an old hypothesis. *Mol Neurobiol* **46**, 588-604, doi:10.1007/s12035-012-8310-9 (2012).
175 Pinto, T., Lanctôt, K. L. & Herrmann, N. Revisiting the cholinergic hypothesis of behavioral and psychological symptoms in dementia of the Alzheimer's type. *Ageing Res Rev* **10**, 404-412, doi:10.1016/j.arr.2011.01.003 (2011).

176 Craig, L. A., Hong, N. S. & McDonald, R. J. Revisiting the cholinergic hypothesis in the development of Alzheimer's disease. *Neurosci Biobehav Rev* **35**, 1397-1409, doi:10.1016/j.neubiorev.2011.03.001 (2011).

177 Tabaton, M. *et al.* Memantine: "hypothesis testing" not "disease modifying" in Alzheimer's disease. *Am J Pathol* **176**, 540-541, doi:10.2353/ajpath.2010.090856 (2010).

178 Scharre, D. W. *et al.* Use of antipsychotic drugs in patients with Alzheimer's disease treated with rivastigmine versus donepezil: a retrospective, parallel-cohort, hypothesis-generating study. *Drugs Aging* **27**, 903-913, doi:10.2165/11584290-000000000-00000 (2010).

179 Martorana, A., Esposito, Z. & Koch, G. Beyond the cholinergic hypothesis: do current drugs work in Alzheimer's disease? *CNS Neurosci Ther* **16**, 235-245, doi:10.1111/j.1755-5949.2010.00175.x (2010).

180 Wu, T. Y. & Chen, C. P. Dual action of memantine in Alzheimer disease: a hypothesis. *Taiwan J Obstet Gynecol* **48**, 273-277, doi:10.1016/s1028-4559(09)60303-x (2009).

181 Zhu, Y. B., Lu, P. H. & Sheng, Z. H. [A new hypothesis for early pathogenesis in Alzheimer's disease: impaired axonal transport mechanism]. *Sheng Li Ke Xue Jin Zhan* **39**, 5-9 (2008).

182 Sugimoto, H. The new approach in development of anti-Alzheimer's disease drugs via the cholinergic hypothesis. *Chem Biol Interact* **175**, 204-208, doi:10.1016/j.cbi.2008.05.031 (2008).

183 Brousseau, G., Rourke, B. P. & Burke, B. Acetylcholinesterase inhibitors, neuropsychiatric symptoms, and Alzheimer's disease subtypes: an alternate hypothesis to global cognitive enhancement. *Exp Clin Psychopharmacol* **15**, 546-554, doi:10.1037/1064-1297.15.6.546 (2007).

184 Sivaprakasam, K. Towards a unifying hypothesis of Alzheimer's disease: cholinergic system linked to plaques, tangles and neuroinflammation. *Curr Med Chem* **13**, 2179-2188, doi:10.2174/092986706777935203 (2006).

185 Claassen, J. A. & Jansen, R. W. Cholinergically mediated augmentation of cerebral perfusion in Alzheimer's disease and related cognitive disorders: the cholinergic-vascular hypothesis. *J Gerontol A Biol Sci Med Sci* **61**, 267-271, doi:10.1093/gerona/61.3.267 (2006).

186 Axonal transport hypothesis moves on to implicate presenilin: Alzheimer research forum live discussion. *J Alzheimers Dis* **6**, 537-545, doi:10.3233/jad-2004-6511 (2004).

187 Terry, A. V., Jr. & Buccafusco, J. J. The cholinergic hypothesis of age and Alzheimer's disease-related cognitive deficits: recent challenges and their implications for novel drug development. *J Pharmacol Exp Ther* **306**, 821-827, doi:10.1124/jpet.102.041616 (2003).

188 Tatton, W. *et al.* Hypothesis for a common basis for neuroprotection in glaucoma and Alzheimer's disease: anti-apoptosis by alpha-2-adrenergic receptor activation. *Surv Ophthalmol* **48 Suppl 1**, S25-37, doi:10.1016/s0039-6257(03)00005-5 (2003).

189 Scarmeas, N. *et al.* Association of life activities with cerebral blood flow in Alzheimer disease: implications for the cognitive reserve hypothesis. *Arch Neurol* **60**, 359-365, doi:10.1001/archneur.60.3.359 (2003).

190 White, K. G. & Ruske, A. C. Memory deficits in Alzheimer's disease: the encoding hypothesis and cholinergic function. *Psychon Bull Rev* **9**, 426-437, doi:10.3758/bf03196301 (2002).
191 Morris, J. C. Challenging assumptions about Alzheimer's disease: mild cognitive impairment and the cholinergic hypothesis. *Ann Neurol* **51**, 143-144, doi:10.1002/ana.10135 (2002).
192 Kagan, B. L., Hirakura, Y., Azimov, R., Azimova, R. & Lin, M. C. The channel hypothesis of Alzheimer's disease: current status. *Peptides* **23**, 1311-1315, doi:10.1016/s0196-9781(02)00067-0 (2002).
193 Bigio, E. H., Hynan, L. S., Sontag, E., Satumtira, S. & White, C. L. Synapse loss is greater in presenile than senile onset Alzheimer disease: implications for the cognitive reserve hypothesis. *Neuropathol Appl Neurobiol* **28**, 218-227, doi:10.1046/j.1365-2990.2002.00385.x (2002).
194 Wevers, A. *et al.* Classical Alzheimer features and cholinergic dysfunction: towards a unifying hypothesis? *Acta Neurol Scand Suppl* **176**, 42-48, doi:10.1034/j.1600-0404.2000.00306.x (2000).
195 Jenkins, R., Fox, N. C., Rossor, A. M., Harvey, R. J. & Rossor, M. N. Intracranial volume and Alzheimer disease: evidence against the cerebral reserve hypothesis. *Arch Neurol* **57**, 220-224, doi:10.1001/archneur.57.2.220 (2000).
196 Danysz, W., Parsons, C. G., Mobius, H. J., Stoffler, A. & Quack, G. Neuroprotective and symptomatological action of memantine relevant for Alzheimer's disease--a unified glutamatergic hypothesis on the mechanism of action. *Neurotox Res* **2**, 85-97, doi:10.1007/bf03033787 (2000).
197 Robert, P. H., Gokalsing, E. & Bertogliati, C. [Cholinergic hypothesis and Alzheimer's disease: the place of donepezil (Aricept)]. *Encephale* **25 Spec No 5**, 23-27; discussion 28-29 (1999).
198 Francis, P. T., Palmer, A. M., Snape, M. & Wilcock, G. K. The cholinergic hypothesis of Alzheimer's disease: a review of progress. *J Neurol Neurosurg Psychiatry* **66**, 137-147, doi:10.1136/jnnp.66.2.137 (1999).
199 Davies, P. Challenging the cholinergic hypothesis in Alzheimer disease. *Jama* **281**, 1433-1434, doi:10.1001/jama.281.15.1433 (1999).
200 Babic, T. The cholinergic hypothesis of Alzheimer's disease: a review of progress. *J Neurol Neurosurg Psychiatry* **67**, 558, doi:10.1136/jnnp.67.4.558 (1999).
201 Marczynski, T. J. GABAergic deafferentation hypothesis of brain aging and Alzheimer's disease revisited. *Brain Res Bull* **45**, 341-379, doi:10.1016/s0361-9230(97)00347-x (1998).
202 Ladner, C. J. & Lee, J. M. Pharmacological drug treatment of Alzheimer disease: the cholinergic hypothesis revisited. *J Neuropathol Exp Neurol* **57**, 719-731, doi:10.1097/00005072-199808000-00001 (1998).
203 Kaufer, D. Beyond the cholinergic hypothesis: the effect of metrifonate and other cholinesterase inhibitors on neuropsychiatric symptoms in Alzheimer's disease. *Dement Geriatr Cogn Disord* **9 Suppl 2**, 8-14, doi:10.1159/000051193 (1998).

204 Fisher, A. *et al.* Novel m1 muscarinic agonists in treatment and delaying the progression of Alzheimer's disease: an unifying hypothesis. *J Physiol Paris* **92**, 337-340, doi:10.1016/s0928-4257(99)80001-1 (1998).
205 Cummings, J. L. & Back, C. The cholinergic hypothesis of neuropsychiatric symptoms in Alzheimer's disease. *Am J Geriatr Psychiatry* **6**, S64-78, doi:10.1097/00019442-199821001-00009 (1998).
206 Robbins, T. W., McAlonan, G., Muir, J. L. & Everitt, B. J. Cognitive enhancers in theory and practice: studies of the cholinergic hypothesis of cognitive deficits in Alzheimer's disease. *Behav Brain Res* **83**, 15-23, doi:10.1016/s0166-4328(97)86040-8 (1997).
207 Olney, J. W., Wozniak, D. F. & Farber, N. B. Excitotoxic neurodegeneration in Alzheimer disease. New hypothesis and new therapeutic strategies. *Arch Neurol* **54**, 1234-1240, doi:10.1001/archneur.1997.00550220042012 (1997).
208 Ying, W. Deleterious network hypothesis of Alzheimer's disease. *Med Hypotheses* **46**, 421-428, doi:10.1016/s0306-9877(96)90021-3 (1996).
209 Cummings, J. L. & Kaufer, D. Neuropsychiatric aspects of Alzheimer's disease: the cholinergic hypothesis revisited. *Neurology* **47**, 876-883, doi:10.1212/wnl.47.4.876 (1996).
210 Weinstock, M. The pharmacotherapy of Alzheimer's disease based on the cholinergic hypothesis: an update. *Neurodegeneration* **4**, 349-356, doi:10.1006/neur.1995.0042 (1995).
211 Neill, D. Alzheimer's disease: maladaptive synaptoplasticity hypothesis. *Neurodegeneration* **4**, 217-232, doi:10.1006/neur.1995.0027 (1995).
212 Marczynski, T. J. GABAergic deafferentation hypothesis of brain aging and Alzheimer's disease; pharmacologic profile of the benzodiazepine antagonist, flumazenil. *Rev Neurosci* **6**, 221-258, doi:10.1515/revneuro.1995.6.3.221 (1995).
213 Gualtieri, F. *et al.* The medicinal chemistry of Alzheimer's and Alzheimer-like diseases with emphasis on the cholinergic hypothesis. *Farmaco* **50**, 489-503 (1995).
214 Ingram, D. K. *et al.* Rodent models of memory dysfunction in Alzheimer's disease and normal aging: moving beyond the cholinergic hypothesis. *Life Sci* **55**, 2037-2049, doi:10.1016/0024-3205(94)00384-x (1994).
215 Förstl, H., Levy, R., Burns, A., Luthert, P. & Cairns, N. Disproportionate loss of noradrenergic and cholinergic neurons as cause of depression in Alzheimer's disease--a hypothesis. *Pharmacopsychiatry* **27**, 11-15, doi:10.1055/s-2007-1014267 (1994).
216 Aarsland, D., Larsen, J. P., Reinvang, I. & Aasland, A. M. Effects of cholinergic blockade on language in healthy young women. Implications for the cholinergic hypothesis in dementia of the Alzheimer type. *Brain* **117 ( Pt 6)**, 1377-1384, doi:10.1093/brain/117.6.1377 (1994).
217 Nordberg, A. Biological markers and the cholinergic hypothesis in Alzheimer's disease. *Acta Neurol Scand Suppl* **139**, 54-58, doi:10.1111/j.1600-0404.1992.tb04455.x (1992).
218 Horn, D. & Ruppin, E. Extra-pyramidal symptoms in Alzheimer's disease: a hypothesis. *Med Hypotheses* **39**, 316-318, doi:10.1016/0306-9877(92)90055-h (1992).
219 Pascual, J. *et al.* High-affinity choline uptake carrier in Alzheimer's disease: implications for the cholinergic hypothesis of dementia. *Brain Res* **552**, 170-174, doi:10.1016/0006-8993(91)90676-m (1991).

220 Vamvakides, A. [Working hypothesis for the pharmacologic research in the glutamatergic approach to Alzheimer's disease]. *Ann Pharm Fr* **47**, 329-336 (1989).
221 Masliah, E., Terry, R. & Buzsáki, G. Thalamic nuclei in Alzheimer disease: evidence against the cholinergic hypothesis of plaque formation. *Brain Res* **493**, 240-246, doi:10.1016/0006-8993(89)91159-1 (1989).
222 Kish, S. J. *et al.* Cognitive deficits in olivopontocerebellar atrophy: implications for the cholinergic hypothesis of Alzheimer's dementia. *Ann Neurol* **24**, 200-206, doi:10.1002/ana.410240205 (1988).
223 Chan-Palay, V. Galanin hyperinnervates surviving neurons of the human basal nucleus of Meynert in dementias of Alzheimer's and Parkinson's disease: a hypothesis for the role of galanin in accentuating cholinergic dysfunction in dementia. *J Comp Neurol* **273**, 543-557, doi:10.1002/cne.902730409 (1988).
224 Adams, P. R. Cholinergic hypothesis of Alzheimer's disease: biophysical aspects. *Res Publ Assoc Res Nerv Ment Dis* **65**, 169-185 (1987).
225 Tucker, L. Tay-Sachs disease, Alzheimer's disease and the cholinergic hypothesis. *S Afr Med J* **70**, 847 (1986).
226 Pomara, N. & Stanley, M. The cholinergic hypothesis of memory dysfunction in Alzheimer's disease--revisited. *Psychopharmacol Bull* **22**, 110-118 (1986).
227 Harrison, P. J. Pathogenesis of Alzheimer's disease--beyond the cholinergic hypothesis: discussion paper. *J R Soc Med* **79**, 347-352, doi:10.1177/014107688607900612 (1986).
228 Coralli, M. V., Zanotti, E. & Salsi, A. [Current concepts on the hypothesis of the cholinergic etiology of Alzheimer's disease ]. *Recenti Prog Med* **77**, 436-439 (1986).
229 Wurtman, R. J., Blusztajn, J. K. & Maire, J. C. "Autocannibalism" of choline-containing membrane phospholipids in the pathogenesis of Alzheimer's disease-A hypothesis. *Neurochem Int* **7**, 369-372, doi:10.1016/0197-0186(85)90127-5 (1985).
230 Rossor, M. N. Focal changes in Alzheimer's disease and cholinergic hypothesis. *Lancet* **2**, 465, doi:10.1016/s0140-6736(83)90441-5 (1983).
231 Jonveaux, T. R. & Fescharek, R. When Art Meets Gardens: Does It Enhance the Benefits? The Nancy Hypothesis of Care for Persons with Alzheimer's Disease. *J Alzheimers Dis* **61**, 885-898, doi:10.3233/jad-170781 (2018).
232 Doig, A. J. Positive Feedback Loops in Alzheimer's Disease: The Alzheimer's Feedback Hypothesis. *J Alzheimers Dis* **66**, 25-36, doi:10.3233/jad-180583 (2018).
233 Fliss, R. *et al.* Theory of Mind and social reserve: Alternative hypothesis of progressive Theory of Mind decay during different stages of Alzheimer's disease. *Soc Neurosci* **11**, 409-423, doi:10.1080/17470919.2015.1101014 (2016).
234 Serino, S. & Riva, G. What is the role of spatial processing in the decline of episodic memory in Alzheimer's disease? The "mental frame syncing" hypothesis. *Front Aging Neurosci* **6**, 33, doi:10.3389/fnagi.2014.00033 (2014).
235 Clark, C. N. & Warren, J. D. A hypnic hypothesis of Alzheimer's disease. *Neurodegener Dis* **12**, 165-176, doi:10.1159/000350060 (2013).
236 Liu, Y. H. & Tian, T. Hypothesis of optineurin as a new common risk factor in normal-tension glaucoma and Alzheimer's disease. *Med Hypotheses* **77**, 591-592, doi:10.1016/j.mehy.2011.06.040 (2011).

237 Giaccone, G., Orsi, L., Cupidi, C. & Tagliavini, F. Lipofuscin hypothesis of Alzheimer's disease. *Dement Geriatr Cogn Dis Extra* **1**, 292-296, doi:10.1159/000329544 (2011).
238 Rubey, R. N. Could lysine supplementation prevent Alzheimer's dementia? A novel hypothesis. *Neuropsychiatr Dis Treat* **6**, 707-710, doi:10.2147/ndt.S14338 (2010).
239 Bilotta, F. *et al.* Postoperative cognitive dysfunction: toward the Alzheimer's disease pathomechanism hypothesis. *J Alzheimers Dis* **22 Suppl 3**, 81-89, doi:10.3233/jad-2010-100825 (2010).
240 Modrego, P. J. Depression in the prodromal phase of Alzheimer disease and the reverse causal hypothesis. *Arch Gen Psychiatry* **66**, 107, doi:10.1001/archgenpsychiatry.2008.502 (2009).
241 Brito, G. N. Exercise and cognitive function: a hypothesis for the association of type II diabetes mellitus and Alzheimer's disease from an evolutionary perspective. *Diabetol Metab Syndr* **1**, 7, doi:10.1186/1758-5996-1-7 (2009).
242 Kemppainen, N. M. *et al.* Cognitive reserve hypothesis: Pittsburgh Compound B and fluorodeoxyglucose positron emission tomography in relation to education in mild Alzheimer's disease. *Ann Neurol* **63**, 112-118, doi:10.1002/ana.21212 (2008).
243 Roe, C. M., Xiong, C., Miller, J. P. & Morris, J. C. Education and Alzheimer disease without dementia: support for the cognitive reserve hypothesis. *Neurology* **68**, 223-228, doi:10.1212/01.wnl.0000251303.50459.8a (2007).
244 Alkon, D. L., Sun, M. K. & Nelson, T. J. PKC signaling deficits: a mechanistic hypothesis for the origins of Alzheimer's disease. *Trends Pharmacol Sci* **28**, 51-60, doi:10.1016/j.tips.2006.12.002 (2007).
245 Wostyn, P. Normal-tension glaucoma and Alzheimer's disease: hypothesis of a possible common underlying risk factor. *Med Hypotheses* **67**, 1255-1256, doi:10.1016/j.mehy.2006.05.012 (2006).
246 Nagy, Z. The last neuronal division: a unifying hypothesis for the pathogenesis of Alzheimer's disease. *J Cell Mol Med* **9**, 531-541, doi:10.1111/j.1582-4934.2005.tb00485.x (2005).
247 Malyshev, I. Y. *et al.* Possible use of adaptation to hypoxia in Alzheimer's disease: a hypothesis. *Med Sci Monit* **11**, Hy31-38 (2005).
248 Chakravarty, A. Unifying concept for Alzheimer's disease, vascular dementia and normal pressure hydrocephalus - a hypothesis. *Med Hypotheses* **63**, 827-833, doi:10.1016/j.mehy.2004.03.029 (2004).
249 Bateman, G. A. Pulse wave encephalopathy: a spectrum hypothesis incorporating Alzheimer's disease, vascular dementia and normal pressure hydrocephalus. *Med Hypotheses* **62**, 182-187, doi:10.1016/s0306-9877(03)00330-x (2004).
250 Wang, D. S. From Lewy body disease to Alzheimer's disease: hypothesis and evidence. *Front Biosci* **8**, s223-227, doi:10.2741/1029 (2003).
251 Silverberg, G. D., Mayo, M., Saul, T., Rubenstein, E. & McGuire, D. Alzheimer's disease, normal-pressure hydrocephalus, and senescent changes in CSF circulatory physiology: a hypothesis. *Lancet Neurol* **2**, 506-511, doi:10.1016/s1474-4422(03)00487-3 (2003).
252 Ji, M., Xiong, C. & Grundman, M. Hypothesis testing of a change point during cognitive decline among Alzheimer's disease patients. *J Alzheimers Dis* **5**, 375-382, doi:10.3233/jad-2003-5504 (2003).

253 Chan, D. K. A new hypothesis (concept) of diagnosing Alzheimer's disease. *J Gerontol A Biol Sci Med Sci* **57**, M645-647, doi:10.1093/gerona/57.10.m645 (2002).

254 Rasgon, N., Jarvik, G. P. & Jarvik, L. Affective disorders and Alzheimer disease: a missing-link hypothesis. *Am J Geriatr Psychiatry* **9**, 444-445 (2001).

255 Heininger, K. A unifying hypothesis of Alzheimer's disease. III. Risk factors. *Hum Psychopharmacol* **15**, 1-70, doi:10.1002/(sici)1099-1077(200001)15:1<1::Aid-hup153>3.0.Co;2-1 (2000).

256 Heininger, K. A unifying hypothesis of Alzheimer's disease. IV. Causation and sequence of events. *Rev Neurosci* **11 Spec No**, 213-328, doi:10.1515/revneuro.2000.11.s1.213 (2000).

257 Milberg, W., McGlinchey-Berroth, R., Duncan, K. M. & Higgins, J. A. Alterations in the dynamics of semantic activation in Alzheimer's disease: evidence for the Gain/Decay hypothesis of a disorder of semantic memory. *J Int Neuropsychol Soc* **5**, 641-658, doi:10.1017/s1355617799577072 (1999).

258 Seiler, N. An ammonia hypothesis of Alzheimer disease. *Adv Exp Med Biol* **420**, 235-255, doi:10.1007/978-1-4615-5945-0_16 (1997).

259 Mozaz, M. J. & Morris, R. G. Identification of body parts in Alzheimer's disease: evidence for a body schema hypothesis. *Int J Neurosci* **89**, 207-216, doi:10.3109/00207459708988475 (1997).

260 Cox, B. D. & Whichelow, M. J. Smoking and Alzheimer's disease: an alternative hypothesis. *J Epidemiol Community Health* **51**, 579, doi:10.1136/jech.51.5.579 (1997).

261 Thome, J. *et al.* [New hypothesis on etiopathogenesis of Alzheimer syndrome. Advanced glycation end products (AGEs)]. *Nervenarzt* **67**, 924-929, doi:10.1007/s001150050073 (1996).

262 Harman, D. A hypothesis on the pathogenesis of Alzheimer's disease. *Ann N Y Acad Sci* **786**, 152-168, doi:10.1111/j.1749-6632.1996.tb39059.x (1996).

263 Wostyn, P. Intracranial pressure and Alzheimer's disease: a hypothesis. *Med Hypotheses* **43**, 219-222, doi:10.1016/0306-9877(94)90069-8 (1994).

264 Landfield, P. W. Nathan Shock Memorial Lecture 1990. The role of glucocorticoids in brain aging and Alzheimer's disease: an integrative physiological hypothesis. *Exp Gerontol* **29**, 3-11, doi:10.1016/0531-5565(94)90058-2 (1994).

265 Vécsei, L. Alzheimer's disease and somatostatin: a therapeutic hypothesis. *Biol Psychiatry* **34**, 673-675, doi:10.1016/0006-3223(93)90039-g (1993).

266 Hardy, J. An 'anatomical cascade hypothesis' for Alzheimer's disease. *Trends Neurosci* **15**, 200-201, doi:10.1016/0166-2236(92)90033-5 (1992).

267 Potter, H. Review and hypothesis: Alzheimer disease and Down syndrome--chromosome 21 nondisjunction may underlie both disorders. *Am J Hum Genet* **48**, 1192-1200 (1991).

268 Baddeley, A., Della Sala, S. & Spinnler, H. The two-component hypothesis of memory deficit in Alzheimer's disease. *J Clin Exp Neuropsychol* **13**, 372-380, doi:10.1080/01688639108401051 (1991).

269 Deshmukh, V. D. & Deshmukh, S. V. Stress-adaptation failure hypothesis of Alzheimer's disease. *Med Hypotheses* **32**, 293-295, doi:10.1016/0306-9877(90)90109-r (1990).

270 Rapoport, S. I. Hypothesis: Alzheimer's disease is a phylogenetic disease. *Med Hypotheses* **29**, 147-150, doi:10.1016/0306-9877(89)90185-0 (1989).

271 Rocca, W. A. The etiology of Alzheimer's disease: epidemiologic contributions with emphasis on the genetic hypothesis. *J Neural Transm Suppl* **24**, 3-12 (1987).
272 Gorenstein, C. A hypothesis concerning the role of endogenous colchicine-like factors in the etiology of Alzheimer's disease. *Med Hypotheses* **23**, 371-374, doi:10.1016/0306-9877(87)90057-0 (1987).
273 Reubi, J. C. & Palacios, J. Somatostatin and Alzheimer's disease: a hypothesis. *J Neurol* **233**, 370-372, doi:10.1007/bf00313925 (1986).
274 Koukolík, F. [Criticism of the hippocampal hypothesis of dementia in Alzheimer's disease]. *Cas Lek Cesk* **124**, 1497-1498 (1985).
275 Abalan, F. Alzheimer's disease and malnutrition: a new etiological hypothesis. *Med Hypotheses* **15**, 385-393, doi:10.1016/0306-9877(84)90154-3 (1984).
276 Appel, S. H. A unifying hypothesis for the cause of amyotrophic lateral sclerosis, parkinsonism, and Alzheimer disease. *Ann Neurol* **10**, 499-505, doi:10.1002/ana.410100602 (1981).
277 Tortora, F. *et al.* CD33 rs2455069 SNP: Correlation with Alzheimer's Disease and Hypothesis of Functional Role. *Int J Mol Sci* **23**, doi:10.3390/ijms23073629 (2022).
278 Ramsden, C. E. *et al.* Lipid Peroxidation Induced ApoE Receptor-Ligand Disruption as a Unifying Hypothesis Underlying Sporadic Alzheimer's Disease in Humans. *J Alzheimers Dis* **87**, 1251-1290, doi:10.3233/jad-220071 (2022).
279 Murchison, A. G. Hypothesis: Modulation of microglial phenotype in Alzheimer's disease drives neurodegeneration. *Alzheimers Dement* **18**, 1537-1544, doi:10.1002/alz.12503 (2022).
280 Martens, Y. A. *et al.* ApoE Cascade Hypothesis in the pathogenesis of Alzheimer's disease and related dementias. *Neuron* **110**, 1304-1317, doi:10.1016/j.neuron.2022.03.004 (2022).
281 Wainberg, M. *et al.* The viral hypothesis: how herpesviruses may contribute to Alzheimer's disease. *Mol Psychiatry* **26**, 5476-5480, doi:10.1038/s41380-021-01138-6 (2021).
282 Pal, A. *et al.* Microglia and Astrocytes in Alzheimer's Disease in the Context of the Aberrant Copper Homeostasis Hypothesis. *Biomolecules* **11**, doi:10.3390/biom11111598 (2021).
283 Naughton, S. X., Raval, U. & Pasinetti, G. M. The Viral Hypothesis in Alzheimer's Disease: Novel Insights and Pathogen-Based Biomarkers. *J Pers Med* **10**, doi:10.3390/jpm10030074 (2020).
284 Edwards, F. A. A Unifying Hypothesis for Alzheimer's Disease: From Plaques to Neurodegeneration. *Trends Neurosci* **42**, 310-322, doi:10.1016/j.tins.2019.03.003 (2019).
285 Chen, S., Bonifati, S., Qin, Z., St Gelais, C. & Wu, L. SAMHD1 Suppression of Antiviral Immune Responses. *Trends Microbiol* **27**, 254-267, doi:10.1016/j.tim.2018.09.009 (2019).
286 Rasmussen, K. L., Nordestgaard, B. G., Frikke-Schmidt, R. & Nielsen, S. F. An updated Alzheimer hypothesis: Complement C3 and risk of Alzheimer's disease-A cohort study of 95,442 individuals. *Alzheimers Dement* **14**, 1589-1601, doi:10.1016/j.jalz.2018.07.223 (2018).

287 Devanand, D. P. Viral Hypothesis and Antiviral Treatment in Alzheimer's Disease. *Curr Neurol Neurosci Rep* **18**, 55, doi:10.1007/s11910-018-0863-1 (2018).
288 Chen, S. *et al.* SAMHD1 suppresses innate immune responses to viral infections and inflammatory stimuli by inhibiting the NF-κB and interferon pathways. *Proc Natl Acad Sci U S A* **115**, E3798-e3807, doi:10.1073/pnas.1801213115 (2018).
289 Sánchez-Valle, J. *et al.* A molecular hypothesis to explain direct and inverse co-morbidities between Alzheimer's Disease, Glioblastoma and Lung cancer. *Sci Rep* **7**, 4474, doi:10.1038/s41598-017-04400-6 (2017).
290 Marchesi, V. T. Gain-of-function somatic mutations contribute to inflammation and blood vessel damage that lead to Alzheimer dementia: a hypothesis. *Faseb j* **30**, 503-506, doi:10.1096/fj.15-282285 (2016).
291 Bozluolcay, M., Andican, G., Fırtına, S., Erkol, G. & Konukoglu, D. Inflammatory hypothesis as a link between Alzheimer's disease and diabetes mellitus. *Geriatr Gerontol Int* **16**, 1161-1166, doi:10.1111/ggi.12602 (2016).
292 Maggioli, E. *et al.* The human leukocyte antigen class III haplotype approach: new insight in Alzheimer's disease inflammation hypothesis. *Curr Alzheimer Res* **10**, 1047-1056, doi:10.2174/15672050113106660169 (2013).
293 Kamer, A. R. *et al.* Alzheimer's disease and peripheral infections: the possible contribution from periodontal infections, model and hypothesis. *J Alzheimers Dis* **13**, 437-449, doi:10.3233/jad-2008-13408 (2008).
294 Fernández, J. A., Rojo, L., Kuljis, R. O. & Maccioni, R. B. The damage signals hypothesis of Alzheimer's disease pathogenesis. *J Alzheimers Dis* **14**, 329-333, doi:10.3233/jad-2008-14307 (2008).
295 Aisen, P. S. The inflammatory hypothesis of Alzheimer disease: dead or alive? *Alzheimer Dis Assoc Disord* **22**, 4-5, doi:10.1097/WAD.0b013e318166ca4c (2008).
296 Arshavsky, Y. I. Alzheimer's disease, brain immune privilege and memory: a hypothesis. *J Neural Transm (Vienna)* **113**, 1697-1707, doi:10.1007/s00702-006-0524-4 (2006).
297 Rogers, J. An IL-1 alpha susceptibility polymorphism in Alzheimer's disease:new fuel for the inflammation hypothesis. *Neurology* **55**, 464-465, doi:10.1212/wnl.55.4.464 (2000).
298 Armstrong, R. A., Winsper, S. J. & Blair, J. A. Hypothesis: is Alzheimer's disease a metal-induced immune disorder? *Neurodegeneration* **4**, 107-111, doi:10.1006/neur.1995.0013 (1995).
299 McRae, A. & Dahlström, A. Immune responses in brains of Alzheimer's and Parkinson's disease patients: hypothesis and reality. *Rev Neurosci* **3**, 79-98, doi:10.1515/revneuro.1992.3.2.79 (1992).
300 Friedland, R. P., May, C. & Dahlberg, J. The viral hypothesis of Alzheimer's disease. Absence of antibodies to lentiviruses. *Arch Neurol* **47**, 177-178, doi:10.1001/archneur.1990.00530020083019 (1990).
301 Li, J., Sun, M., Cui, X. & Li, C. Protective Effects of Flavonoids against Alzheimer's Disease: Pathological Hypothesis, Potential Targets, and Structure-Activity Relationship. *Int J Mol Sci* **23**, doi:10.3390/ijms231710020 (2022).
302 Teixeira, J. P., de Castro, A. A., Soares, F. V., da Cunha, E. F. F. & Ramalho, T. C. Future Therapeutic Perspectives into the Alzheimer's Disease Targeting the Oxidative Stress Hypothesis. *Molecules* **24**, doi:10.3390/molecules24234410 (2019).

303 Akilo, O. D. *et al.* Hypothesis: apo-lactoferrin-Galantamine Proteo-alkaloid Conjugate for Alzheimer's disease Intervention. *J Cell Mol Med* **22**, 1957-1963, doi:10.1111/jcmm.13484 (2018).
304 Hofer, T. & Perry, G. Nucleic acid oxidative damage in Alzheimer's disease-explained by the hepcidin-ferroportin neuronal iron overload hypothesis? *J Trace Elem Med Biol* **38**, 1-9, doi:10.1016/j.jtemb.2016.06.005 (2016).
305 Ulusu, N. N. Glucose-6-phosphate dehydrogenase deficiency and Alzheimer's disease: Partners in crime? The hypothesis. *Med Hypotheses* **85**, 219-223, doi:10.1016/j.mehy.2015.05.006 (2015).
306 Gezen-Ak, D., Yılmazer, S. & Dursun, E. Why vitamin D in Alzheimer's disease? The hypothesis. *J Alzheimers Dis* **40**, 257-269, doi:10.3233/jad-131970 (2014).
307 Padurariu, M. *et al.* The oxidative stress hypothesis in Alzheimer's disease. *Psychiatr Danub* **25**, 401-409 (2013).
308 Nunomura, A. [Oxidative stress hypothesis for Alzheimer's disease and its potential therapeutic implications]. *Rinsho Shinkeigaku* **53**, 1043-1045, doi:10.5692/clinicalneurol.53.1043 (2013).
309 Proctor, C. J. & Gray, D. A. A unifying hypothesis for familial and sporadic Alzheimer's disease. *Int J Alzheimers Dis* **2012**, 978742, doi:10.1155/2012/978742 (2012).
310 Praticò, D. Oxidative stress hypothesis in Alzheimer's disease: a reappraisal. *Trends Pharmacol Sci* **29**, 609-615, doi:10.1016/j.tips.2008.09.001 (2008).
311 Erol, A. An integrated and unifying hypothesis for the metabolic basis of sporadic Alzheimer's disease. *J Alzheimers Dis* **13**, 241-253, doi:10.3233/jad-2008-13302 (2008).
312 Zhu, X., Lee, H. G., Perry, G. & Smith, M. A. Alzheimer disease, the two-hit hypothesis: an update. *Biochim Biophys Acta* **1772**, 494-502, doi:10.1016/j.bbadis.2006.10.014 (2007).
313 Zhu, X., Raina, A. K., Perry, G. & Smith, M. A. Alzheimer's disease: the two-hit hypothesis. *Lancet Neurol* **3**, 219-226, doi:10.1016/s1474-4422(04)00707-0 (2004).
314 Barger, S. W. An unconventional hypothesis of oxidation in Alzheimer's disease: intersections with excitotoxicity. *Front Biosci* **9**, 3286-3295, doi:10.2741/1481 (2004).
315 Celec, P. & Príhodová, M. Pathogenesis of Alzheimer's disease and the role of heme oxygenase: new perspectives and hypothesis. *Folia Neuropathol* **41**, 155-160 (2003).
316 Zhu, X. *et al.* Differential activation of neuronal ERK, JNK/SAPK and p38 in Alzheimer disease: the 'two hit' hypothesis. *Mech Ageing Dev* **123**, 39-46, doi:10.1016/s0047-6374(01)00342-6 (2001).
317 Tabet, N., Mantle, D. & Orrell, M. Free radicals as mediators of toxicity in Alzheimer's disease: a review and hypothesis. *Adverse Drug React Toxicol Rev* **19**, 127-152 (2000).
318 Harman, D. Alzheimer's disease: A hypothesis on pathogenesis. *J Am Aging Assoc* **23**, 147-161, doi:10.1007/s11357-000-0017-6 (2000).
319 Van Dyke, K. The possible role of peroxynitrite in Alzheimer's disease: a simple hypothesis that could be tested more thoroughly. *Med Hypotheses* **48**, 375-380, doi:10.1016/s0306-9877(97)90031-1 (1997).
320 Markesbery, W. R. Oxidative stress hypothesis in Alzheimer's disease. *Free Radic Biol Med* **23**, 134-147, doi:10.1016/s0891-5849(96)00629-6 (1997).
321 Volicer, L. & Crino, P. B. Involvement of free radicals in dementia of the Alzheimer type: a hypothesis. *Neurobiol Aging* **11**, 567-571, doi:10.1016/0197-4580(90)90119-k (1990).

322 Henderson, A. S. The risk factors for Alzheimer's disease: a review and a hypothesis. *Acta Psychiatr Scand* **78**, 257-275, doi:10.1111/j.1600-0447.1988.tb06336.x (1988).
323 Wang, F. *et al.* Iron Dyshomeostasis and Ferroptosis: A New Alzheimer's Disease Hypothesis? *Front Aging Neurosci* **14**, 830569, doi:10.3389/fnagi.2022.830569 (2022).
324 Solioz, M. Low copper-2 intake in Switzerland does not result in lower incidence of Alzheimer's disease and contradicts the Copper-2 Hypothesis. *Exp Biol Med (Maywood)* **245**, 177-179, doi:10.1177/1535370219899898 (2020).
325 Siblerud, R., Mutter, J., Moore, E., Naumann, J. & Walach, H. A Hypothesis and Evidence That Mercury May be an Etiological Factor in Alzheimer's Disease. *Int J Environ Res Public Health* **16**, doi:10.3390/ijerph16245152 (2019).
326 Brewer, G. J. Copper-2 Hypothesis for Causation of the Current Alzheimer's Disease Epidemic Together with Dietary Changes That Enhance the Epidemic. *Chem Res Toxicol* **30**, 763-768, doi:10.1021/acs.chemrestox.6b00373 (2017).
327 Virk, S. A. & Eslick, G. D. Brief Report: Meta-analysis of Antacid Use and Alzheimer's Disease: Implications for the Aluminum Hypothesis. *Epidemiology* **26**, 769-773, doi:10.1097/ede.0000000000000326 (2015).
328 Liang, S. H. *et al.* Novel Fluorinated 8-Hydroxyquinoline Based Metal Ionophores for Exploring the Metal Hypothesis of Alzheimer's Disease. *ACS Med Chem Lett* **6**, 1025-1029, doi:10.1021/acsmedchemlett.5b00281 (2015).
329 Davenward, S. *et al.* Silicon-rich mineral water as a non-invasive test of the 'aluminum hypothesis' in Alzheimer's disease. *J Alzheimers Dis* **33**, 423-430, doi:10.3233/jad-2012-121231 (2013).
330 Squitti, R. & Polimanti, R. Copper hypothesis in the missing hereditability of sporadic Alzheimer's disease: ATP7B gene as potential harbor of rare variants. *J Alzheimers Dis* **29**, 493-501, doi:10.3233/jad-2011-111991 (2012).
331 Craddock, T. J. *et al.* The zinc dyshomeostasis hypothesis of Alzheimer's disease. *PLoS One* **7**, e33552, doi:10.1371/journal.pone.0033552 (2012).
332 Sabayan, B., Farshchi, S., Zamiri, N. & Sabayan, B. Can tetrathiomolybdate be a potential agent against Alzheimer disease? A hypothesis based on abnormal copper homeostasis in brain. *Alzheimer Dis Assoc Disord* **24**, 309-310, doi:10.1097/WAD.0b013e3181d5e5a3 (2010).
333 Bush, A. I. & Tanzi, R. E. Therapeutics for Alzheimer's disease based on the metal hypothesis. *Neurotherapeutics* **5**, 421-432, doi:10.1016/j.nurt.2008.05.001 (2008).
334 Bush, A. I. Drug development based on the metals hypothesis of Alzheimer's disease. *J Alzheimers Dis* **15**, 223-240, doi:10.3233/jad-2008-15208 (2008).
335 Bellés, M., Sánchez, D. J., Gómez, M., Corbella, J. & Domingo, J. L. Silicon reduces aluminum accumulation in rats: relevance to the aluminum hypothesis of Alzheimer disease. *Alzheimer Dis Assoc Disord* **12**, 83-87, doi:10.1097/00002093-199806000-00005 (1998).
336 Zatta, P. F. Aluminum binds to the hyperphosphorylated tau in Alzheimer's disease: a hypothesis. *Med Hypotheses* **44**, 169-172, doi:10.1016/0306-9877(95)90131-0 (1995).
337 McCaddon, A. & Kelly, C. L. Alzheimer's disease: a 'cobalaminergic' hypothesis. *Med Hypotheses* **37**, 161-165, doi:10.1016/0306-9877(92)90074-m (1992).

338 Glick, J. L. Dementias: the role of magnesium deficiency and an hypothesis concerning the pathogenesis of Alzheimer's disease. *Med Hypotheses* **31**, 211-225, doi:10.1016/0306-9877(90)90095-v (1990).
339 Zatta, P., Giordano, R., Corain, B. & Bombi, G. G. Alzheimer dementia and the aluminum hypothesis. *Med Hypotheses* **26**, 139-142, doi:10.1016/0306-9877(88)90068-0 (1988).
340 Olsen, I. & Singhrao, S. K. Low levels of salivary lactoferrin may affect oral dysbiosis and contribute to Alzheimer's disease: A hypothesis. *Med Hypotheses* **146**, 110393, doi:10.1016/j.mehy.2020.110393 (2021).
341 Li, Z. *et al.* The connectome from the cerebral cortex to the viscera using viral transneuronal tracers. *Am J Transl Res* **13**, 12152-12167 (2021).
342 Seaks, C. E. & Wilcock, D. M. Infectious hypothesis of Alzheimer disease. *PLoS Pathog* **16**, e1008596, doi:10.1371/journal.ppat.1008596 (2020).
343 Lathe, J. C. & Lathe, R. Evidence against a geographic gradient of Alzheimer's disease and the hygiene hypothesis. *Evol Med Public Health* **2020**, 141-144, doi:10.1093/emph/eoaa023 (2020).
344 Frölich, L. Alzheimer's disease - the 'microbial hypothesis' from a clinical and neuroimaging perspective. *Psychiatry Res Neuroimaging* **306**, 111181, doi:10.1016/j.pscychresns.2020.111181 (2020).
345 Nath, A. Herpes Viruses, Alzheimer's Disease, and Related Dementias: Unifying or Confusing Hypothesis? *Neurotherapeutics* **16**, 180-181, doi:10.1007/s13311-018-00701-4 (2019).
346 Mielcarska, M. B. *et al.* Syk and Hrs Regulate TLR3-Mediated Antiviral Response in Murine Astrocytes. *Oxid Med Cell Longev* **2019**, 6927380, doi:10.1155/2019/6927380 (2019).
347 Moir, R. D., Lathe, R. & Tanzi, R. E. The antimicrobial protection hypothesis of Alzheimer's disease. *Alzheimers Dement* **14**, 1602-1614, doi:10.1016/j.jalz.2018.06.3040 (2018).
348 Contaldi, F. *et al.* Author Correction: The hypothesis that Helicobacter pylori predisposes to Alzheimer's disease is biologically plausible. *Sci Rep* **8**, 6061, doi:10.1038/s41598-018-23613-x (2018).
349 Rubin, K. & Glazer, S. The pertussis hypothesis: Bordetella pertussis colonization in the pathogenesis of Alzheimer's disease. *Immunobiology* **222**, 228-240, doi:10.1016/j.imbio.2016.09.017 (2017).
350 Contaldi, F. *et al.* The hypothesis that Helicobacter pylori predisposes to Alzheimer's disease is biologically plausible. *Sci Rep* **7**, 7817, doi:10.1038/s41598-017-07532-x (2017).
351 Roubaud Baudron, C., Varon, C., Mégraud, F. & Salles, N. [Alzheimer's disease: the infectious hypothesis]. *Geriatr Psychol Neuropsychiatr Vieil* **13**, 418-424, doi:10.1684/pnv.2015.0574 (2015).
352 Piacentini, R. *et al.* HSV-1 and Alzheimer's disease: more than a hypothesis. *Front Pharmacol* **5**, 97, doi:10.3389/fphar.2014.00097 (2014).
353 Robinson, S. R., Dobson, C. & Lyons, J. Challenges and directions for the pathogen hypothesis of Alzheimer's disease. *Neurobiol Aging* **25**, 629-637, doi:10.1016/j.neurobiolaging.2003.12.022 (2004).

354 Ryu, W. I., Cohen, B. M. & Sonntag, K. C. Hypothesis and Theory: Characterizing Abnormalities of Energy Metabolism Using a Cellular Platform as a Personalized Medicine Approach for Alzheimer's Disease. *Front Cell Dev Biol* **9**, 697578, doi:10.3389/fcell.2021.697578 (2021).
355 Swerdlow, R. H. The mitochondrial hypothesis: Dysfunction, bioenergetic defects, and the metabolic link to Alzheimer's disease. *Int Rev Neurobiol* **154**, 207-233, doi:10.1016/bs.irn.2020.01.008 (2020).
356 Kosenko, E., Tikhonova, L., Alilova, G., Urios, A. & Montoliu, C. The Erythrocytic Hypothesis of Brain Energy Crisis in Sporadic Alzheimer Disease: Possible Consequences and Supporting Evidence. *J Clin Med* **9**, doi:10.3390/jcm9010206 (2020).
357 Ebanks, B., Ingram, T. L. & Chakrabarti, L. ATP synthase and Alzheimer's disease: putting a spin on the mitochondrial hypothesis. *Aging (Albany NY)* **12**, 16647-16662, doi:10.18632/aging.103867 (2020).
358 Müller, W. E. *et al.* Therapeutic efficacy of the Ginkgo special extract EGb761(®) within the framework of the mitochondrial cascade hypothesis of Alzheimer's disease. *World J Biol Psychiatry* **20**, 173-189, doi:10.1080/15622975.2017.1308552 (2019).
359 Blonz, E. R. Alzheimer's Disease as the Product of a Progressive Energy Deficiency Syndrome in the Central Nervous System: The Neuroenergetic Hypothesis. *J Alzheimers Dis* **60**, 1223-1229, doi:10.3233/jad-170549 (2017).
360 Area-Gomez, E. & Schon, E. A. On the Pathogenesis of Alzheimer's Disease: The MAM Hypothesis. *Faseb j* **31**, 864-867, doi:10.1096/fj.201601309 (2017).
361 Swerdlow, R. H., Burns, J. M. & Khan, S. M. The Alzheimer's disease mitochondrial cascade hypothesis: progress and perspectives. *Biochim Biophys Acta* **1842**, 1219-1231, doi:10.1016/j.bbadis.2013.09.010 (2014).
362 Swerdlow, R. H., Burns, J. M. & Khan, S. M. The Alzheimer's disease mitochondrial cascade hypothesis. *J Alzheimers Dis* **20 Suppl 2**, S265-279, doi:10.3233/jad-2010-100339 (2010).
363 Agnati, L. F. *et al.* A new hypothesis of pathogenesis based on the divorce between mitochondria and their host cells: possible relevance for Alzheimer's disease. *Curr Alzheimer Res* **7**, 307-322, doi:10.2174/156720510791162395 (2010).
364 Swerdlow, R. H. & Khan, S. M. The Alzheimer's disease mitochondrial cascade hypothesis: an update. *Exp Neurol* **218**, 308-315, doi:10.1016/j.expneurol.2009.01.011 (2009).
365 Roses, A. D. *et al.* Complex disease-associated pharmacogenetics: drug efficacy, drug safety, and confirmation of a pathogenetic hypothesis (Alzheimer's disease). *Pharmacogenomics J* **7**, 10-28, doi:10.1038/sj.tpj.6500397 (2007).
366 Mancuso, M., Coppedè, F., Murri, L. & Siciliano, G. Mitochondrial cascade hypothesis of Alzheimer's disease: myth or reality? *Antioxid Redox Signal* **9**, 1631-1646, doi:10.1089/ars.2007.1761 (2007).
367 Sager, M. A. & Johnson, S. C. Commentary on "Perspective on a pathogenesis and treatment of Alzheimer's disease." Comment on the mitochondrial metabolism hypothesis. *Alzheimers Dement* **2**, 74-75, doi:10.1016/j.jalz.2005.12.002 (2006).

368 Huang, Y. & Mahley, R. W. Commentary on "Perspective on a pathogenesis and treatment of Alzheimer's disease." Apolipoprotein E and the mitochondrial metabolic hypothesis. *Alzheimers Dement* **2**, 71-73, doi:10.1016/j.jalz.2005.12.006 (2006).
369 Swerdlow, R. H. & Khan, S. M. A "mitochondrial cascade hypothesis" for sporadic Alzheimer's disease. *Med Hypotheses* **63**, 8-20, doi:10.1016/j.mehy.2003.12.045 (2004).
370 Alexander, G. E. *et al.* Association of premorbid intellectual function with cerebral metabolism in Alzheimer's disease: implications for the cognitive reserve hypothesis. *Am J Psychiatry* **154**, 165-172, doi:10.1176/ajp.154.2.165 (1997).
371 Davis, J. N., Hunnicutt, E. J., Jr. & Chisholm, J. C. A mitochondrial bottleneck hypothesis of Alzheimer's disease. *Mol Med Today* **1**, 240-247, doi:10.1016/s1357-4310(95)91532-x (1995).
372 Scheffer, S., Hermkens, D. M. A., van der Weerd, L., de Vries, H. E. & Daemen, M. Vascular Hypothesis of Alzheimer Disease: Topical Review of Mouse Models. *Arterioscler Thromb Vasc Biol* **41**, 1265-1283, doi:10.1161/atvbaha.120.311911 (2021).
373 Fujino, T., Hossain, M. S. & Mawatari, S. Therapeutic Efficacy of Plasmalogens for Alzheimer's Disease, Mild Cognitive Impairment, and Parkinson's Disease in Conjunction with a New Hypothesis for the Etiology of Alzheimer's Disease. *Adv Exp Med Biol* **1299**, 195-212, doi:10.1007/978-3-030-60204-8_14 (2020).
374 Kehoe, P. G. The Coming of Age of the Angiotensin Hypothesis in Alzheimer's Disease: Progress Toward Disease Prevention and Treatment? *J Alzheimers Dis* **62**, 1443-1466, doi:10.3233/jad-171119 (2018).
375 de la Torre, J. The Vascular Hypothesis of Alzheimer's Disease: A Key to Preclinical Prediction of Dementia Using Neuroimaging. *J Alzheimers Dis* **63**, 35-52, doi:10.3233/jad-180004 (2018).
376 Østergaard, L. *et al.* The capillary dysfunction hypothesis of Alzheimer's disease. *Neurobiol Aging* **34**, 1018-1031, doi:10.1016/j.neurobiolaging.2012.09.011 (2013).
377 Kehoe, P. G. & Passmore, P. A. The renin-angiotensin system and antihypertensive drugs in Alzheimer's disease: current standing of the angiotensin hypothesis? *J Alzheimers Dis* **30 Suppl 2**, S251-268, doi:10.3233/jad-2012-111376 (2012).
378 Fang, B., Wang, D., Huang, M., Yu, G. & Li, H. Hypothesis on the relationship between the change in intracellular pH and incidence of sporadic Alzheimer's disease or vascular dementia. *Int J Neurosci* **120**, 591-595, doi:10.3109/00207454.2010.505353 (2010).
379 de la Torre, J. C. The vascular hypothesis of Alzheimer's disease: bench to bedside and beyond. *Neurodegener Dis* **7**, 116-121, doi:10.1159/000285520 (2010).
380 Milionis, H. J., Florentin, M. & Giannopoulos, S. Metabolic syndrome and Alzheimer's disease: a link to a vascular hypothesis? *CNS Spectr* **13**, 606-613, doi:10.1017/s1092852900016886 (2008).
381 Birkenhäger, W. H. & Staessen, J. A. Is angiogenesis a plausible hypothesis in Alzheimer's disease? *J Hypertens* **21**, 1426-1427, doi:10.1097/00004872-200307000-00036 (2003).
382 de la Torre, J. C. Critically attained threshold of cerebral hypoperfusion: the CATCH hypothesis of Alzheimer's pathogenesis. *Neurobiol Aging* **21**, 331-342, doi:10.1016/s0197-4580(00)00111-1 (2000).

383 Arnsten, A. F. T., Datta, D., Del Tredici, K. & Braak, H. Hypothesis: Tau pathology is an initiating factor in sporadic Alzheimer's disease. *Alzheimers Dement* **17**, 115-124, doi:10.1002/alz.12192 (2021).
384 Sonawane, S. K. & Chinnathambi, S. Prion-Like Propagation of Post-Translationally Modified Tau in Alzheimer's Disease: A Hypothesis. *J Mol Neurosci* **65**, 480-490, doi:10.1007/s12031-018-1111-5 (2018).
385 Maccioni, R. B., Farías, G., Morales, I. & Navarrete, L. The revitalized tau hypothesis on Alzheimer's disease. *Arch Med Res* **41**, 226-231, doi:10.1016/j.arcmed.2010.03.007 (2010).
386 Hooper, C., Killick, R. & Lovestone, S. The GSK3 hypothesis of Alzheimer's disease. *J Neurochem* **104**, 1433-1439, doi:10.1111/j.1471-4159.2007.05194.x (2008).
387 Iqbal, K. & Grundke-Iqbal, I. Metabolic/signal transduction hypothesis of Alzheimer's disease and other tauopathies. *Acta Neuropathol* **109**, 25-31, doi:10.1007/s00401-004-0951-y (2005).
388 Chen, M. The Alzheimer's plaques, tangles and memory deficits may have a common origin; part I; a calcium deficit hypothesis. *Front Biosci* **3**, a27-31, doi:10.2741/a248 (1998).
389 Rudge, J. D. A New Hypothesis for Alzheimer's Disease: The Lipid Invasion Model. *J Alzheimers Dis Rep* **6**, 129-161, doi:10.3233/adr-210299 (2022).
390 Needham, H. *et al.* A Dichotomous Role for FABP7 in Sleep and Alzheimer's Disease Pathogenesis: A Hypothesis. *Front Neurosci* **16**, 798994, doi:10.3389/fnins.2022.798994 (2022).
391 Martinez, A. E. *et al.* The small HDL particle hypothesis of Alzheimer's disease. *Alzheimers Dement*, doi:10.1002/alz.12649 (2022).
392 Lyssenko, N. N. & Praticò, D. ABCA7 and the altered lipidostasis hypothesis of Alzheimer's disease. *Alzheimers Dement* **17**, 164-174, doi:10.1002/alz.12220 (2021).
393 Yu, Q. & Zhong, C. Membrane Aging as the Real Culprit of Alzheimer's Disease: Modification of a Hypothesis. *Neurosci Bull* **34**, 369-381, doi:10.1007/s12264-017-0192-4 (2018).
394 Nixon, D. W. Down Syndrome, Obesity, Alzheimer's Disease, and Cancer: A Brief Review and Hypothesis. *Brain Sci* **8**, doi:10.3390/brainsci8040053 (2018).
395 Wood, W. G., Li, L., Müller, W. E. & Eckert, G. P. Cholesterol as a causative factor in Alzheimer's disease: a debatable hypothesis. *J Neurochem* **129**, 559-572, doi:10.1111/jnc.12637 (2014).
396 Fowler, C. J. The role of the phosphoinositide signalling system in the pathogenesis of sporadic Alzheimer's disease: a hypothesis. *Brain Res Brain Res Rev* **25**, 373-380, doi:10.1016/s0165-0173(97)00024-6 (1997).
397 Peers, R. J. Alzheimer's disease and omega-3 fatty acids: hypothesis. *Med J Aust* **153**, 563-564 (1990).
398 Bertoni-Freddari, C. Age-dependent deterioration of neuronal membranes and the pathogenesis of Alzheimer's disease: a hypothesis. *Med Hypotheses* **25**, 147-149, doi:10.1016/0306-9877(88)90052-7 (1988).
399 O'Day, D. H. Alzheimer's Disease: A short introduction to the calmodulin hypothesis. *AIMS Neurosci* **6**, 231-239, doi:10.3934/Neuroscience.2019.4.231 (2019).

400 Overk, C. & Masliah, E. Perspective on the calcium dyshomeostasis hypothesis in the pathogenesis of selective neuronal degeneration in animal models of Alzheimer's disease. *Alzheimers Dement* **13**, 183-185, doi:10.1016/j.jalz.2017.01.005 (2017).
401 Calcium Hypothesis of Alzheimer's disease and brain aging: A framework for integrating new evidence into a comprehensive theory of pathogenesis. *Alzheimers Dement* **13**, 178-182.e117, doi:10.1016/j.jalz.2016.12.006 (2017).
402 Yamashima, T. Can 'calpain-cathepsin hypothesis' explain Alzheimer neuronal death? *Ageing Res Rev* **32**, 169-179, doi:10.1016/j.arr.2016.05.008 (2016).
403 Chen, M. & Nguyen, H. T. Our "energy-Ca(2+) signaling deficits" hypothesis and its explanatory potential for key features of Alzheimer's disease. *Front Aging Neurosci* **6**, 329, doi:10.3389/fnagi.2014.00329 (2014).
404 Yamashima, T. Reconsider Alzheimer's disease by the 'calpain-cathepsin hypothesis'--a perspective review. *Prog Neurobiol* **105**, 1-23, doi:10.1016/j.pneurobio.2013.02.004 (2013).
405 Riazantseva, M. A., Mozhaeva, G. N. & Kaznacheeva, E. V. [Calcium hypothesis of Alzheimer disease]. *Usp Fiziol Nauk* **43**, 59-72 (2012).
406 Berridge, M. J. Calcium hypothesis of Alzheimer's disease. *Pflugers Arch* **459**, 441-449, doi:10.1007/s00424-009-0736-1 (2010).
407 Thibault, O., Gant, J. C. & Landfield, P. W. Expansion of the calcium hypothesis of brain aging and Alzheimer's disease: minding the store. *Aging Cell* **6**, 307-317, doi:10.1111/j.1474-9726.2007.00295.x (2007).
408 O'Day, D. H. & Myre, M. A. Calmodulin-binding domains in Alzheimer's disease proteins: extending the calcium hypothesis. *Biochem Biophys Res Commun* **320**, 1051-1054, doi:10.1016/j.bbrc.2004.06.070 (2004).
409 The current status of the calcium hypothesis of brain aging and Alzheimer's disease. Heidelberg, Germany, October 23-25, 1995. Proceedings of a conference. *Life Sci* **59**, vii, 357-510 (1996).
410 Khachaturian, Z. S. Calcium hypothesis of Alzheimer's disease and brain aging. *Ann N Y Acad Sci* **747**, 1-11, doi:10.1111/j.1749-6632.1994.tb44398.x (1994).
411 Atwood, C. S. & Bowen, R. L. A Unified Hypothesis of Early- and Late-Onset Alzheimer's Disease Pathogenesis. *J Alzheimers Dis* **47**, 33-47, doi:10.3233/jad-143210 (2015).
412 Hunter, S., Arendt, T. & Brayne, C. The senescence hypothesis of disease progression in Alzheimer disease: an integrated matrix of disease pathways for FAD and SAD. *Mol Neurobiol* **48**, 556-570, doi:10.1007/s12035-013-8445-3 (2013).
413 Yurov, Y. B., Vorsanova, S. G. & Iourov, I. Y. The DNA replication stress hypothesis of Alzheimer's disease. *ScientificWorldJournal* **11**, 2602-2612, doi:10.1100/2011/625690 (2011).
414 Frade, J. M. & López-Sánchez, N. A novel hypothesis for Alzheimer disease based on neuronal tetraploidy induced by p75 (NTR). *Cell Cycle* **9**, 1934-1941, doi:10.4161/cc.9.10.11582 (2010).
415 Golde, T. E. & Miller, V. M. Proteinopathy-induced neuronal senescence: a hypothesis for brain failure in Alzheimer's and other neurodegenerative diseases. *Alzheimers Res Ther* **1**, 5, doi:10.1186/alzrt5 (2009).

416 Woods, J., Snape, M. & Smith, M. A. The cell cycle hypothesis of Alzheimer's disease: suggestions for drug development. *Biochim Biophys Acta* **1772**, 503-508, doi:10.1016/j.bbadis.2006.12.004 (2007).
417 Marshak, D. R. & Peña, L. A. Potential role of S100 beta in Alzheimer's disease: an hypothesis involving mitotic protein kinases. *Prog Clin Biol Res* **379**, 289-307 (1992).
418 Movassat, J., Delangre, E., Liu, J., Gu, Y. & Janel, N. Hypothesis and Theory: Circulating Alzheimer's-Related Biomarkers in Type 2 Diabetes. Insight From the Goto-Kakizaki Rat. *Front Neurol* **10**, 649, doi:10.3389/fneur.2019.00649 (2019).
419 Morgen, K. & Frölich, L. The metabolism hypothesis of Alzheimer's disease: from the concept of central insulin resistance and associated consequences to insulin therapy. *J Neural Transm (Vienna)* **122**, 499-504, doi:10.1007/s00702-015-1377-5 (2015).
420 Domínguez, R. O. *et al.* Alzheimer disease and cognitive impairment associated with diabetes mellitus type 2: associations and a hypothesis. *Neurologia* **29**, 567-572, doi:10.1016/j.nrl.2013.05.006 (2014).
421 Kuljiš, R. O. & Salković-Petrišić, M. Dementia, diabetes, Alzheimer's disease, and insulin resistance in the brain: progress, dilemmas, new opportunities, and a hypothesis to tackle intersecting epidemics. *J Alzheimers Dis* **25**, 29-41, doi:10.3233/jad-2011-101392 (2011).
422 Rasgon, N. L. & Kenna, H. A. Insulin resistance in depressive disorders and Alzheimer's disease: revisiting the missing link hypothesis. *Neurobiol Aging* **26 Suppl 1**, 103-107, doi:10.1016/j.neurobiolaging.2005.09.004 (2005).
423 Rasgon, N. & Jarvik, L. Insulin resistance, affective disorders, and Alzheimer's disease: review and hypothesis. *J Gerontol A Biol Sci Med Sci* **59**, 178-183; discussion 184-192, doi:10.1093/gerona/59.2.m178 (2004).
424 Hoyer, S. Is sporadic Alzheimer disease the brain type of non-insulin dependent diabetes mellitus? A challenging hypothesis. *J Neural Transm (Vienna)* **105**, 415-422, doi:10.1007/s007020050067 (1998).
425 Brokaw, D. L. *et al.* Cell death and survival pathways in Alzheimer's disease: an integrative hypothesis testing approach utilizing -omic data sets. *Neurobiol Aging* **95**, 15-25, doi:10.1016/j.neurobiolaging.2020.06.022 (2020).
426 Offringa-Hup, A. Alzheimer's disease: The derailed repair hypothesis. *Med Hypotheses* **136**, 109516, doi:10.1016/j.mehy.2019.109516 (2020).
427 Kaeser, G. E. & Chun, J. Mosaic Somatic Gene Recombination as a Potentially Unifying Hypothesis for Alzheimer's Disease. *Front Genet* **11**, 390, doi:10.3389/fgene.2020.00390 (2020).
428 Kimura, N. & Yanagisawa, K. Traffic jam hypothesis: Relationship between endocytic dysfunction and Alzheimer's disease. *Neurochem Int* **119**, 35-41, doi:10.1016/j.neuint.2017.07.002 (2018).
429 Funk, K. E. & Kuret, J. Lysosomal fusion dysfunction as a unifying hypothesis for Alzheimer's disease pathology. *Int J Alzheimers Dis* **2012**, 752894, doi:10.1155/2012/752894 (2012).
430 Snow, A. D., Cummings, J. A. & Lake, T. The Unifying Hypothesis of Alzheimer's Disease: Heparan Sulfate Proteoglycans/Glycosaminoglycans Are Key as First Hypothesized Over 30 Years Ago. *Front Aging Neurosci* **13**, 710683, doi:10.3389/fnagi.2021.710683 (2021).

431 Strittmatter, W. J. *et al.* Hypothesis: microtubule instability and paired helical filament formation in the Alzheimer disease brain are related to apolipoprotein E genotype. *Exp Neurol* **125**, 163-171; discussion 172-164, doi:10.1006/exnr.1994.1019 (1994).
432 Celesia, G. G. Alzheimer's disease: the proteoglycans hypothesis. *Semin Thromb Hemost* **17 Suppl 2**, 158-160 (1991).
433 Matsuyama, S. S. & Jarvik, L. F. Hypothesis: microtubules, a key to Alzheimer disease. *Proc Natl Acad Sci U S A* **86**, 8152-8156, doi:10.1073/pnas.86.20.8152 (1989).
434 Sambamurti, K. *et al.* A partial failure of membrane protein turnover may cause Alzheimer's disease: a new hypothesis. *Curr Alzheimer Res* **3**, 81-90, doi:10.2174/156720506775697142 (2006).
435 Orpiszewski, J., Schormann, N., Kluve-Beckerman, B., Liepnieks, J. J. & Benson, M. D. Protein aging hypothesis of Alzheimer disease. *Faseb j* **14**, 1255-1263, doi:10.1096/fasebj.14.9.1255 (2000).
436 Liautard, J. P. A hypothesis on the aetiology of Alzheimer's disease: description of a model involving a misfolded chaperone. *Med Hypotheses* **43**, 372-380, doi:10.1016/0306-9877(94)90012-4 (1994).
437 Robakis, N. K. An Alzheimer's disease hypothesis based on transcriptional dysregulation. *Amyloid* **10**, 80-85, doi:10.3109/13506120309041729 (2003).
